# Risk Factors Associated with Post-Acute Sequelae of SARS-CoV-2 in an EHR Cohort: A National COVID Cohort Collaborative (N3C) Analysis as part of the NIH RECOVER program

**DOI:** 10.1101/2022.08.15.22278603

**Authors:** Elaine Hill, Hemal Mehta, Suchetha Sharma, Klint Mane, Catherine Xie, Emily Cathey, Johanna Loomba, Seth Russell, Heidi Spratt, Peter E. DeWitt, Nariman Ammar, Charisse Madlock-Brown, Donald Brown, Julie A. McMurry, Christopher G. Chute, Melissa A. Haendel, Richard Moffitt, Emily R. Pfaff, Tellen D. Bennett, the N3C Consortium, the RECOVER Consortium

## Abstract

**Background:** More than one-third of individuals experience post-acute sequelae of SARS-CoV-2 infection (PASC, which includes long-COVID).

**Objective:** To identify risk factors associated with PASC/long-COVID.

**Design:** Retrospective case-control study.

**Setting:** 31 health systems in the United States from the National COVID Cohort Collaborative (N3C).

**Patients:** 8,325 individuals with PASC (defined by the presence of the International Classification of Diseases, version 10 code U09.9 or a long-COVID clinic visit) matched to 41,625 controls within the same health system.

**Measurements:** Risk factors included demographics, comorbidities, and treatment and acute characteristics related to COVID-19. Multivariable logistic regression, random forest, and XGBoost were used to determine the associations between risk factors and PASC.

**Results:** Among 8,325 individuals with PASC, the majority were >50 years of age (56.6%), female (62.8%), and non-Hispanic White (68.6%). In logistic regression, middle-age categories (40 to 69 years; OR ranging from 2.32 to 2.58), female sex (OR 1.4, 95% CI 1.33-1.48), hospitalization associated with COVID-19 (OR 3.8, 95% CI 3.05-4.73), long (8-30 days, OR 1.69, 95% CI 1.31-2.17) or extended hospital stay (30+ days, OR 3.38, 95% CI 2.45-4.67), receipt of mechanical ventilation (OR 1.44, 95% CI 1.18-1.74), and several comorbidities including depression (OR 1.50, 95% CI 1.40-1.60), chronic lung disease (OR 1.63, 95% CI 1.53-1.74), and obesity (OR 1.23, 95% CI 1.16-1.3) were associated with increased likelihood of PASC diagnosis or care at a long-COVID clinic. Characteristics associated with a lower likelihood of PASC diagnosis or care at a long-COVID clinic included younger age (18 to 29 years), male sex, non-Hispanic Black race, and comorbidities such as substance abuse, cardiomyopathy, psychosis, and dementia. More doctors per capita in the county of residence was associated with an increased likelihood of PASC diagnosis or care at a long-COVID clinic. Our findings were consistent in sensitivity analyses using a variety of analytic techniques and approaches to select controls.

**Conclusions:** This national study identified important risk factors for PASC such as middle age, severe COVID-19 disease, and specific comorbidities. Further clinical and epidemiological research is needed to better understand underlying mechanisms and the potential role of vaccines and therapeutics in altering PASC course.

**KEY POINTS:** *Question:* What risk factors are associated with post-acute sequelae of SARS-CoV-2 (PASC) in the National COVID Cohort Collaborative (N3C) EHR Cohort?

*Findings:* This national study identified important risk factors for PASC such as middle age, severe COVID-19 disease, specific comorbidities, and the number of physicians per capita.

*Meaning:* Clinicians can use these risk factors to identify patients at high risk for PASC while they are still in the acute phase of their infection and also to support targeted enrollment in clinical trials for preventing or treating PASC.

## INTRODUCTION

Globally, over 500 million individuals have confirmed cases of COVID-19, including 86 million in the United States (U.S.) [1,2]. Although COVID-19 has resulted in short-term complications and deaths [3], long-term consequences are poorly understood. Many of those infected have developed long-term complications, commonly known as post-acute sequelae of SARS-CoV-2 infection (PASC) or long-COVID. The World Health Organization (WHO) defines long-COVID as the illness that occurs in people with a history of probable or confirmed SARS-CoV-2 infection, usually within 3 months from the onset of COVID-19 with symptoms that last for at least 2 months [4]. Long-COVID symptoms and complications include fatigue, cognitive dysfunction, post-exertional malaise, shortness of breath, depression, and many others [5,6]. Although it is difficult to estimate the true rate of PASC or long-COVID, nearly one-third of individuals in the U.S. have long-COVID [7–9].

Considerable research effort is geared toward identifying risk factors for PASC. Studies have identified that female sex, increased age, greater viral load, severity of acute illness, and comorbidities are associated with an increased likelihood of PASC [10–12]. Although age >70 was associated with increased likelihood of PASC diagnosis, recent data suggests that younger people aged 35 to 69 are at the highest risk of PASC [13]. The role of comorbidities in PASC risk needs to be explored in greater detail. Moreover, some prior studies relied on self-reported data captured through mobile app-based or web-based surveys, which can result in selection and responder bias [6,10]. Although social determinants of health (SDoH) such as poverty and access to healthcare are important risk factors for adverse COVID-19 outcomes, [14–17] their association with PASC is not well characterized [18], [19].

As a part of the NIH Researching COVID to Enhance Recovery (RECOVER) Initiative, we conducted this study to identify risk factors associated with PASC using the National COVID Cohort Collaborative (N3C) data, the largest publicly available electronic health records (EHRs) for COVID-19 in the U.S. We evaluated the association of demographic, comorbidity, clinical course, and patient-level SDoH factors on PASC risk.

## METHODS

### Data

N3C structure, access, and analytic capabilities have been described in detail previously [20]. The N3C collects information from single- and multi-hospital health systems across the U.S. and stores data in a central location, the N3C data enclave. As of April 14, 2022, it contained data from 72 health systems and >4.9 million individuals with COVID-19. For this study, we used a limited data set, which contains deidentified data, five-digit patient ZIP codes, and exact dates of COVID-19 diagnoses and service use **(eMethods)** [21].

### Study design and cohort (Figure 1)

The study cohort is based on 4,559,795 potentially eligible patients from 59 health systems who were diagnosed with SARS-CoV-2 infection or had a positive polymerase chain reaction (PCR) or antigen (AG) lab test for SARS-CoV-2. Of these, 3,884,477 were adults (>18 years of age). Individuals may have multiple SARS-CoV-2 infections, so we considered the earliest documented date of positive test or diagnosis as the COVID index date. An index date was required to determine the relative timing of infection and long-COVID diagnosis (International Classification of Diseases, Tenth Revision, Clinical Modification [ICD-10-CM] code U09.9) or long-COVID clinic visit. Not all health systems currently use U09.9 or have clinics dedicated to long-COVID treatment [22]. Therefore, we limited our cohort to patients from the 31 health systems with at least one documented long-COVID case using U09.9 or a long-COVID clinic visit between Oct 1, 2021 and Feb 28, 2022 (n=1,490,823). We excluded patients who died within 45 days of the index date because by definition they would not be at risk of developing PASC (n=1,467,804). Finally, in order for patients to have an adequate observation period after acute infection, we required them to have their index acute infection date between March 1, 2020 and December 1, 2021 (N=1,062,661). In this way, we employed a restrictive case definition to maximize the likelihood of selecting true cases of PASC from this base cohort.

**Figure 1.**
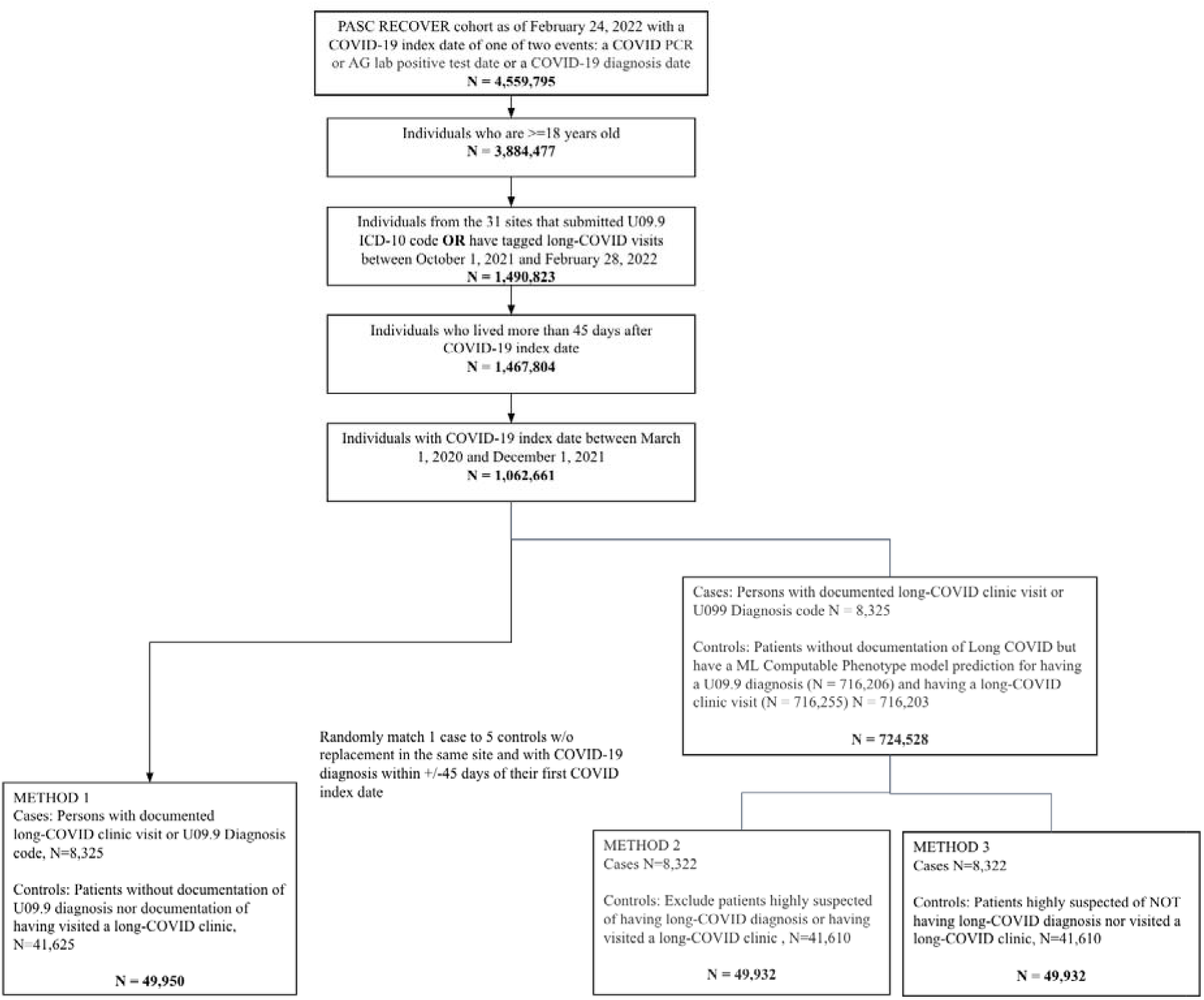
Cohort selection diagram

### Case and control selection

In our primary analyses, we defined cases as those with a documented U09.9 diagnosis or a documented long-COVID clinic visit flag in the N3C (n=8,325). As a sensitivity analysis, we also defined cases as 1) U09.9 only (n=7,512) or 2) long-COVID clinic visits only (n=1,241).

Controls were challenging to select because individuals may have had PASC but not received a diagnosis. We used three methods to identify controls, i.e., individuals without PASC. Our base analysis allowed any patient who was not a case to be considered as a possible matched control (not restricted controls). Additionally, for two control cohorts, we applied our previously developed computable phenotype (CP) model for long-COVID to refine our control patient pool [23]. We applied CP model to the 1,054,336 non-cases (1,062,661 - 8,325) to generate a predicted probability for U09.9 diagnosis or long-COVID clinic visit. The models generate the predicted probability of PASC for 716,203 individuals who became eligible for ***matched control selection* (eMethods)**.

1. Unrestricted controls (Method 1): All individuals who were not identified as cases became eligible (n=1,054,336).
2. Restricted controls (Method 2): We excluded individuals highly suspected of having long-COVID, defined as a predicted probability >= 0.75 based on the CP model of having a U09.9 diagnosis and having visited a long-COVID clinic. Overall, 621,374 individuals became eligible for controls.
3. More restricted controls (Method 3): We included individuals highly suspected of *<u>not</u>* having long-COVID (predicted probability <=0.25) based on the CP model of having a U09.9 diagnosis and a long-COVID clinic visit. Overall, 496,073 individuals became eligible for controls.

In each of the above three methods, we randomly matched 1 case to 5 controls without replacement from the same health system and COVID index date within +/- 45 days of the corresponding case’s earliest COVID index date. In the “unrestricted” method, We matched 8,325 cases to 41,625 controls in the “unrestricted” method, and 8,322 cases to 41,610 controls in the “restricted” and “more restricted controls” methods.

### Risk factors

We used existing literature [10–12], clinical expertise, and availability of information in the N3C to identify potential risk factors for PASC that are identifiable in EHR data (**Table 1** and Supplemental **eTable 1** for full list). We used information before COVID-19 diagnosis date to identify an individual’s age, gender, race/ethnicity (non-Hispanic White, non-Hispanic Black, Hispanic, Asians, others), obesity (a diagnosis of obesity or a body mass index [BMI]>=30), smoking status, substance abuse status, and comorbidities. We included 17 common comorbidities used in the Charlson Comorbidity Index [24] and additional comorbidities and treatments (e.g., use of corticosteroids) which are considered risk factors for severe acute COVID-19 as per the U.S. Centers for Disease Control (CDC) [25]. We also identified hospitalization for COVID-19, invasive mechanical ventilation use, extracorporeal membrane oxygenation (ECMO) use, vasopressor use, acute kidney injury diagnosis, sepsis diagnosis, remdesivir use, and total length of hospital stay **(eMethods)**.

**Table 1.** Cohort Characteristics for PASC Cases defined by U09.9 or long-COVID clinic visit.

|  | PASC<br>(N=8325)<sup>b</sup> | Method 1<br>Unrestricted<br>controls<br>(N=41625) | Method 2<br>Restricted<br>controls<br>(N=41610) | Method 3<br>Most restricted<br>controls<br>(N=41610) |
| --- | --- | --- | --- | --- |
| **Demographics** |  |  |  |  |
| Age (Mean[SD]) | 52.3 (15.5) | 47.5 (18.4) | 46.8 (17.8) | 48.1 (18.2) |
| Sex |  |  |  |  |
| Female | 5225 (62.8%) | 23090 (55.5%) | 24112 (57.9%) | 24530 (59.0%) |
| Male | 3096 (37.2%) | 18481 (44.4%) | 17482 (42.0%) | 17051 (41.0%) |
| Race/ethnicity |  |  |  |  |
| White non-Hispanic (NH) | 5707 (68.6%) | 26490 (63.6%) | 27818 (66.9%) | 27654 (66.5%) |
| Hispanic | 835 (10.0%) | 4851 (11.7%) | 4430 (10.6%) | 4452 (10.7%) |
| Black NH | 1235 (14.8%) | 6244 (15.0%) | 6455 (15.5%) | 6538 (15.7%) |
| Asian NH | 136 (1.6%) | 883 (2.1%) | 921 (2.2%) | 953 (2.3%) |
| Other race NH | 54 (0.6%) | 314 (0.8%) | 267 (0.6%) | 292 (0.7%) |
| **Comorbidities Prior to COVID Index Date** |  |  |  |  |
| Chronic Lung Disease | 2404 (28.9%) | 5717 (13.7%) | 6956 (16.7%) | 6816 (16.4%) |
| Complicated Diabetes | 1210 (14.5%) | 3582 (8.6%) | 4377 (10.5%) | 4336 (10.4%) |
| Congestive Heart Failure | 573 (6.9%) | 1530 (3.7%) | 2007 (4.8%) | 1910 (4.6%) |
| Hypertension | 3365 (40.4%) | 10894 (26.2%) | 13528 (32.5%) | 13698 (32.9%) |
| Kidney Disease | 1262 (15.2%) | 3616 (8.7%) | 4503 (10.8%) | 4388 (10.5%) |
| Obesity | 4691 (56.4%) | 16575 (39.8%) | 19588 (47.1%) | 19430 (46.7%) |
| Uncomplicated Diabetes | 1708 (20.5%) | 5547 (13.3%) | 6642 (16.0%) | 6751 (16.2%) |
| **Characteristics during Acute COVID Phase** |  |  |  |  |
| COVID-associated Hospitalization | 3100 (37.3%) | 6165 (14.8%) | 6306 (15.2%) | 6162 (14.8%) |
| COVID-associated ED Visit | 1564 (18.8%) | 6468 (15.5%) | 6060 (14.6%) | 5865 (14.1%) |
| Hospitalization stay (Mean [SD]) | 5.6 (15.3) | 1.1 (6.1) | 1.1 (5.3) | 1.0 (4.8) |
| COVID treatment |  |  |  |  |
| Corticosteroids<sup>a</sup> | 2025 (24.3%) | 3054 (7.3%) | 2991 (7.2%) | 2807 (6.7%) |
| Remdesivir<sup>a</sup> | 1409 (16.9%) | 1913 (4.6%) | 1794 (4.3%) | 1631 (3.9%) |
| Vasopressors<sup>a</sup> | 601 (7.2%) | 682 (1.6%) | 703 (1.7%) | 720 (1.7%) |
| ECMO<sup>a</sup> | 66 (0.8%) | 35 (0.1%) | 24 (0.1%) | <20 |
| Mechanical Ventilation<sup>a</sup> | 615 (7.4%) | 450 (1.1%) | 398 (1.0%) | 404 (1.0%) |
| AKI during COVID-associated Hospitalization | 664 (8.0%) | 1016 (2.4%) | 1084 (2.6%) | 1026 (2.5%) |
| Sepsis during COVID-associated Hospitalization | 614 (7.4%) | 823 (2.0%) | 835 (2.0%) | 770 (1.9%) |

<sup>a</sup>Only captured for individuals hospitalized for COVID-19 <sup>b</sup>The restricted samples (Methods 2 and 3) lose 3 cases due to not having sufficient controls (<5 available controls). Comorbidities shown in this Table are selected. A comprehensive stratification by comorbidities is in the Supplement.

For SDoH, we used county-level variables from the Sharecare-Boston University School of Public Health Social Determinants of Health dataset [26]. Specifically, we used percent of households with income below poverty, percent of residents with college degree, percent of residents 19-64 with public insurance, and physicians per 1000 residents [26]. These are all included as tertiles in the analyses.

### Statistical analysis

We used descriptive statistics to compare PASC cases with the three non-PASC control cohorts, including counts and percentages for categorical variables and means and standard deviation for continuous variables.

We used multivariable logistic regression to determine associations between risk factors and PASC. We constructed three separate logistic regression models for the three cohorts of matched cases and controls. All patient characteristics, with and without SDoH, were included as independent variables in the three models. We reported odds ratios (OR) and 95% confidence intervals (CI) for risk factors.

In addition to logistic regression, we used two machine learning methods, random forest (RF) [27] and XGBoost, to identify influential risk factors for developing PASC [28]. Machine learning methods provide the ability to investigate massive datasets and reveal patterns within data without relying on a priori assumptions such as pre-specified statistical interactions, specific variable associations, or linearity in variable relationships [29]. We conducted feature importance analysis for both RF and XGBoost models [30], and display SHAP (SHapley Additive exPlanations) plots [31] from the XGboost models **(eMethods)**.

### Secondary and stratified analysis

For the unrestricted controls and PASC cases defined by U09.9 or a long-COVID visit (primary cohort), we performed planned secondary analysis by including SDoH variables in logistic regression and two machine learning models. We performed stratified analysis by hospitalization status to assess whether risk factors differed for these two groups **(eMethods)**.

### Sensitivity analyses

To check the robustness of our results, we examined risk factors using the matched case-control design separately for cases identified: (a) using U09.9 diagnosis code and (b) based on long-COVID clinic visits, each with five matched controls. We refit each of the three model types in the above six cohorts of PASC cases and matched controls.

## RESULTS

### Study cohort

Among the 8,325 individuals with PASC, the majority were >50 years of age (56.6%), female (62.8%), and non-Hispanic White (68.6%) **(Table 1)**. The most common comorbidities were obesity (56.4%), hypertension (40.4%), chronic lung disease (28.9%), and uncomplicated diabetes (20.5%). Compared to unrestricted controls (N=41,625), PASC cases were older (mean age 52 [SD 15.5] vs. 46 [SD 17.8] years), and greater proportion were male (37.2% vs. 44.4%) and non-Hispanic White (68.6% vs. 63.6%). The prevalence of all comorbidities was higher among PASC cases compared to controls, such as hypertension (40.4% vs. 26.2%), chronic lung disease (28.9% vs. 13.7%), and uncomplicated diabetes (20.5% vs. 13.3%). The rate of COVID-associated hospitalization was much higher among cases (37.3% vs. 14.8%) compared to all controls. We found similar patterns when comparing PASC cases with the less restrictive and more restrictive control cohorts (**Table 1** and **eTable 1**).

### Risk factors associated with PASC

#### Unrestricted Controls (Primary Analysis)

Using logistic regression **(eFigure 2, eTable 2)** we identified that age was a risk factor for PASC, with particularly high risk among individuals between 40 and 69 years (OR ranging from 2.32 to 2.58). Females had a greater likelihood of having PASC (OR 1.40, CI 1.33-1.48). Non-Hispanic Blacks (OR 0.78, CI 0.73-0.85), Hispanics (OR 0.80, CI 0.73-0.87), and Asians (OR 0.80, CI 0.66-0.97) had a lower likelihood of having PASC than non-Hispanic Whites. The top five comorbidities associated with PASC were tuberculosis (OR 1.65, CI 1.03-2.65), chronic lung disease (OR 1.63, CI 1.53-1.74), rheumatologic disease (OR 1.27, CI 1.11-1.46), peptic ulcer (OR 1.25, CI 1.07-1.46) and obesity (OR 1.23, CI 1.16-1.30). Severe acute infection were the strongest predictors of PASC including extended hospital stays (31+ days, OR 3.38, CI 2.45-4.67), long hospital stays (8-30 days, OR 1.69, CI 1.31-2.17), COVID-associated hospitalizations (OR 3.8, CI 3.05-4.73), and mechanical ventilation (OR 1.44, CI 1.18-1.74). Characteristics associated with a lower likelihood of PASC included psychosis, cardiomyopathies, metastatic cancer, moderate to severe liver disease, substance abuse, tobacco smoking, and COVID-19 diagnosis during hospitalization.

The performance of XGBoost and logistic regression models was similar (both AUC 0.73), closely followed by RF model (AUC 0.69) **(eTable 3)**. Risk factors for PASC identified by the XGBoost models had a similar direction compared to logistic regression models **(Table2, eTable 4)**. However, risk factors’ magnitude and order of importance varied between XGBoost and logistic regression. For example, invasive mechanical ventilation was ranked 6 by XGBoost versus 21 by logistic regression.

**Table 2.** Comparison of Feature Importance for PASC Models defined by U09.9 or long-COVID clinic visit and unrestricted controls (Top 15 positive and negative features) This Table shows the top 15 features associated with increased risk and top 15 features associated with decreased risk. Complete models are shown in the Supplement. Unrestricted sample, U09.9 or long-COVID clinic visit target (see text). Grouped by median direction (increased/decreased) and ordered by mean rank. Model rank calculated based on sklearn.inspection.permutation_importance() (XGB/RF) or absolute ordered size of coefficient (LR). Mean rank is based on the rank of each model that had the variable in the model. Mint color indicates features associated with increased risk. Salmon color indicates features associated with decreased risk. An uncolored cell indicates that that feature was the reference group for the logistic regression model.

| Features | Logistic Regression | XGBoost | Random Forest | Mean Rank |  |
| --- | --- | --- | --- | --- | --- |
| Hospitalization Extended Stay (31+ days) | 2 | 1 | 4 | 2.33 | FEATURES ASSOCIATED WITH INCREASED RISK |
| COVID-associated Hospitalization | 4 | 4 | 1 | 3.00 |  |
| Age 40-49 | 6 | 8 | 7 | 7.00 |  |
| Age 50-59 | 5 | 14 | 6 | 8.33 |  |
| Hospitalization Long Stay (8-30 days) | 8 | 2 | 18 | 9.33 |  |
| Female | 22 | 13 | 2 | 12.33 |  |
| Depression | 19 | 10 | 15 | 14.67 |  |
| Age 60-69 | 7 | 28 | 10 | 15.00 |  |
| COVID Treatment: Mechanical Ventilation | 21 | 5 | 24 | 16.67 |  |
| COVID Treatment: Remdesivir | 26 | 6 | 20 | 17.33 |  |
| Chronic Lung Disease | 16 | 38 | 9 | 21.00 |  |
| COVID-associated ED Visit | 36 | 7 | 25 | 22.67 |  |
| Obesity | 37 | 15 | 27 | 26.33 |  |
| Systemic Corticosteroids | 24 | 34 | 22 | 26.67 |  |
| Malignant Cancer | 54 | 21 | 12 | 29.00 |  |
| COVID Diagnosis during COVID-associated Hospitalization | 11 | 3 | 5 | 6.33 | FEATURES ASSOC. WITH DECREASED RISK |
| Age 18-29 | ref. | 9 | 11 | 10.00 |  |
| Male | ref. | 19 | 3 | 11.00 |  |
| Age 70-79 | 10 | 35 | 16 | 20.33 |  |
| Substance Abuse | 14 | 16 | 37 | 22.33 |  |
| Tobacco Smoker | 23 | 11 | 34 | 22.67 |  |
| Cardiomyopathies | 32 | 22 | 17 | 23.67 |  |
| Age 30-39 | 12 | 53 | 8 | 24.33 |  |
| Metastatic Solid Tumor Cancers | 13 | 18 | 45 | 25.33 |  |
| Psychosis | 9 | 23 | 44 | 25.33 |  |
| Uncomplicated Diabetes | 45 | 20 | 13 | 26.00 |  |
| Myocardial Infarction | 33 | 32 | 19 | 28.00 |  |
| Age 80-89 | 20 | 12 | 56 | 29.33 |  |
| Dementia | 38 | 29 | 26 | 31.00 |  |
| Race/Ethnicity: Black NH | 28 | 42 | 29 | 33.00 |  |

#### Restricted Controls

**<u>eTable 5</u>** <u>and</u> **<u>eTable 6</u>** <u>shows the importance of risk factors</u> among less restrictive and more restrictive controls, respectively. For most patient characteristics, the direction and magnitude of the odds ratios were similar to the primary analysis **(eTable 2)**. However, obesity was no longer significant when we used the less and more restrictive controls. Also, ECMO was associated with PASC when the more restrictive controls were used, but it was not a statistically significant factor when the unrestricted controls were used.

### Secondary Analysis Including SDoH

We repeated our primary analysis (U09.9 or long-COVID clinic model, unrestricted control cohort) by adding SdoH variables**(Figure 2, eTable 7)**. The number of medical doctors per 1000 residents in the county of residence was associated with PASC, indicating having access to healthcare services increases the likelihood of diagnosis and/or treatment at a long-COVID clinic. Other SDoH factors were not associated with PASC in logistic regression but were important features in the machine learning models (**eFigure 3, Table 3**).

**Figure 2.**
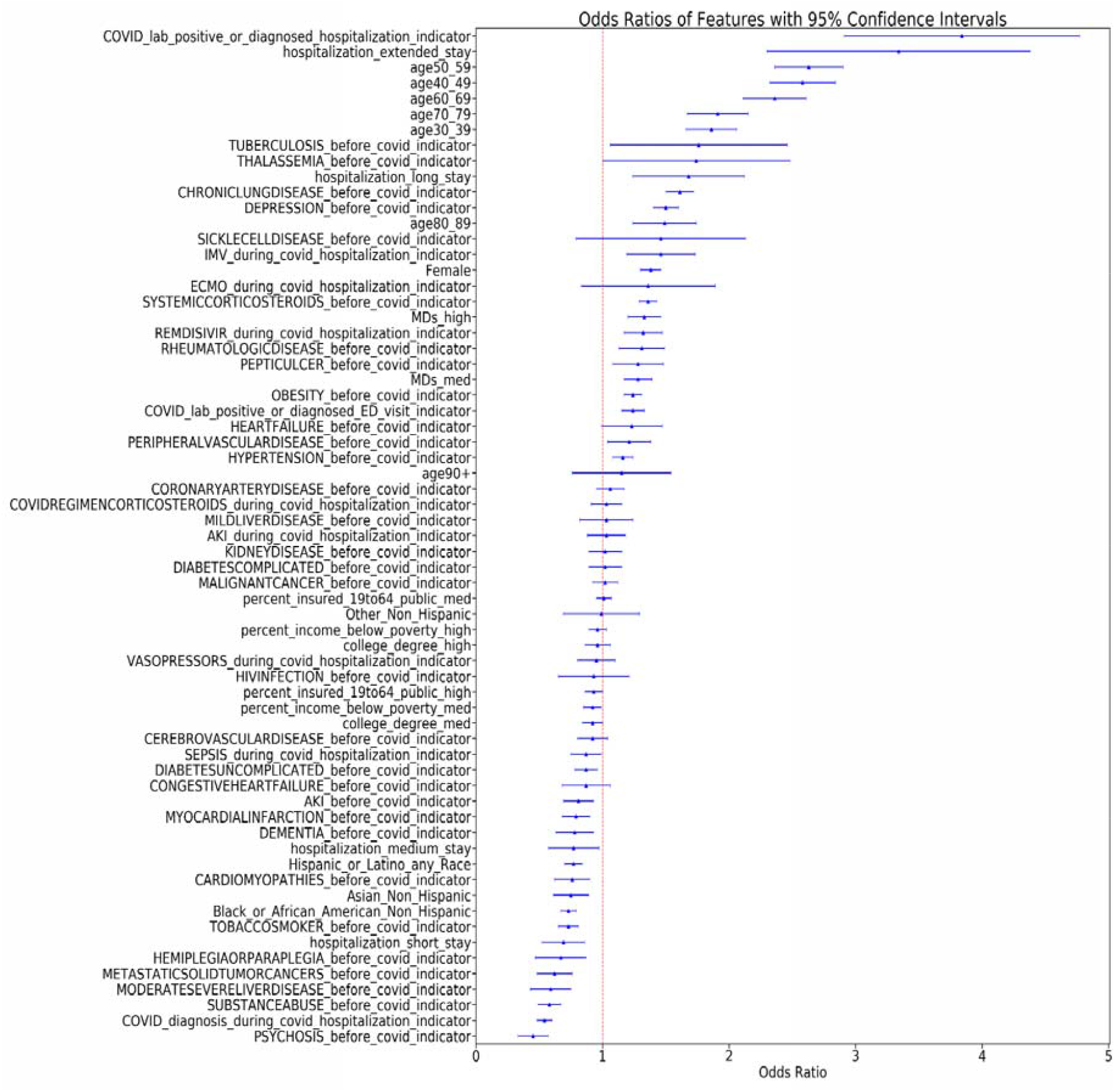
Forest Plots from Logistic Regression for Unrestricted Controls with SDoH (PASC defined as U09.9 or long-COVID Clinic Visit)

**Table 3.** Comparison of Feature Importance for PASC Models defined by U09.9 or long-COVID clinic visit and unrestricted controls (Top 15 positive and negative features) with SDoH variables included. This Table shows the Top 15 features associated with increased risk and top 15 features associated with decreased risk. Complete models are shown in the Supplement. Not restricted sample, U09.9 or long-COVID clinic visit target (see text). Grouped by median direction (increased/decreased) and ordered by mean rank. Model rank calculated based on sklearn.inspection.permutation_importance() (XGB/RF) or absolute ordered size of coefficient (LR). Mean rank is based on the rank of each model that had the variable in the model. Mint color indicates features associated with increased risk. Salmon color indicates features associated with decreased risk. An uncolored cell indicates that that feature was the reference group for the logistic regression model.

| features | Logistic Regression SDOH | XGBoost SDOH | Random Forest SDOH | Mean Rank |  |
| --- | --- | --- | --- | --- | --- |
| Hospitalization Extended Stay (31+ days) | 2 | 1 | 19 | 7.33 | FEATURES ASSOCIATED WITH INCREASED RISK |
| COVID-associated Hospitalization | 4 | 2 | 22 | 9.33 |  |
| Age 40-49 | 6 | 16 | 8 | 10.00 |  |
| Age 50-59 | 5 | 11 | 15 | 10.33 |  |
| Hospitalization Long Stay (8-30 days) | 8 | 4 | 23 | 11.67 |  |
| Households with Income below poverty: low (<11%) | ref. | 19 | 5 | 12.00 |  |
| MDs per 1000 residents: High (>3.61%) | 30 | 8 | 6 | 14.67 |  |
| COVID Treatment: Mechanical Ventilation | 22 | 5 | 27 | 18.00 |  |
| Depression | 19 | 17 | 31 | 22.33 |  |
| Female | 24 | 42 | 1 | 22.33 |  |
| Age 60-69 | 7 | 49 | 17 | 24.33 |  |
| MDs per 1000 residents: medium (1.91-3.61%) | 36 | 34 | 4 | 24.67 |  |
| Obesity | 39 | 13 | 30 | 27.33 |  |
| Chronic Lung Disease | 17 | 53 | 16 | 28.67 |  |
| COVID-associated ED Visit | 42 | 9 | 40 | 30.33 |  |
| MDs per 1000 residents: Low (<1.91%) | ref. | 7 | 10 | 8.50 | FEATURES ASSOCIATED WITH DECREASED RISK |
| College Degree low (<19%) | ref. | 18 | 7 | 12.50 |  |
| COVID Diagnosis during COVID-associated Hospitalization | 11 | 3 | 26 | 13.33 |  |
| Age 18-29 | ref. | 10 | 20 | 15.00 |  |
| Male | ref. | 33 | 2 | 17.50 |  |
| Age 30-39 | 12 | 38 | 11 | 20.33 |  |
| College Degree medium (19-25%) | 50 | 12 | 3 | 21.67 |  |
| Public health Insurance for ages 19-64: Low (<13%) | ref. | 31 | 13 | 22.00 |  |
| Substance Abuse | 15 | 15 | 38 | 22.67 |  |
| Psychosis | 9 | 26 | 45 | 26.67 |  |
| Tobacco Smoker | 26 | 14 | 40 | 26.67 |  |
| Age 80-89 | 20 | 25 | 43 | 29.33 |  |
| Households with Income below poverty: high (>15%) | 57 | 21 | 12 | 30.00 |  |
| Metastatic Solid Tumor Cancers | 18 | 24 | 51 | 31.00 |  |
| Age 70-79 | 10 | 60 | 24 | 31.33 |  |

### Stratified Analysis by COVID-index Hospitalization

To assess risk factors unique to less severe SARS-CoV-2 infections, we stratified analysis by whether the patient was hospitalized at the time of COVID-19 index date (**eTables 8-13)**. For the hospitalized sample, the strongest risk factors across LR, XGBoost, and RF models are possible markers of COVID-19 severity (e.g., ECMO, ED Visit, Mechanical Ventilation) and obesity. Living in a community with higher education increased likelihood of diagnosis or care at a long-COVID clinic (**eFigure 4**). For those not hospitalized at COVID index date, the following risk factors pre-COVID differ from hospitalized patients: systemic corticosteroid use and depression, peptic ulcer, or coronary artery disease diagnosis. When we limit to non-hospitalized patients during COVID-19 index, some SDoH factors were also strong predictors including lower poverty and higher education communities (**eFigure 4, eFigure 5**). Some risk factors are common to both the hospitalized and non-hospitalized samples, including middle age (40-69), chronic lung disease, and white non-Hispanic race/ethnicity (**eFigure 4, eFigure 5**).

### Sensitivity Analysis: Other Definitions of PASC

We have described sensitivity analysis in detail in **eResults**. Overall, sensitivity analysis results based on only U09.9 definition or only long-COVID clinic visits were similar to the primary analysis.

## DISCUSSION

In this first large-scale US study of PASC risk factors, we found that middle age (40 to 69 years), female sex, severity of acute infection (e.g., hospitalization for COVID-19, long or extended hospital stay, treatment for acute COVID-19 during hospitalization), and several comorbidities including depression, chronic lung disease, obesity, and malignant cancer were associated with increased likelihood of PASC diagnosis or care at a long-COVID clinic. Risk factors associated with a lower likelihood of PASC diagnosis or care at a long-COVID clinic included younger age (18 to 29 years), male sex, non-Hispanic Black race, and comorbidities such as substance abuse, cardiomyopathy, psychosis, and dementia. We also found that a greater number of physicians per capita in the county of residence were associated with an increased likelihood of PASC diagnosis or care. Our findings were consistent in sensitivity analyses using a variety of approaches to select controls and several robust analytic techniques.

Our findings add to the growing body of evidence identifying and characterizing PASC risk factors. Although females were less likely to die or be hospitalized due to acute COVID-19, [32,33], they appear to have a greater risk of developing PASC. Our finding that there is a higher likelihood of PASC diagnosis among middle-aged individuals is consistent with a recent United Kingdom Office for National Statistics analysis, but is in contrast with another report that found that older individuals were at the highest risk for PASC[8,12]. Risk factors such as chronic lung disease, rheumatologic disease, and obesity were associated with both hospitalization and death due to COVID-19 and also increased risk of PASC diagnosis or care.

We previously established a machine learning phenotype [23] that used clinical features observed *after* COVID-19 infection to generate a probability for whether a patient *currently has* PASC. In contrast, the current analysis uses features selected from the acute phase of COVID-19 (such as pre-existing clinical comorbidities and hospitalization characteristics at the time of the initial infection) to assess risk factors for the later emergence of PASC as indicated by a U09.9 diagnosis or long-COVID clinic visit. The models in this analysis can be applied by clinicians to identify patients at risk for PASC while they are still in the acute phase of their infection and also to support targeted enrollment in clinical trials for preventing or treating PASC.

The association we found between more severe acute COVID-19 and increased likelihood of PASC is consistent with prior literature [34]. Individuals who were hospitalized for COVID-19 or received intensive treatment may have long-lasting effects on the brain, heart, lungs, and other organs [35–39]. Counterintuitively, we found that diabetes, a strong risk factor for worse outcomes after acute COVID-19, was associated with less likelihood of PASC diagnosis. Our previous work has demonstrated that glycemic control in patients with diabetes, as measured by pre-infection HbA1c levels, is an important risk factor for poor acute infection outcomes[40]. The level of granularity available in EHR data may not be sufficient to completely disentangle PASC risk associated with some comorbidities from PASC risk from SDoH and unmeasured biological features. We found that a pre-existing diagnosis of depression was associated with a higher risk of subsequent PASC. Interestingly, however, prior diagnoses of other mental health diagnoses (e.g., psychosis) were associated with lower risk. Comorbid substance abuse (also associated with lower likelihood of PASC diagnosis) with psychosis may explain some of this difference, as those with substance abuse disorders may have challenges accessing health care. Antidepressants and antipsychotics have differential immunomodulatory effects, which could also contribute to this observation. Another interesting finding is that we found patients with comorbidities such as cardiomyopathy, metastatic solid tumors, and liver disease that made them vulnerable to worse outcomes after acute COVID-19 had lower likelihood of PASC diagnosis. Although we cannot determine causality from this association, this finding may be hypothesis-generating.

The association we found between higher numbers of doctors per capita with PASC diagnosis or care underscores the importance of access to medical care. Given the disruption of medical care for both COVID and non-COVID illnesses during the pandemic, it is important to improve access to care, particularly for minorities [41]. Our findings of lower likelihood of PASC diagnosis among non-Hispanic Blacks support this hypothesis. The focus of this study was to investigate patient-level factors and therefore we did not consider several SDoH that can impact PASC risk such as essential worker status, financial issues, housing, and isolation. These are excellent candidate variables for future study[42]. Future research is also required to delineate the complex relationship of individual vs. contextual factors in the diagnosis and care for PASC. Policy measures such as strengthening primary care, optimizing SDoH data quality, and addressing SDoH are required to reduce inequalities in diagnosis and care for PASC [17].

The US Government Accountability Office estimates that between 7.7 and 23 million US adults have PASC [43]. Given the potential clinical and economic consequences, the US government has allocated over a billion dollars to study it[44]. Our study validates some findings of prior studies on PASC risk factors and provides novel information including the impact of SDoH. With the sample size available in N3C, we can evaluate more risk factors simultaneously than previous studies. Also, this study can be used to generate hypotheses about possible mechanisms and potential treatments for PASC. For example, because this study found that rheumatological conditions are a risk factor for PASC, future studies can assess whether treatment for rheumatological conditions can alter the likelihood of PASC diagnosis.

Our study has several limitations. First, the N3C only contains EHR data, which has inherent limitations and may encode biases related to health care access and racism. N3C collects data from health systems that maintain a data warehouse using one of four common data models (OMOP, PCORnet, ACT, and TriNetX) [20]. However, the age, sex, race, and ethnicity distribution in N3C is representative of many segments of the U.S. population. Therefore, our findings on risk factors may generalize to the broader US population. Second, because identification of individuals without PASC (controls) is not straightforward without clear definitions or biomarkers, we used three approaches to identify controls. Two of those leveraged our CP classification model for long-COVID [23]. Some pre-existing conditions can carry forward from the acute phase and appear later as features in the PASC phase. We acknowledge some potential for circularity. Importantly, however, model performance did not have clinically meaningful differences across different cohort selection methods. Third, further analysis is needed to determine the role of SDoH and how it impacts individual-level risk factors for PASC. While research shows that county-level SDoH variables can be significant for patient-level analysis, more granular geographic unit or patient-level data would likely provide a greater understanding of the relationship between SDoH and PASC outcomes [45,46]. Fourth, we did not evaluate the role of vaccines and therapeutics such as paxlovid for the likelihood of PASC diagnosis. Fifth, we did not evaluate the association of COVID-19 reinfection and PASC diagnosis or care.

## CONCLUSIONS

This national study using N3C data identified important risk factors for PASC such as middle age, severe COVID-19 disease, and comorbidities. Further clinical and epidemiological research is needed to better understand underlying mechanisms and the potential role of vaccines and therapeutics in altering the course of PASC.

## Data Availability

All data produced in the present study are available upon reasonable request to the authors

## Acknowledgements

This work was supported by the National Institutes of Health awards as follows: CD2H NCATS U24 TR002306, NHLBI RECOVER Agreement OT2HL161847-01; and was conducted under the N3C DUR RP-5677B5. The views and conclusions contained in this document are those of the authors and should not be interpreted as representing the official policies, either expressed or implied, of the NIH.

This research was possible because of the patients whose information is included within the data from participating organizations (covid.cd2h.org/dtas) and the organizations and scientists (covid.cd2h.org/duas) who have contributed to the on-going development of this community resource.

The analyses described in this publication were conducted with data or tools accessed through the NCATS N3C Data Enclave covid.cd2h.org/enclave and supported by NCATS U24 TR002306. This research was possible because of the patients whose information is included within the data from participating organizations (covid.cd2h.org/dtas) and the organizations and scientists (covid.cd2h.org/duas) who have contributed to the on-going development of this community resource.[20]

The N3C data transfer to NCATS is performed under a Johns Hopkins University Reliance Protocol # IRB00249128 or individual site agreements with NIH. The N3C Data Enclave is managed under the authority of the NIH; information can be found at https://ncats.nih.gov/n3c/resources.

We also thank Dr. Nina Cesare and colleagues at the Boston University School of Public Health for access to the public dataset provided was created under the support of Sharecare through a partnership with Boston University’s School of Public Health.

Authorship was determined using ICMJE recommendations. The authors gratefully acknowledge Evan Volkin for his excellent research assistance. We also acknowledge the following core contributors to N3C: Anita Walden, Leonie Misquitta, Joni L. Rutter, Kenneth R. Gersing, Penny Wung Burgoon, Samuel Bozzette, Mariam Deacy, Christopher Dillon, Rebecca Erwin-Cohen, Nicole Garbarini, Valery Gordon, Michael G. Kurilla, Emily Carlson Marti, Sam G. Michael, Lili Portilla, Clare Schmitt, Meredith Temple-O’Connor, David A. Eichmann, Warren A. Kibbe, Hongfang Liu, Philip R.O. Payne, Emily R. Pfaff, Peter N. Robinson, Joel H. Saltz, Heidi Spratt, Justin Starren, Christine Suver, Adam B. Wilcox, Andrew E. Williams, Chunlei Wu, Davera Gabriel, Stephanie S. Hong, Kristin Kostka, Harold P. Lehmann, Michele Morris, Matvey B. Palchuk, Xiaohan Tanner Zhang, Richard L. Zhu, Benjamin Amor, Mark M. Bissell, Marshall Clark, Andrew T. Girvin, Stephanie S. Hong, Kristin Kostka, Adam M. Lee, Robert T. Miller, Michele Morris, Matvey B. Palchuk, Kellie M. Walters, Will Cooper, Patricia A. Francis, Rafael Fuentes, Alexis Graves, Julie A. McMurry, Shawn T. O’Neil, Usman Sheikh, Elizabeth Zampino, Katie Rebecca Bradwell, Andrew T. Girvin, Amin Manna, Nabeel Qureshi, Christine Suver, Julie A. McMurry, Carolyn Bramante, Jeremy Richard Harper, Wenndy Hernandez, Farrukh M Koraishy, Amit Saha, Satyanarayana Vedula, Johanna Loomba, Andrea Zhou, Steve Johnson, Evan French, Alfred (Jerrod) Anzalone, Umit Topaloglu, Amy Olex, Hythem Sidkey. Details of core contributions available at covid.cd2h.org/acknowledgements

We acknowledge the following institutions whose data is released or pending:

## Available

Advocate Health Care Network — UL1TR002389: The Institute for Translational Medicine (ITM) • Boston University Medical Campus — UL1TR001430: Boston University Clinical and Translational Science Institute • Brown University — U54GM115677: Advance Clinical Translational Research (Advance-CTR) • Carilion Clinic — UL1TR003015: iTHRIV Integrated Translational health Research Institute of Virginia • Charleston Area Medical Center — U54GM104942: West Virginia Clinical and Translational Science Institute (WVCTSI) • Children’s Hospital Colorado — UL1TR002535: Colorado Clinical and Translational Sciences Institute • Columbia University Irving Medical Center — UL1TR001873: Irving Institute for Clinical and Translational Research • Duke University — UL1TR002553: Duke Clinical and Translational Science Institute • George Washington Children’s Research Institute — UL1TR001876: Clinical and Translational Science Institute at Children’s National (CTSA-CN) • George Washington University — UL1TR001876: Clinical and Translational Science Institute at Children’s National (CTSA-CN) • Indiana University School of Medicine — UL1TR002529: Indiana Clinical and Translational Science Institute • Johns Hopkins University — UL1TR003098: Johns Hopkins Institute for Clinical and Translational Research • Loyola Medicine — Loyola University Medical Center Loyola University Medical Center — UL1TR002389: The Institute for Translational Medicine (ITM) • Maine Medical Center — U54GM115516: Northern New England Clinical & Translational Research (NNE-CTR) Network • Massachusetts General Brigham — UL1TR002541: Harvard Catalyst • Mayo Clinic Rochester — UL1TR002377: Mayo Clinic Center for Clinical and Translational Science (CCaTS) • Medical University of South Carolina — UL1TR001450: South Carolina Clinical & Translational Research Institute (SCTR) • Montefiore Medical Center — UL1TR002556: Institute for Clinical and Translational Research at Einstein and Montefiore • Nemours — U54GM104941: Delaware CTR ACCEL Program • NorthShore University HealthSystem — UL1TR002389: The Institute for Translational Medicine (ITM) • Northwestern University at Chicago — UL1TR001422: Northwestern University Clinical and Translational Science Institute (NUCATS) • OCHIN — INV-018455: Bill and Melinda Gates Foundation grant to Sage Bionetworks • Oregon Health & Science University — UL1TR002369: Oregon Clinical and Translational Research Institute • Penn State Health Milton S. Hershey Medical Center — UL1TR002014: Penn State Clinical and Translational Science Institute • Rush University Medical Center UL1TR002389: The Institute for Translational Medicine (ITM) • Rutgers, The State University of New Jersey — UL1TR003017: New Jersey Alliance for Clinical and Translational Science • Stony Brook University — U24TR002306 • The Ohio State University — UL1TR002733: Center for Clinical and Translational Science • The State University of New York at Buffalo — UL1TR001412: Clinical and Translational Science Institute • The University of Chicago — UL1TR002389: The Institute for Translational Medicine (ITM) • The University of Iowa — UL1TR002537: Institute for Clinical and Translational Science • The University of Miami Leonard M. Miller School of Medicine — UL1TR002736: University of Miami Clinical and Translational Science Institute • The University of Michigan at Ann Arbor — UL1TR002240: Michigan Institute for Clinical and Health Research • The University of Texas Health Science Center at Houston — UL1TR003167: Center for Clinical and Translational Sciences (CCTS) • The University of Texas Medical Branch at Galveston — UL1TR001439: The Institute for Translational Sciences • The University of Utah — UL1TR002538: Uhealth Center for Clinical and Translational Science • Tufts Medical Center — UL1TR002544: Tufts Clinical and Translational Science Institute • Tulane University — UL1TR003096: Center for Clinical and Translational Science • University Medical Center New Orleans — U54GM104940: Louisiana Clinical and Translational Science (LA CaTS) Center • University of Alabama at Birmingham — UL1TR003096: Center for Clinical and Translational Science • University of Arkansas for Medical Sciences — UL1TR003107: UAMS Translational Research Institute • University of Cincinnati — UL1TR001425: Center for Clinical and Translational Science and Training University of Colorado Denver, Anschutz Medical Campus — UL1TR002535: Colorado Clinical and Translational Sciences Institute • University of Illinois at Chicago — UL1TR002003: UIC Center for Clinical and Translational Science • University of Kansas Medical Center — UL1TR002366: Frontiers: University of Kansas Clinical and Translational Science Institute • University of Kentucky — UL1TR001998: UK Center for Clinical and Translational Science • University of Massachusetts Medical School Worcester — UL1TR001453: The UMass Center for Clinical and Translational Science (UMCCTS) • University of Minnesota — UL1TR002494: Clinical and Translational Science Institute • University of Mississippi Medical Center — U54GM115428: Mississippi Center for Clinical and Translational Research (CCTR) • University of Nebraska Medical Center — U54GM115458: Great Plains IDeA-Clinical & Translational Research • University of North Carolina at Chapel Hill — UL1TR002489: North Carolina Translational and Clinical Science Institute • University of Oklahoma Health Sciences Center — U54GM104938: Oklahoma Clinical and Translational Science Institute (OCTSI) • University of Rochester — UL1TR002001: UR Clinical & Translational Science Institute • University of Southern California — UL1TR001855: The Southern California Clinical and Translational Science Institute (SC CTSI) • University of Vermont U54GM115516: Northern New England Clinical & Translational Research (NNE-CTR) Network • University of Virginia — UL1TR003015: iTHRIV Integrated Translational health Research Institute of Virginia • University of Washington — UL1TR002319: Institute of Translational Health Sciences • University of Wisconsin-Madison — UL1TR002373: UW Institute for Clinical and Translational Research • Vanderbilt University Medical Center — UL1TR002243: Vanderbilt Institute for Clinical and Translational Research • Virginia Commonwealth University — UL1TR002649: C. Kenneth and Dianne Wright Center for Clinical and Translational Research • Wake Forest University Health Sciences — UL1TR001420: Wake Forest Clinical and Translational Science Institute • Washington University in St. Louis — UL1TR002345: Institute of Clinical and Translational Sciences • Weill Medical College of Cornell University — UL1TR002384: Weill Cornell Medicine Clinical and Translational Science Center • West Virginia University — U54GM104942: West Virginia Clinical and Translational Science Institute (WVCTSI)

## Submitted

Icahn School of Medicine at Mount Sinai — UL1TR001433: ConduITS Institute for Translational Sciences • The University of Texas Health Science Center at Tyler — UL1TR003167: Center for Clinical and Translational Sciences (CCTS) • University of California, Davis — UL1TR001860: UCDavis Health Clinical and Translational Science Center • University of California, Irvine — UL1TR001414: The UC Irvine Institute for Clinical and Translational Science (ICTS) • University of California, Los Angeles — UL1TR001881: UCLA Clinical Translational Science Institute • University of California, San Diego — UL1TR001442: Altman Clinical and Translational Research Institute • University of California, San Francisco — UL1TR001872: UCSF Clinical and Translational Science Institute

## Pending

Arkansas Children’s Hospital — UL1TR003107: UAMS Translational Research Institute • Baylor College of Medicine — None (Voluntary) • Children’s Hospital of Philadelphia — UL1TR001878: Institute for Translational Medicine and Therapeutics • Cincinnati Children’s Hospital Medical Center — UL1TR001425: Center for Clinical and Translational Science and Training • Emory University — UL1TR002378: Georgia Clinical and Translational Science Alliance • HonorHealth — None (Voluntary) • Loyola University Chicago — UL1TR002389: The Institute for Translational Medicine (ITM) • Medical College of Wisconsin — UL1TR001436: Clinical and Translational Science Institute of Southeast Wisconsin • MedStar Health Research Institute — UL1TR001409: The Georgetown-Howard Universities Center for Clinical and Translational Science (GHUCCTS) • MetroHealth — None (Voluntary) • Montana State University — U54GM115371: American Indian/Alaska Native CTR • NYU Langone Medical Center UL1TR001445: Langone Health’s Clinical and Translational Science Institute • Ochsner Medical Center — U54GM104940: Louisiana Clinical and Translational Science (LA CaTS) Center • Regenstrief Institute — UL1TR002529: Indiana Clinical and Translational Science Institute • Sanford Research — None (Voluntary) • Stanford University — UL1TR003142: Spectrum: The Stanford Center for Clinical and Translational Research and Education • The Rockefeller University — UL1TR001866: Center for Clinical and Translational Science • The Scripps Research Institute — UL1TR002550: Scripps Research Translational Institute • University of Florida — UL1TR001427: UF Clinical and Translational Science Institute • University of New Mexico Health Sciences Center — UL1TR001449: University of New Mexico Clinical and Translational Science Center • University of Texas Health Science Center at San Antonio — UL1TR002645: Institute for Integration of Medicine and Science • Yale New Haven Hospital — UL1TR001863: Yale Center for Clinical Investigation

