## Supplemental Material for "Risk Factors Associated with Post-Acute Sequelae of SARS-CoV-2 in an EHR Cohort: A National COVID Cohort Collaborative (N3C) Analysis as part of the NIH RECOVER program"

**eMethods**

**eResults**

**eFigures**

eFigure 1. Feature Importance XGBoost (PASC defined as U09.9 or Long-COVID Clinic Visit)

eFigure 2. Forest Plot for Unrestricted Controls without SDoH (PASC defined as U09.9 or Long-COVID Clinic Visit)

eFigure 3. Feature Importance XGBoost for Unrestricted Controls with SDoH (PASC defined as U09.9 or Long-COVID Clinic Visit)

eFigure 4. Feature Importance XGBoost for Unrestricted Controls with SDoH–Hospitalized Sample (PASC defined as U09.9 or Long COVID Visit)

eFigure 5. Feature Importance XGBoost for Unrestricted Controls with SDoH –non-Hospitalized Sample (PASC defined as U09.9 or Long COVID Visit)

**eTables**

eTable 1. Cohort Characteristics for PASC cases defined by U09.9 or clinic visit (Additional Characteristics Not Shown in Table 1)

eTable 2. PASC Risk Factors from Logistic Regression (PASC defined as U09.9 or long-COVID Clinic Visit)

eTable 3. AUC scores for our three models of PASC defined by U09.9 or Long-COVID Clinic Visit

eTable 4. Comparison of Feature Importance Across Models for Unrestricted Sample (PASC defined as U09.9 or Long-COVID Clinic Visit)

eTable 5. Comparison of Feature Importance Across Models for Restricted Sample (PASC defined as U09.9 or Long-COVID Clinic Visit)

eTable 6. Comparison of Feature Importance Across Models for More Restricted Sample (PASC defined as U09.9 or Long-COVID Clinic Visit)

eTable 7. Comparison of Feature Importance Across Models for Unrestricted Sample including SDOH variables (PASC defined as U09.9 or Long-COVID Clinic Visit)

eTable 8. Cohort Characteristics for PASC cases defined by U09.9 or clinic visit for Hospitalized During COVID Index

eTable 9. Cohort Characteristics for PASC cases defined by U09.9 or clinic visit for Not Hospitalized During COVID Index11

eTable 10. PASC Risk Factors from Logistic Regression (PASC defined as U09.9 or long-COVID visit) Hospitalized during COVID Index

eTable 11. PASC Risk Factors from Logistic Regression including SDoH (PASC defined as U09.9 or long-COVID visit) Not Hospitalized during COVID Index

eTable 12. Comparison of Feature Importance Across Models for Hospitalized during Index COVID for Unrestricted Sample including SDOH variables (PASC defined as U09.9 or Long-COVID Clinic Visit)

eTable 13. Comparison of Feature Importance Across Models for Not-Hospitalized during Index COVID for Unrestricted Sample including SDOH variables

eTable 14. Characteristics of Cohorts for U09.9 only

eTable 15. PASC Risk Factors from Logistic Regression (PASC defined as U09.9)

eTable 16. Characteristics of Cohorts for Long-COVID Clinic Visits

eTable 17. PASC Risk Factors from Logistic Regression (PASC defined as Long-COVID Clinic Visits)

eTable 18. Comparison of U09.9 and Long-COVID Clinic Visit Cohorts

**eMethods**

**Data**

We used the National COVID Cohort Collaborative (N3C) data in this study. It has been used extensively to study risk factors, medication use, and long-term health consequences of COVID-19 [[23–26]](https://paperpile.com/c/npl7jJ/aQek+Gkdh+8BSa+dHNl).

We used a limited version of N3C data. Data from numerical lab values were harmonized into canonical units of measure as previously described [28]. Site-level quality filters were applied prior to analysis in order to remove 13 of the 72 data partners with systematic missingness according to the following criteria: sites (1) should not shift dates by more than 30 days, (2) should have serum creatinine and lymphocyte count results for at least 25% of hospitalizations, and (3) should not have >10% of their hospitalizations where the COVID-19 index date is more than 200 days after the visit start date.

The N3C data transfer to NCATS is performed under Johns Hopkins University Reliance Protocol #IRB00249128 or individual site agreements with NIH. The N3C Data Enclave is managed under the authority of the NIH; information can be found at <https://ncats.nih.gov/n3c/resources>.

**Case and control selection**

For two control cohorts, we applied our previously developed computable phenotype (CP) model for long-COVID to refine our control patient pool [[31]](https://paperpile.com/c/npl7jJ/p0Jt). Our group has previously developed two distinct CPs: one for U09.9 diagnosis and another for long-COVID clinic visits. Both CP models have moderate discrimination in classifying long-COVID (correctly categorizing patients as long-COVID cases versus controls based on patient symptoms several months after their infection) and are well-calibrated. We applied CP model to the 1,054,336 non-cases (1,062,661 - 8,325) to generate a predicted probability for U09.9 diagnosis or long-COVID clinic visit. Importantly, to leverage the CP models for refining our control cases while minimizing circularity, we retrained that model without the 4 pre-existing chronic conditions it had previously used (end-stage renal disease, obstructive sleep apnea, multiple sclerosis, and diabetes mellitus types 1 and 2).

In each of the three matching methods, we randomly matched 1 case to 5 controls from the same health system without replacement. To achieve temporal alignment during different pandemic phases, we also matched on COVID index date within +/- 45 days of the corresponding case's earliest COVID index date. In the “unrestricted” method, we matched 8,325 cases to 41,625 controls. In the “restricted” and “more restricted controls” methods, we matched 8,322 cases to 41,610 controls; we lost 3 cases because they did not match 5 controls.

**Risk factors**

To identify comorbidities in N3C, we created concept sets using the primary conditions listed in the Systematized Nomenclature of Medicine – Clinical Terms (SNOMED-CT) hierarchy and included all descendants of the key related concepts (concept sets are reproducible in the N3C Enclave and can be surfaced externally in GitHub). Because this analysis is focused on risk factors, we only included reported instances of these comorbidities that were recorded any time prior to or on the COVID-19 index date.

It is challenging to determine if an individual was hospitalized for or with SARS-CoV-2 infection. To address this, we applied the following criteria to flag an inpatient stay as being associated with COVID-19: the patient was hospitalized +/-14 days from their (a) first positive SARS-CoV-2 PCR or AG lab test OR (b) the first COVID-19 diagnosis (U07.1) that was charted during an inpatient stay or emergency room visit. For individuals hospitalized within +/- 14 days of their index date for COVID-19 infection, we used information from the acute hospitalized phase, i.e, between COVID-19 hospitalization admission and discharge date, and identified invasive mechanical ventilation (IMV) use, extracorporeal membrane oxygenation (ECMO) use, vasopressor use, acute kidney injury diagnosis, sepsis diagnosis, remdesivir use, and total length of hospital stay. The set of comorbidities recorded up to and at the time of infection, the events during the COVID-19-associated hospitalization, and basic patient demographics were included as risk factors in all analyses.

For SDoH, we used county-level variables from the Sharecare-Boston University School of Public Health Social Determinants of Health dataset [34] linked to patients based on the preferred county (majority residence) associated with the patient’s 5-digit ZIP code. Because joining these data requires a five digit zip code, these fields are only available for patients with a five digit zip code. This dataset provides measures across domains identified by the CDC Healthy People 2020 framework: economic stability, education, health and healthcare. We operationalized these variables as: percent of households with income below poverty, percent of residents with college degree, percent of residents 19-64 with public insurance, and physicians per 1000 residents [34]. These are all included as tertiles in the analyses.

**Statistical analysis**

In addition to logistic regression, we used two machine learning methods, random forest (RF) [35] and XGBoost, to identify influential risk factors for developing PASC [36]. Machine learning methods provide the ability to investigate massive datasets and reveal patterns within data without relying on a priori assumptions such as pre-specified statistical interactions, specific variable associations, or linearity in variable relationships [37]. Each machine learning model used 5-fold cross-validation (K = 5) repeated 5 times to decrease overfitting. We used the average area under the receiver operator curve (AUC) as the primary metric to evaluate model performance. Model performance was important to establish as it relates to the effectiveness of the models in their ranking of risk factors associated with the outcome (PASC). We also conducted feature importance analysis for both RF and XGBoost models [38]. See the Supplement for hyperparameter tuning information. We calculated the average of the feature importance scores across the folds to determine which PASC risk factors were most influential. We also display feature importance and estimated SHAP (SHapley Additive exPlanations) plots [39] from the XGboost models (SHAP plots not shown, underlying data shown in Table 2).

For the unrestricted controls and PASC cases defined by U09.9 or a long-COVID visit (primary cohort), we stratified LR, XGBoost, and RF models by patients who were and were not hospitalized at the time of acute COVID infection to assess whether risk factors differed for these two groups.

Hyperparameter Tuning of Machine Learning Models: As XGBoost and random forest models consist of many decision trees, we fine-tuned the tree-specific hyperparameters with ensemble hyperparameters. After multiple iterations, we found our best performing XGBoost classifier model with the following hyperparameters: colsample_bytree=0.1, gamma=0.4, learning_rate=0.09, max_depth=8, min_child_weight=0, n_estimators=400, subsample=0.9. For the random forest classifier, the best performing model had the following hyperparameters: n_estimators=1000, verbose=1, criterion='entropy’. In both cases random_state was set to 42.

**Secondary and stratified analysis**

In the primary cohort (PASC cases defined by U09.9 and a long-COVID visit and unrestricted controls), we then included SDoH variables and refit each of the three model types. These comparisons were important because SDoH have been associated with racial/ethnic disparities in COVID incidence, COVID severity, and development of PASC[19,40,41].

**eResults**

**Sensitivity Analysis: Other Definitions of PASC**

Cases defined as U09.9 When we conducted a sensitivity analysis using only the 7,512 cases identified using ICD10 code U09.9, the magnitudes of PASC risk factors were largely similar to those observed in the primary analysis (eTable 14 and eTable 15). For example, patients between 40 and 69 years, female patients, and patients with comorbidities such as chronic lung disease, rheumatologic disease, and peptic ulcer had higher likelihood of PASC diagnosis. Risk factors negatively associated with PASC include Non-Hispanic Black race and Hispanic ethnicity and behavioral risk factors. As before, the XGBoost indicated similar feature importance and direction of the associations (eFigure 1 Panel B).

Cases defined by long-COVID clinic Visit We then conducted a sensitivity analysis using only data from the 5 health systems that reported long-COVID clinic visits. We matched 1,241 cases to 6,205 controls. We found similar results in this analysis (eTable 16 and eTable 17). However, confidence intervals for some risk factors were wide due to small sample size and limited power. XGBoost also showed similar features with this outcome (eFigure 1 Panel C).

Comparison of Risk Factors across PASC definitions In eTable 18 we compare the three case definitions of PASC (U09.9 alone, long-COVID visit alone, or either). Risk factors across the three possible definitions are fairly similar (eTable 2, eTable 18, and eTable 15). Across logistic regression models, statistical precision varies in part by sample size. The U09.9 sample appears to dominate the long-COVID visit sample in the combined model.

**eFigures**

**PASC defined as U09.9 or Long-COVID Clinic Visit**

**eFigure1. Feature Importance XGBoost (PASC defined as U09.9 or Long-COVID Clinic Visit)**

- **Panel A Unrestricted Controls (Method 1)**

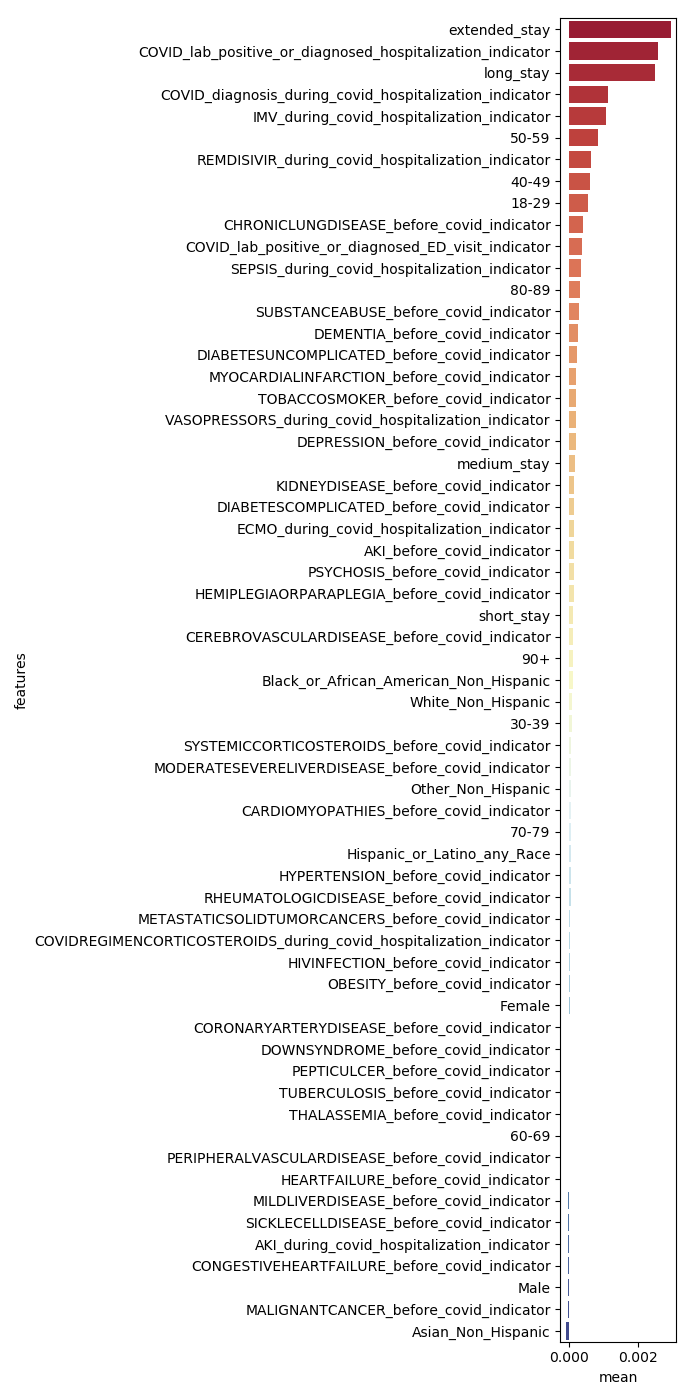

- **Panel B Less Restricted Controls (Method 2)**

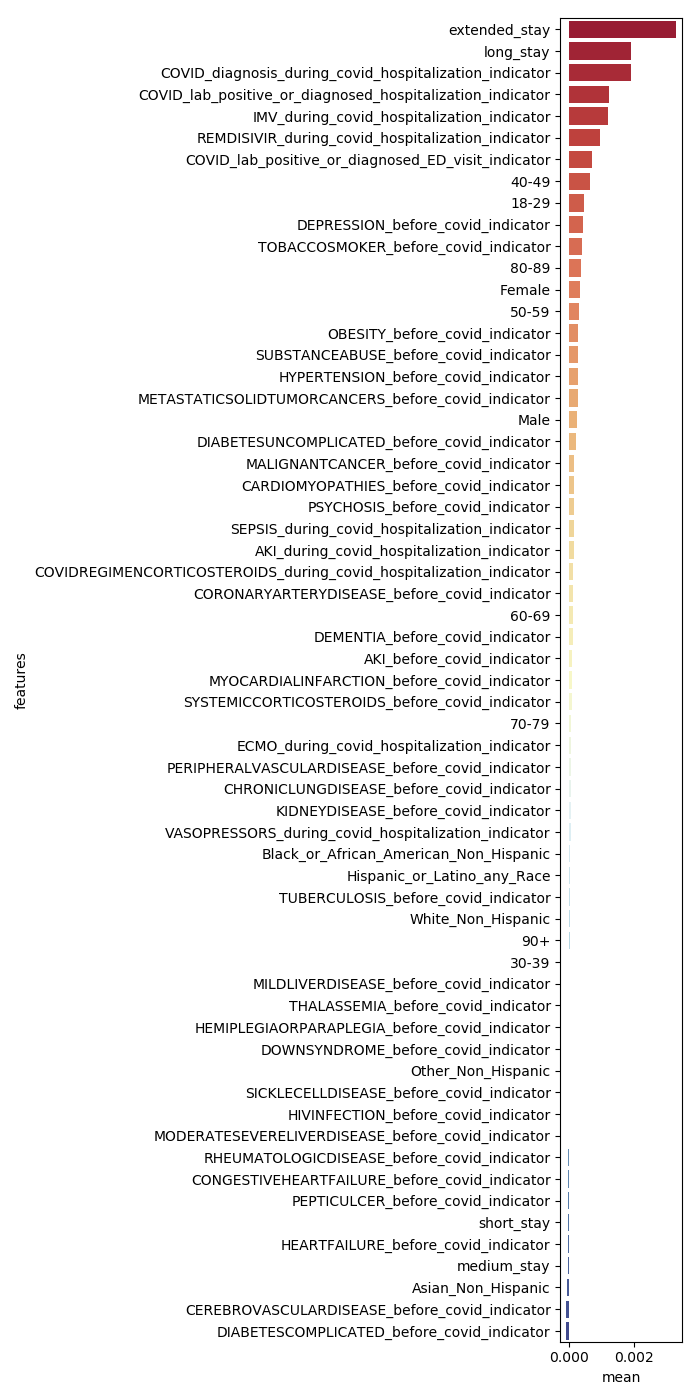

- **Panel C More Restricted Controls (Method 3)**

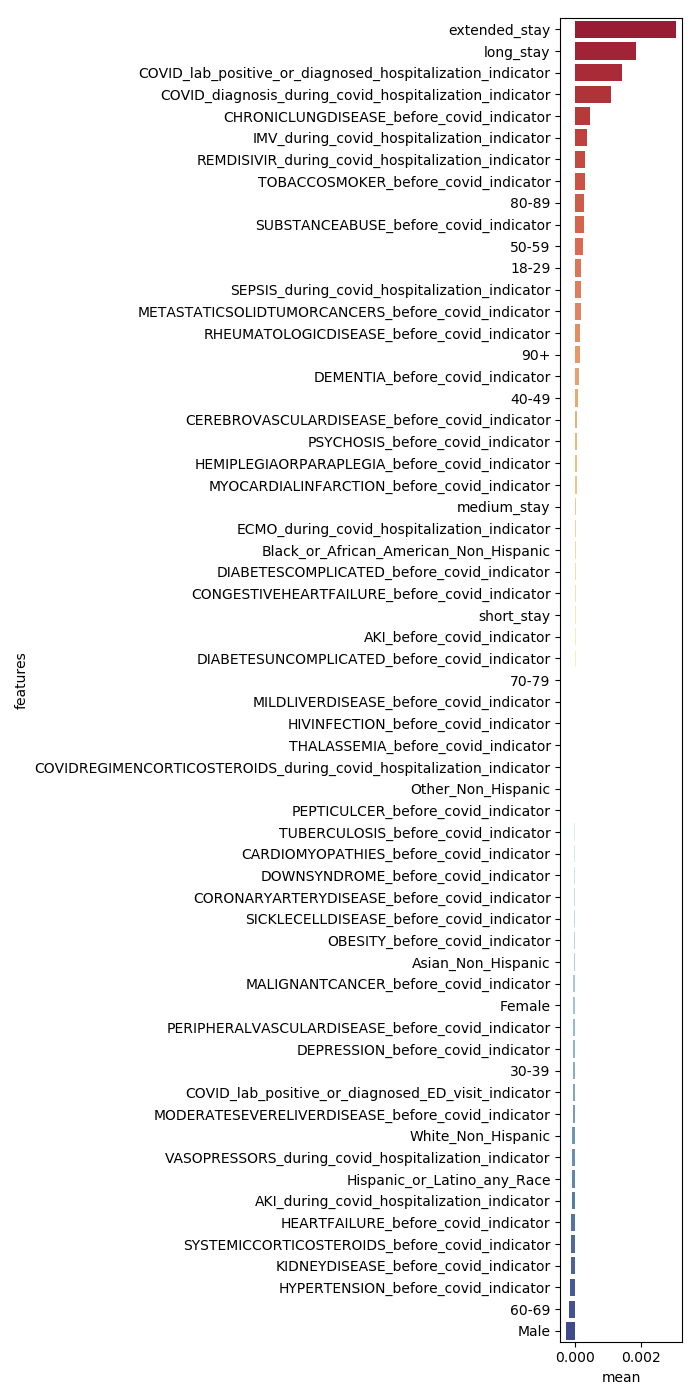

**eFigure 2. Forest Plot for Unrestricted Controls without SDoH (PASC defined as U09.9 or Long-COVID Clinic Visit)**

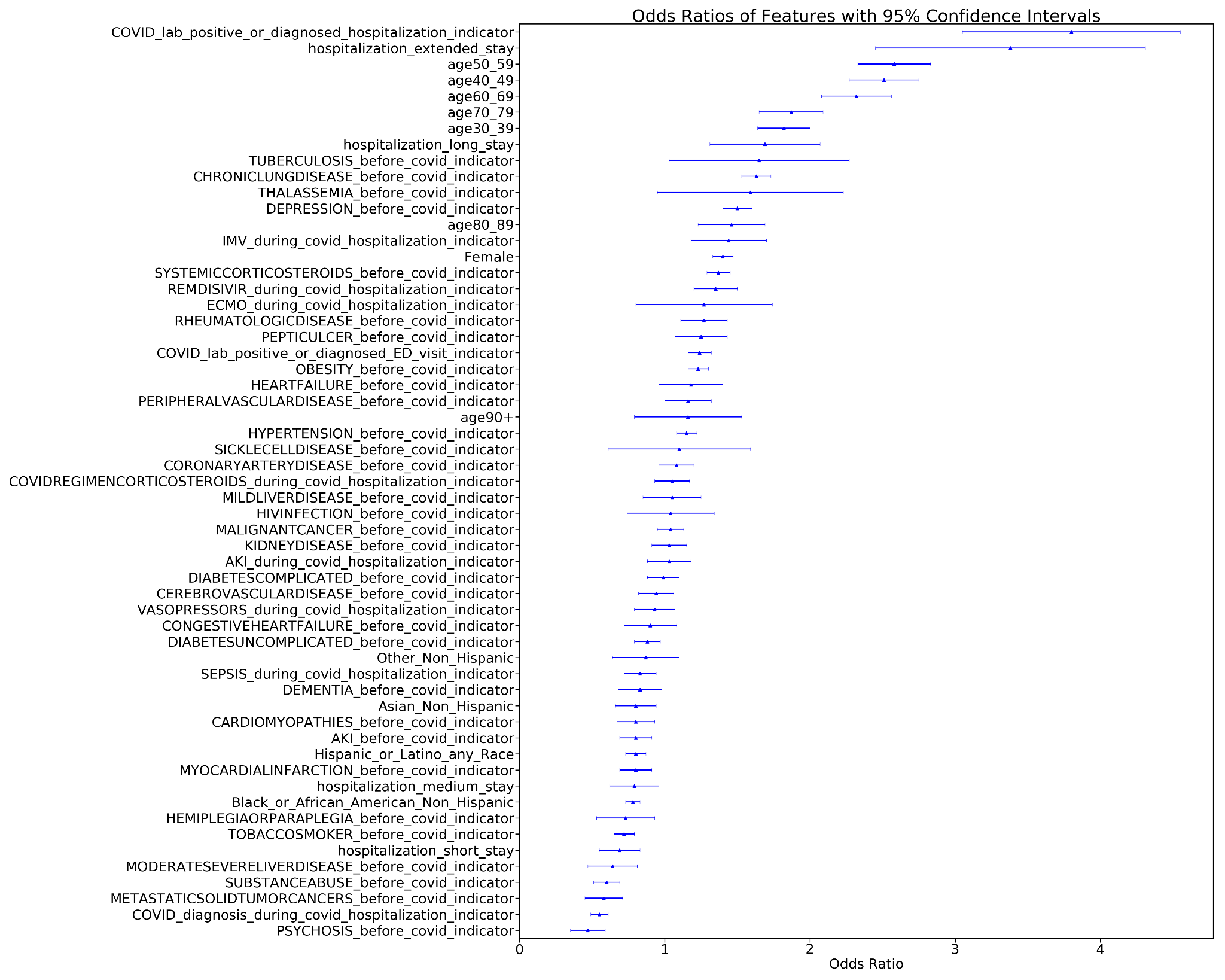

**eFigure 3. Feature Importance XGBoost for Unrestricted Controls with SDoH (PASC defined as U09.9 or Long-COVID Clinic Visit)**

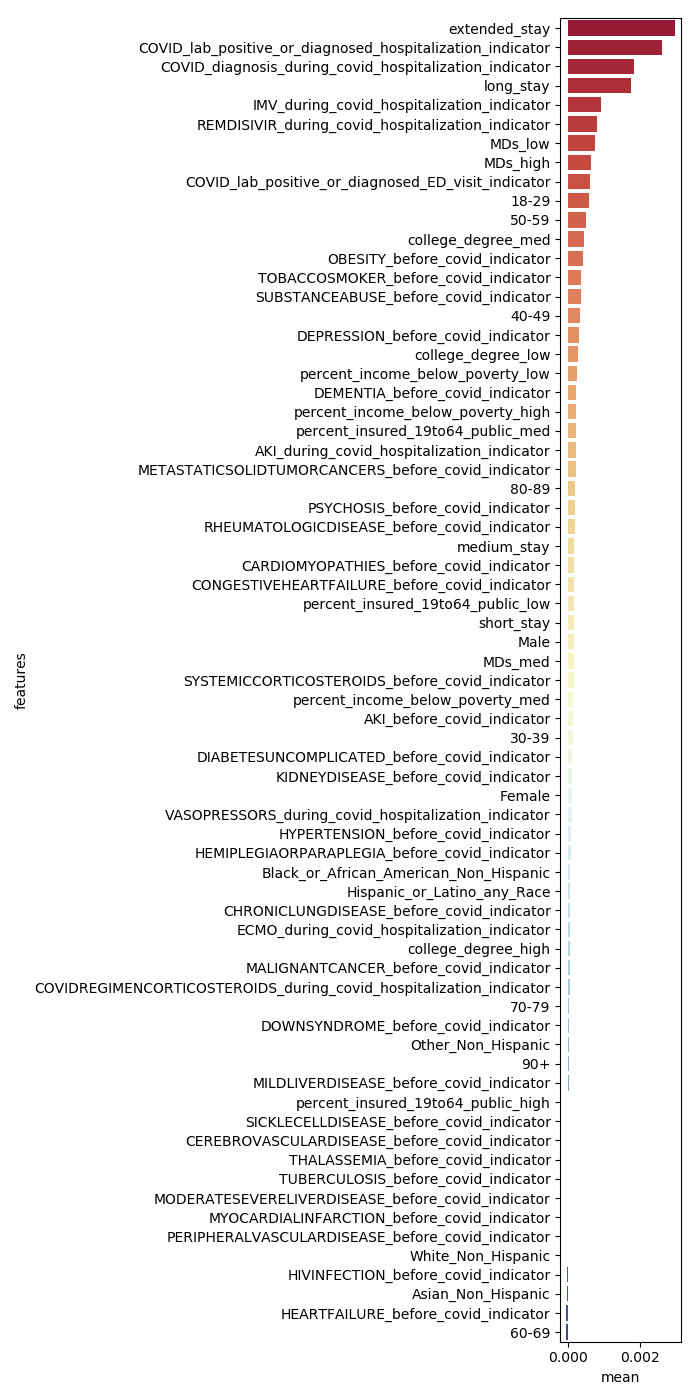

**eFigure 4. Feature Importance XGBoost for Unrestricted Controls with SDoH–Hospitalized Sample (PASC defined as U09.9 or Long COVID Visit)**

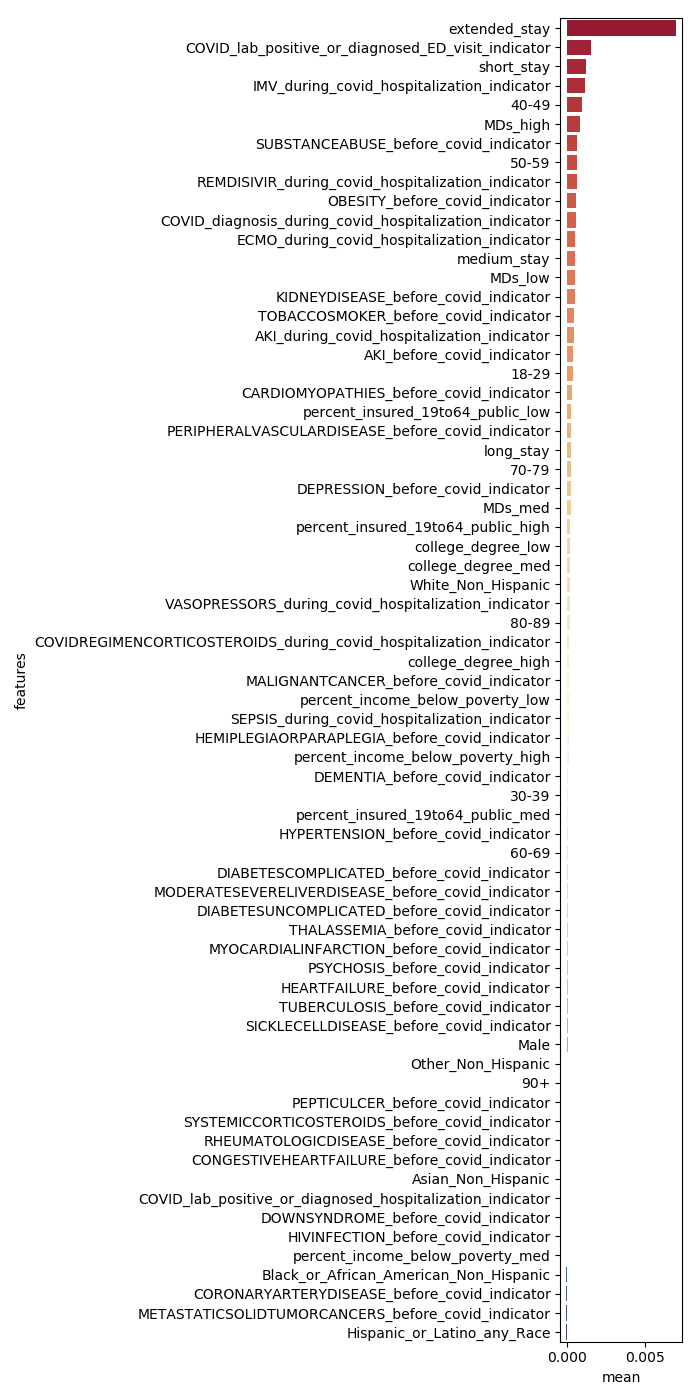

**eFigure 5. Feature Importance XGBoost for Unrestricted Controls with SDoH –non-Hospitalized Sample (PASC defined as U09.9 or Long COVID Visit)**
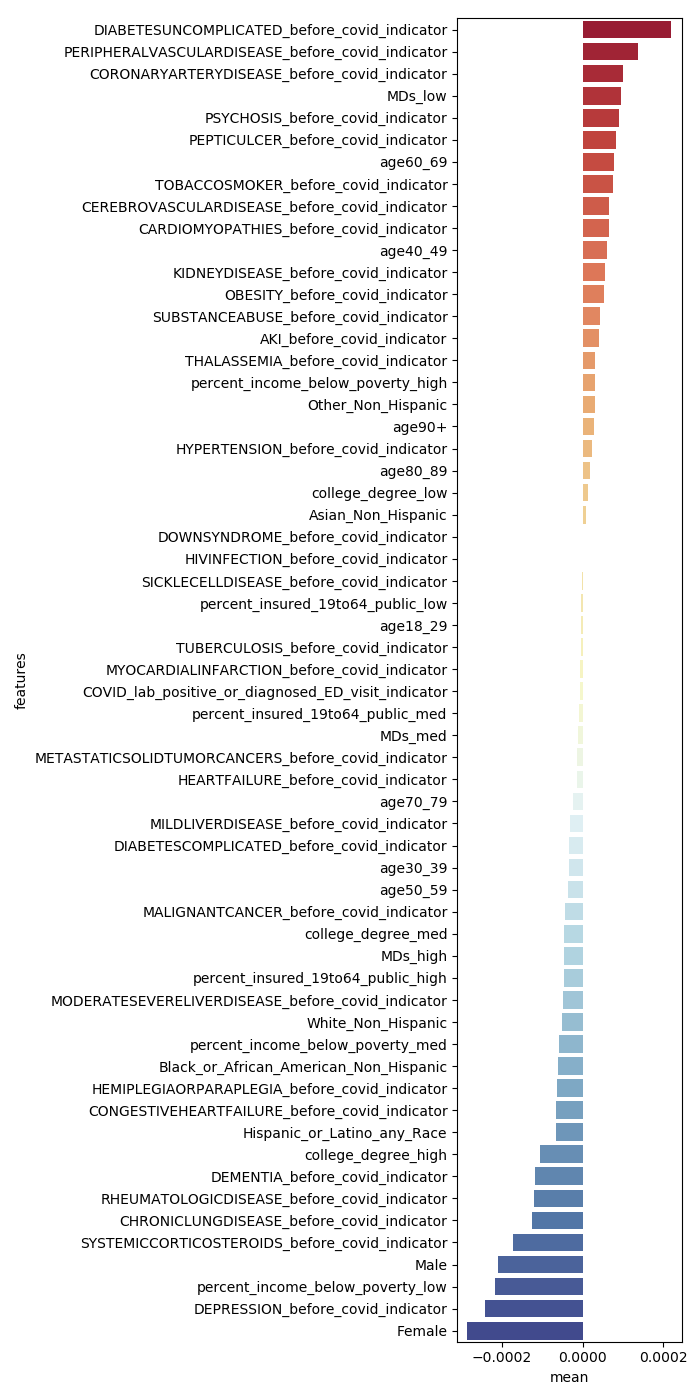

### **eTables**

**eTable 1. Cohort Characteristics for PASC cases defined by U09.9 or clinic visit (Additional Characteristics Not Shown in Table 1)**

|  | PASC  (N=8325) | Method 1  Unrestricted controls (N=41625) | Method 2  Restricted controls (N=41610) | Method 3  Most restricted controls (N=41610) |
| --- | --- | --- | --- | --- |
| **Demographics** |  |  |  |  |
| Age |  |  |  |  |
| 18-29 | 630 (7.57%) | 8366 (20.1%) | 8351 (20.1%) | 7887 (19.0%) |
| 30-39 | 1229 (14.8%) | 7920 (19.0%) | 7823 (18.8%) | 7426 (17.8%) |
| 40-49 | 1749 (21.0%) | 7321 (17.6%) | 6808 (16.4%) | 7022 (16.9%) |
| 50-59 | 1933 (23.2%) | 7171 (17.2%) | 6842 (16.4%) | 7161 (17.2%) |
| 60-69 | 1605 (19.3%) | 5806 (13.9%) | 5990 (14.4%) | 6400 (15.4%) |
| 70-79 | 840 (10.1%) | 3377 (8.1%) | 3773 (9.1%) | 3742 (9.0%) |
| 80-89 | 293 (3.5%) | 1391 (3.3%) | 1673 (4.0%) | 1588 (3.8%) |
| 90+ | 37 (0.4%) | 224 (0.5%) | 287 (0.7%) | 305 (0.7%) |
| **Comorbidities Prior to COVID Index Date** |  |  |  |  |
| AKI | 862 (10.4%) | 2287 (5.5%) | 2783 (6.7%) | 2738 (6.6%) |
| Cardiomyopathies | 225 (2.7%) | 768 (1.8%) | 927 (2.2%) | 897 (2.2%) |
| Cerebrovascular Disease | 390 (4.7%) | 1297 (3.1%) | 1707 (4.1%) | 1647 (4.0%) |
| Coronary Artery Disease | 832 (10.0%) | 2569 (6.2%) | 3319 (8.0%) | 3307 (7.9%) |
| Dementia | 153 (1.8%) | 643 (1.5%) | 846 (2.0%) | 832 (2.0%) |
| Down Syndrome | <20 | <20 | <20 | <20 |
| Heart Failure | 737 (8.9%) | 1936 (4.7%) | 2473 (5.9%) | 2375 (5.7%) |
| Hemiplegia or Paraplegia | 61 (0.7%) | 240 (0.6%) | 313 (0.8%) | 344 (0.8%) |
| HIV | 51 (0.6%) | 214 (0.5%) | 307 (0.7%) | 341 (0.8%) |
| Malignant Cancer | 837 (10.1%) | 2879 (6.9%) | 3826 (9.2%) | 3772 (9.1%) |
| Metastatic Solid Tumor Cancers | 91 (1.1%) | 426 (1.0%) | 568 (1.4%) | 551 (1.3%) |
| Mild Liver Disease | 170 (2.0%) | 600 (1.4%) | 780 (1.9%) | 791 (1.9%) |
| Moderate to Severe Liver Disease | 82 (1.0%) | 305 (0.7%) | 391 (0.9%) | 353 (0.8%) |
| Myocardial Infarction | 392 (4.7%) | 1311 (3.1%) | 1616 (3.9%) | 1659 (4.0%) |
| Peptic Ulcer | 279 (3.4%) | 714 (1.7%) | 915 (2.2%) | 954 (2.3%) |
| Peripheral Vascular Disease | 405 (4.9%) | 1045 (2.5%) | 1453 (3.5%) | 1362 (3.3%) |
| Rheumatologic Disease | 350 (4.2%) | 804 (1.9%) | 1020 (2.5%) | 981 (2.4%) |
| Sickle Cell Disease | <20 | 64 (0.2%) | 95 (0.2%) | 77 (0.2%) |
| Systemic Corticosteroids | 4325 (52.0%) | 13754 (33.0%) | 16385 (39.4%) | 16177 (38.9%) |
| Thalassemia | 21 (0.3%) | 66 (0.2%) | 78 (0.2%) | 89 (0.2%) |
| Tuberculosis | 27 (0.3%) | 69 (0.2%) | 107 (0.3%) | 90 (0.2%) |
| **Behavioral Health Indicators** |  |  |  |  |
| Depression | 2059 (24.7%) | 5851 (14.1%) | 7649 (18.4%) | 7526 (18.1%) |
| Psychosis | 65 (0.8%) | 416 (1.0%) | 483 (1.2%) | 471 (1.1%) |
| Substance Abuse | 205 (2.5%) | 1291 (3.1%) | 1541 (3.7%) | 1526 (3.7%) |
| Tobacco Smoker | 515 (6.2%) | 2656 (6.4%) | 3065 (7.4%) | 3122 (7.5%) |
| **Characteristics during Acute COVID Phase** |  |  |  |  |
| COVID Diagnosis during COVID-associated Hospitalization | 2065 (24.8%) | 4668 (11.2%) | 4534 (10.9%) | 4448 (10.7%) |
| Hospitalization stay |  |  |  |  |
| Short Stay (0-2 days) | 610 (7.3%) | 2155 (5.2%) | 2158 (5.2%) | 2188 (5.3%) |
| Medium Stay (3-7 days) | 870 (10.5%) | 2226 (5.3%) | 2282 (5.5%) | 2141 (5.1%) |
| Long Stay (8-30 days) | 1029 (12.4%) | 1274 (3.1%) | 1309 (3.1%) | 1270 (3.1%) |
| Extended Stay (31+ days) | 449 (5.4%) | 241 (0.6%) | 239 (0.6%) | 216 (0.5%) |

^a^Only captured for individuals hospitalized for COVID-19

**eTable 2. PASC Risk Factors from Logistic Regression (PASC defined as U09.9 or long-COVID Clinic Visit)**

|  | Method 1  Unrestricted Controls  (N=49950) | Method 2  Restricted  controls  (N=49932) | Method 3  Most Restricted  controls  (N=49932) |
| --- | --- | --- | --- |
| **Demographics** |  |  |  |
| Age |  |  |  |
| 18-29 | REF | REF | REF |
| 30-39 | 1.82 (1.64-2.02) | 1.96 (1.77-2.18) | 1.99 (1.79-2.21) |
| 40-49 | 2.51 (2.27-2.78) | 3.08 (2.78-3.4) | 2.81 (2.54-3.11) |
| 50-59 | 2.58 (2.33-2.86) | 3.24 (2.93-3.59) | 2.9 (2.62-3.21) |
| 60-69 | 2.32 (2.08-2.58) | 2.73 (2.46-3.04) | 2.47 (2.22-2.75) |
| 70-79 | 1.87 (1.65-2.12) | 2.15 (1.9-2.43) | 1.99 (1.76-2.26) |
| 80-89 | 1.46 (1.23-1.73) | 1.57 (1.33-1.85) | 1.56 (1.32-1.84) |
| 90+ | 1.16 (0.79-1.7) | 1.05 (0.72-1.53) | 0.93 (0.64-1.35) |
| Sex |  |  |  |
| Male or Unknown Sex | REF | REF | REF |
| Female | 1.4 (1.33-1.48) | 1.33 (1.26-1.41) | 1.28 (1.21-1.35) |
| Race/ethnicity |  |  |  |
| White NH | REF | REF | REF |
| Hispanic | 0.8 (0.73-0.87) | 0.85 (0.78-0.93) | 0.82 (0.75-0.9) |
| Black NH | 0.78 (0.73-0.85) | 0.8 (0.75-0.86) | 0.77 (0.72-0.83) |
| Asian NH | 0.8 (0.66-0.97) | 0.71 (0.59-0.87) | 0.72 (0.6-0.88) |
| Other race NH | 0.87 (0.64-1.19) | 0.99 (0.72-1.35) | 0.98 (0.71-1.33) |
| **Comorbidities Prior to COVID Index Date** |  |  |  |
| AKI | 0.8 (0.69-0.94) | 0.84 (0.72-0.98) | 0.8 (0.69-0.93) |
| Cardiomyopathies | 0.8 (0.67-0.96) | 0.92 (0.77-1.1) | 0.88 (0.73-1.05) |
| Cerebrovascular Disease | 0.94 (0.82-1.08) | 0.87 (0.76-0.99) | 0.91 (0.8-1.04) |
| Chronic Lung Disease | 1.63 (1.53-1.74) | 1.57 (1.47-1.67) | 1.63 (1.53-1.74) |
| Complicated Diabetes | 0.99 (0.88-1.12) | 0.99 (0.87-1.11) | 1.07 (0.94-1.21) |
| Congestive Heart Failure | 0.9 (0.72-1.14) | 0.87 (0.69-1.09) | 0.9 (0.72-1.13) |
| Coronary Artery Disease | 1.08 (0.96-1.2) | 1.03 (0.93-1.15) | 1.03 (0.92-1.14) |
| Dementia | 0.83 (0.68-1.01) | 0.74 (0.61-0.9) | 0.76 (0.63-0.92) |
| Down Syndrome | 4.02 (0.53-30.38) | 1.73 (0.4-7.39) | 3.08 (0.6-15.94) |
| Heart Failure | 1.18 (0.96-1.44) | 1.18 (0.96-1.45) | 1.19 (0.97-1.46) |
| Hemiplegia or Paraplegia | 0.73 (0.53-0.99) | 0.7 (0.52-0.95) | 0.53 (0.39-0.72) |
| HIV | 1.04 (0.74-1.45) | 0.85 (0.62-1.17) | 0.77 (0.57-1.06) |
| Hypertension | 1.15 (1.08-1.23) | 0.98 (0.92-1.05) | 0.98 (0.92-1.05) |
| Kidney Disease | 1.03 (0.91-1.17) | 1.03 (0.91-1.16) | 1.07 (0.95-1.21) |
| Malignant Cancer | 1.04 (0.95-1.15) | 0.93 (0.85-1.02) | 0.94 (0.85-1.03) |
| Metastatic Solid Tumor Cancers | 0.58 (0.45-0.75) | 0.61 (0.47-0.78) | 0.6 (0.47-0.77) |
| Mild Liver Disease | 1.05 (0.85-1.29) | 0.93 (0.76-1.14) | 0.88 (0.72-1.07) |
| Moderate to Severe Liver Disease | 0.64 (0.47-0.85) | 0.64 (0.48-0.85) | 0.76 (0.57-1.03) |
| Myocardial Infarction | 0.8 (0.69-0.93) | 0.79 (0.68-0.91) | 0.77 (0.66-0.88) |
| Obesity | 1.23 (1.16-1.3) | 1.04 (0.99-1.1) | 1.06 (1.0-1.12) |
| Peptic Ulcer | 1.25 (1.07-1.46) | 1.23 (1.06-1.42) | 1.17 (1.01-1.35) |
| Peripheral Vascular Disease | 1.16 (1.0-1.33) | 1.06 (0.92-1.21) | 1.11 (0.97-1.27) |
| Rheumatologic Disease | 1.27 (1.11-1.46) | 1.22 (1.07-1.4) | 1.31 (1.14-1.5) |
| Sickle Cell Disease | 1.1 (0.61-1.96) | 0.87 (0.5-1.52) | 0.89 (0.5-1.58) |
| Systemic Corticosteroids | 1.37 (1.29-1.45) | 1.19 (1.12-1.25) | 1.21 (1.15-1.28) |
| Thalassemia | 1.59 (0.95-2.68) | 1.41 (0.84-2.37) | 1.2 (0.72-2.01) |
| Tuberculosis | 1.65 (1.03-2.65) | 1.16 (0.74-1.81) | 1.53 (0.97-2.41) |
| Uncomplicated Diabetes | 0.88 (0.79-0.98) | 0.87 (0.79-0.97) | 0.82 (0.74-0.91) |
| **Behavioral Health** |  |  |  |
| Depression | 1.5 (1.4-1.6) | 1.3 (1.22-1.39) | 1.34 (1.26-1.43) |
| Psychosis | 0.47 (0.35-0.62) | 0.56 (0.42-0.74) | 0.56 (0.42-0.74) |
| Substance Abuse | 0.6 (0.51-0.71) | 0.59 (0.5-0.69) | 0.59 (0.5-0.7) |
| Tobacco Smoker | 0.72 (0.65-0.81) | 0.72 (0.65-0.8) | 0.66 (0.6-0.74) |
| **Characteristics of Index COVID "Acute Phase"** |  |  |  |
| COVID Diagnosis during COVID-associated Hospitalization | 0.55 (0.49-0.62) | 0.73 (0.66-0.81) | 0.68 (0.61-0.76) |
| COVID-associated Hospitalization | 3.8 (3.05-4.73) | 3.35 (2.72-4.13) | 3.08 (2.5-3.79) |
| COVID-associated ED Visit | 1.24 (1.16-1.33) | 1.4 (1.31-1.5) | 1.44 (1.34-1.54) |
| Hospitalization stay |  |  |  |
| Not Hospitalized | REF | REF | REF |
| Short Stay | 0.69 (0.55-0.88) | 0.67 (0.54-0.85) | 0.75 (0.6-0.94) |
| Medium Stay | 0.79 (0.62-1.01) | 0.74 (0.59-0.94) | 0.87 (0.69-1.1) |
| Long Stay | 1.69 (1.31-2.17) | 1.54 (1.21-1.97) | 1.79 (1.41-2.28) |
| Extended Stay | 3.38 (2.45-4.67) | 2.78 (2.02-3.82) | 3.44 (2.51-4.73) |
| COVID treatment |  |  |  |
| Corticosteroidsᵃ | 1.05 (0.93-1.18) | 1.14 (1.01-1.28) | 1.2 (1.07-1.35) |
| Remdesivirᵃ | 1.35 (1.2-1.51) | 1.53 (1.36-1.72) | 1.59 (1.41-1.79) |
| Vasopressorsᵃ | 0.93 (0.79-1.09) | 0.9 (0.77-1.06) | 0.81 (0.69-0.95) |
| ECMOᵃ | 1.27 (0.8-2.0) | 1.66 (1.0-2.78) | 2.58 (1.45-4.6) |
| Mechanical Ventilationᵃ | 1.44 (1.18-1.74) | 1.72 (1.42-2.09) | 1.6 (1.32-1.94) |
| AKI during COVID-associated Hospitalization | 1.03 (0.88-1.2) | 1.0 (0.86-1.16) | 1.02 (0.88-1.19) |
| Sepsis during COVID-associated Hospitalization | 0.83 (0.72-0.96) | 0.81 (0.7-0.93) | 0.86 (0.75-1.0) |

ᵃOnly captured for individuals hospitalized for COVID-19

Odds ratios presented with 95% CI in parenthesis

**eTable 3. AUC scores for our three models of PASC defined by U09.9 or Long-COVID Clinic Visit**

| **Cohorts** | **XGBoost (AUC score)** | **RF (AUC score)** | **LR (AUC score)** |
| --- | --- | --- | --- |
| Cohort A | 0.713 | 0.665 | 0.709 |
| Cohort B | 0.731 | 0.685 | 0.726 |
| Cohort C | 0.712 | 0.644 | 0.708 |

**eTable 4. Comparison of Feature Importance Across Models for Unrestricted Sample (PASC defined as U09.9 or Long-COVID Clinic Visit)**

| features | Random Forest | XGBoost | Logistic Regression | Mean Rank |
| --- | --- | --- | --- | --- |
| Hospitalization Extended Stay (31+ days) | 4 | 1 | 2 | 2.33 |
| COVID-associated Hospitalization | 1 | 4 | 4 | 3.00 |
| COVID Diagnosis during COVID-associated Hospitalization | 5 | 3 | 11 | 6.33 |
| Age 40-49 | 7 | 8 | 6 | 7.00 |
| Age 50-59 | 6 | 14 | 5 | 8.33 |
| Hospitalization Long Stay (8-30 days) | 18 | 2 | 8 | 9.33 |
| Age 18-29 | 11 | 9 | ref. | 10.00 |
| Male | 3 | 19 | ref. | 11.00 |
| Female | 2 | 13 | 22 | 12.33 |
| Depression | 15 | 10 | 19 | 14.67 |
| Age 60-69 | 10 | 28 | 7 | 15.00 |
| COVID Treatment: Mechanical Ventilation | 24 | 5 | 21 | 16.67 |
| COVID Treatment: Remdesivir | 20 | 6 | 26 | 17.33 |
| Age 70-79 | 16 | 35 | 10 | 20.33 |
| Chronic Lung Disease | 9 | 38 | 16 | 21.00 |
| Substance Abuse | 37 | 16 | 14 | 22.33 |
| COVID-associated ED Visit | 25 | 7 | 36 | 22.67 |
| Tobacco Smoker | 34 | 11 | 23 | 22.67 |
| Cardiomyopathies | 17 | 22 | 32 | 23.67 |
| Age 30-39 | 8 | 53 | 12 | 24.33 |
| Metastatic Solid Tumor Cancers | 45 | 18 | 13 | 25.33 |
| Psychosis | 44 | 23 | 9 | 25.33 |
| Uncomplicated Diabetes | 13 | 20 | 45 | 26.00 |
| Obesity | 27 | 15 | 37 | 26.33 |
| Systemic Corticosteroids | 22 | 34 | 24 | 26.67 |
| Myocardial Infarction | 19 | 32 | 33 | 28.00 |
| Malignant Cancer | 12 | 21 | 54 | 29.00 |
| Age 80-89 | 56 | 12 | 20 | 29.33 |
| Coronary Artery Disease | 14 | 27 | 49 | 30.00 |
| Dementia | 26 | 29 | 38 | 31.00 |
| Hypertension | 36 | 17 | 44 | 32.33 |
| Race/Ethnicity: Black NH | 29 | 42 | 28 | 33.00 |
| AKI prior to COVID | 41 | 31 | 35 | 35.67 |
| Race/Ethnicity: Asian NH | 40 | 39 | 30 | 36.33 |
| Peripheral Vascular Disease | 31 | 37 | 41 | 36.33 |
| Rheumatologic Disease | 30 | 52 | 27 | 36.33 |
| Race/Ethnicity: White NH | 23 | 50 | ref. | 36.50 |
| COVID Treatment: Corticosteroids | 33 | 26 | 55 | 38.00 |
| COVID Treatment: ECMO | 50 | 36 | 29 | 38.33 |
| Sepsis during COVID-associated Hospitalization | 52 | 24 | 39 | 38.33 |
| Complicated Diabetes | 28 | 30 | 58 | 38.67 |
| Kidney Disease | 21 | 40 | 56 | 39.00 |
| Hospitalization Short Stay (0-2 days) | 32 | 47 | ref. | 39.50 |
| Moderate to Severe Liver Disease | 48 | 55 | 18 | 40.33 |
| Tuberculosis | 60 | 46 | 15 | 40.33 |
| Cerebrovascular Disease | 38 | 33 | 51 | 40.67 |
| Race/Ethnicity: Hispanic | 46 | 45 | 31 | 40.67 |
| Down Syndrome | 61 | 61 | 3 | 41.67 |
| COVID Treatment: Vasopressors | 35 | 41 | 50 | 42.00 |
| Heart Failure | 43 | 44 | 40 | 42.33 |
| Thalassemia | 54 | 56 | 17 | 42.33 |
| AKI during COVID-associated Hospitalization | 47 | 25 | 57 | 43.00 |
| Peptic Ulcer | 51 | 48 | 34 | 44.33 |
| Congestive Heart Failure | 42 | 49 | 46 | 45.67 |
| Hospitalization Medium Stay (3-7 days) | 49 | 43 | 48 | 46.67 |
| Hemiplegia or Paraplegia | 59 | 58 | 25 | 47.33 |
| Mild Liver Disease | 39 | 54 | 53 | 48.67 |
| Age 90+ | 57 | 51 | 42 | 50.00 |
| Race/Ethnicity: Other race NH | 53 | 60 | 43 | 52.00 |
| HIV | 55 | 57 | 52 | 54.67 |
| Sickle Cell Disease | 58 | 59 | 47 | 54.67 |

**eTable 5. Comparison of Feature Importance Across Models for Restricted Sample (PASC defined as U09.9 or Long-COVID Clinic Visit)**

| features | Random Forest | XGBoost | Logistic Regression | Mean Rank |
| --- | --- | --- | --- | --- |
| COVID-associated Hospitalization | 1 | 2 | 6 | 3.00 |
| Hospitalization Extended Stay (31+ days) | 9 | 1 | 2 | 4.00 |
| Age 50-59 | 4 | 6 | 3 | 4.33 |
| Age 40-49 | 6 | 8 | 4 | 6.00 |
| Hospitalization Long Stay (8-30 days) | 12 | 3 | 7 | 7.33 |
| Age 18-29 | 10 | 9 | ref. | 9.50 |
| Chronic Lung Disease | 5 | 10 | 16 | 10.33 |
| COVID Treatment: Mechanical Ventilation | 15 | 5 | 11 | 10.33 |
| COVID Diagnosis during COVID-associated Hospitalization | 13 | 4 | 22 | 13.00 |
| COVID Treatment: Remdesivir | 23 | 7 | 19 | 16.33 |
| Age 30-39 | 8 | 34 | 9 | 17.00 |
| COVID-associated ED Visit | 17 | 11 | 25 | 17.67 |
| Substance Abuse | 34 | 14 | 13 | 20.33 |
| Age 70-79 | 14 | 39 | 8 | 20.33 |
| Tobacco Smoker | 24 | 18 | 24 | 22.00 |
| Age 60-69 | 7 | 56 | 5 | 22.67 |
| Dementia | 28 | 15 | 26 | 23.00 |
| Psychosis | 36 | 26 | 10 | 24.00 |
| Hospitalization Short Stay (0-2 days) | 20 | 28 | ref. | 24.00 |
| Age 80-89 | 42 | 13 | 18 | 24.33 |
| Male | 3 | 46 | ref. | 24.50 |
| Race/Ethnicity: Black NH | 16 | 31 | 30 | 25.67 |
| Myocardial Infarction | 32 | 17 | 29 | 26.00 |
| Female | 2 | 53 | 27 | 27.33 |
| Race/Ethnicity: White NH | 22 | 33 | ref. | 27.50 |
| Sepsis during COVID-associated Hospitalization | 41 | 12 | 31 | 28.00 |
| Systemic Corticosteroids | 18 | 35 | 35 | 29.33 |
| COVID Treatment: ECMO | 54 | 24 | 14 | 30.67 |
| Depression | 45 | 20 | 28 | 31.00 |
| Uncomplicated Diabetes | 35 | 16 | 43 | 31.33 |
| Cerebrovascular Disease | 29 | 29 | 40 | 32.67 |
| AKI prior to COVID | 40 | 25 | 34 | 33.00 |
| Complicated Diabetes | 21 | 23 | 57 | 33.67 |
| Cardiomyopathies | 19 | 38 | 46 | 34.33 |
| Hemiplegia or Paraplegia | 57 | 27 | 20 | 34.67 |
| Race/Ethnicity: Asian NH | 49 | 32 | 23 | 34.67 |
| Kidney Disease | 31 | 22 | 54 | 35.67 |
| Metastatic Solid Tumor Cancers | 50 | 43 | 15 | 36.00 |
| Moderate to Severe Liver Disease | 59 | 36 | 17 | 37.33 |
| COVID Treatment: Vasopressors | 48 | 19 | 45 | 37.33 |
| Congestive Heart Failure | 25 | 47 | 41 | 37.67 |
| COVID Treatment: Corticosteroids | 26 | 44 | 44 | 38.00 |
| Hospitalization Medium Stay (3-7 days) | 47 | 21 | 50 | 39.33 |
| Rheumatologic Disease | 44 | 42 | 33 | 39.67 |
| Coronary Artery Disease | 11 | 58 | 53 | 40.67 |
| Age 90+ | 43 | 30 | 51 | 41.33 |
| Heart Failure | 38 | 54 | 36 | 42.67 |
| Malignant Cancer | 37 | 45 | 47 | 43.00 |
| Race/Ethnicity: Hispanic | 52 | 40 | 37 | 43.00 |
| Mild Liver Disease | 33 | 50 | 48 | 43.67 |
| Obesity | 27 | 52 | 52 | 43.67 |
| Peptic Ulcer | 39 | 60 | 32 | 43.67 |
| Down Syndrome | 61 | 59 | 12 | 44.00 |
| Thalassemia | 55 | 57 | 21 | 44.33 |
| Peripheral Vascular Disease | 30 | 55 | 49 | 44.67 |
| Sickle Cell Disease | 53 | 49 | 42 | 48.00 |
| HIV | 56 | 51 | 38 | 48.33 |
| Hypertension | 51 | 41 | 55 | 49.00 |
| Race/Ethnicity: Other race NH | 58 | 37 | 56 | 50.33 |
| AKI during COVID-associated Hospitalization | 46 | 48 | 58 | 50.67 |
| Tuberculosis | 60 | 61 | 39 | 53.33 |

**eTable 6. Comparison of Feature Importance Across Models for More Restricted Sample (PASC defined as U09.9 or Long-COVID Clinic Visit)**

| features | Random Forest | XGBoost | Logistic Regression | Mean Rank |
| --- | --- | --- | --- | --- |
| COVID-associated Hospitalization | 1 | 3 | 8 | 4.00 |
| Hospitalization Extended Stay (31+ days) | 10 | 1 | 2 | 4.33 |
| Age 50-59 | 7 | 11 | 4 | 7.33 |
| Male | 3 | 12 | ref. | 7.50 |
| Hospitalization Long Stay (8-30 days) | 15 | 2 | 9 | 8.67 |
| Chronic Lung Disease | 9 | 5 | 16 | 10.00 |
| Age 60-69 | 8 | 16 | 7 | 10.33 |
| Age 40-49 | 4 | 24 | 5 | 11.00 |
| COVID Diagnosis during COVID-associated Hospitalization | 11 | 4 | 22 | 12.33 |
| Age 18-29 | 14 | 13 | ref. | 13.50 |
| COVID Treatment: Mechanical Ventilation | 18 | 6 | 17 | 13.67 |
| COVID Treatment: Remdesivir | 23 | 7 | 18 | 16.00 |
| Age 30-39 | 6 | 37 | 11 | 18.00 |
| Age 80-89 | 29 | 9 | 19 | 19.00 |
| Substance Abuse | 38 | 10 | 14 | 20.67 |
| Metastatic Solid Tumor Cancers | 33 | 15 | 15 | 21.00 |
| Dementia | 21 | 20 | 26 | 22.33 |
| Tobacco Smoker | 42 | 8 | 21 | 23.67 |
| Rheumatologic Disease | 28 | 17 | 28 | 24.33 |
| Age 70-79 | 13 | 51 | 10 | 24.67 |
| Female | 2 | 42 | 32 | 25.33 |
| Systemic Corticosteroids | 19 | 22 | 37 | 26.00 |
| Myocardial Infarction | 16 | 33 | 29 | 26.00 |
| Sepsis during COVID-associated Hospitalization | 31 | 14 | 42 | 29.00 |
| Race/Ethnicity: Hispanic | 27 | 26 | 36 | 29.67 |
| Hypertension | 12 | 19 | 58 | 29.67 |
| Race/Ethnicity: Black NH | 20 | 38 | 31 | 29.67 |
| COVID Treatment: Vasopressors | 30 | 27 | 34 | 30.33 |
| Hemiplegia or Paraplegia | 50 | 31 | 12 | 31.00 |
| Heart Failure | 34 | 23 | 38 | 31.67 |
| Depression | 32 | 39 | 25 | 32.00 |
| COVID-associated ED Visit | 40 | 34 | 23 | 32.33 |
| Psychosis | 55 | 30 | 13 | 32.67 |
| COVID Treatment: ECMO | 60 | 36 | 6 | 34.00 |
| Uncomplicated Diabetes | 22 | 48 | 35 | 35.00 |
| Kidney Disease | 36 | 21 | 51 | 36.00 |
| Complicated Diabetes | 17 | 41 | 52 | 36.67 |
| Cerebrovascular Disease | 35 | 28 | 49 | 37.33 |
| Coronary Artery Disease | 5 | 54 | 55 | 38.00 |
| Age 90+ | 47 | 18 | 50 | 38.33 |
| Race/Ethnicity: Asian NH | 46 | 46 | 24 | 38.67 |
| Moderate to Severe Liver Disease | 59 | 32 | 27 | 39.33 |
| Race/Ethnicity: White NH | 51 | 29 | ref. | 40.00 |
| Down Syndrome | 61 | 56 | 3 | 40.00 |
| AKI during COVID-associated Hospitalization | 39 | 25 | 56 | 40.00 |
| Hospitalization Medium Stay (3-7 days) | 43 | 35 | 47 | 41.67 |
| Peripheral Vascular Disease | 37 | 40 | 48 | 41.67 |
| COVID Treatment: Corticosteroids | 26 | 60 | 40 | 42.00 |
| Cardiomyopathies | 25 | 57 | 44 | 42.00 |
| Obesity | 24 | 49 | 54 | 42.33 |
| AKI prior to COVID | 48 | 47 | 33 | 42.67 |
| Tuberculosis | 57 | 58 | 20 | 45.00 |
| HIV | 52 | 53 | 30 | 45.00 |
| Congestive Heart Failure | 45 | 44 | 46 | 45.00 |
| Malignant Cancer | 44 | 43 | 53 | 46.67 |
| Hospitalization Short Stay (0-2 days) | 49 | 45 | ref. | 47.00 |
| Peptic Ulcer | 41 | 59 | 41 | 47.00 |
| Mild Liver Disease | 53 | 52 | 43 | 49.33 |
| Thalassemia | 56 | 55 | 39 | 50.00 |
| Sickle Cell Disease | 58 | 50 | 45 | 51.00 |
| Race/Ethnicity: Other races NH | 54 | 61 | 57 | 57.33 |

**eTable 7. Comparison of Feature Importance Across Models for Unrestricted Sample including SDOH variables (PASC defined as U09.9 or Long-COVID Clinic Visit)**

| features | Random Forest SDOH | XGBoost SDOH | Logistic Regression SDOH | Mean Rank |
| --- | --- | --- | --- | --- |
| Hospitalization Extended Stay (31+ days) | 19 | 1 | 2 | 7.33 |
| MDs per 1000 residents: Low (<1.91%) | 10 | 7 | ref. | 8.50 |
| COVID-associated Hospitalization | 22 | 2 | 4 | 9.33 |
| Age 40-49 | 8 | 16 | 6 | 10.00 |
| Age 50-59 | 15 | 11 | 5 | 10.33 |
| Hospitalization Long Stay (8-30 days) | 23 | 4 | 8 | 11.67 |
| Households with Income below poverty: low (<11%) | 5 | 19 | ref. | 12.00 |
| College Degree low (<19%) | 7 | 18 | ref. | 12.50 |
| COVID Diagnosis during COVID-associated Hospitalization | 26 | 3 | 11 | 13.33 |
| MDs per 1000 residents: High (>3.61%) | 6 | 8 | 30 | 14.67 |
| Age 18-29 | 20 | 10 | ref. | 15.00 |
| Male | 2 | 33 | ref. | 17.50 |
| COVID Treatment: Mechanical Ventilation | 27 | 5 | 22 | 18.00 |
| Age 30-39 | 11 | 38 | 12 | 20.33 |
| College Degree medium (19-25%) | 3 | 12 | 50 | 21.67 |
| Public health Insurance for ages 19-64: Low (<13%) | 13 | 31 | ref. | 22.00 |
| Depression | 31 | 17 | 19 | 22.33 |
| Female | 1 | 42 | 24 | 22.33 |
| Substance Abuse | 38 | 15 | 15 | 22.67 |
| Age 60-69 | 17 | 49 | 7 | 24.33 |
| MDs per 1000 residents: medium (1.91-3.61%) | 4 | 34 | 36 | 24.67 |
| Psychosis | 45 | 26 | 9 | 26.67 |
| Tobacco Smoker | 40 | 14 | 26 | 26.67 |
| Obesity | 30 | 13 | 39 | 27.33 |
| Chronic Lung Disease | 16 | 53 | 17 | 28.67 |
| Age 80-89 | 43 | 25 | 20 | 29.33 |
| Households with Income below poverty: high (>15%) | 12 | 21 | 57 | 30.00 |
| COVID-associated ED Visit | 42 | 9 | 40 | 30.33 |
| Metastatic Solid Tumor Cancers | 51 | 24 | 18 | 31.00 |
| COVID Treatment: Remdesivir | 56 | 6 | 31 | 31.00 |
| Age 70-79 | 24 | 60 | 10 | 31.33 |
| Households with Income below poverty: medium (11-15%) | 9 | 36 | 49 | 31.33 |
| Systemic Corticosteroids | 36 | 35 | 27 | 32.67 |
| Dementia | 44 | 20 | 35 | 33.00 |
| Race/Ethnicity: Black NH | 25 | 51 | 25 | 33.67 |
| Public health Insurance for ages 19-64: Medium (13-18%) | 18 | 22 | 66 | 35.33 |
| Cardiomyopathies | 49 | 29 | 32 | 36.67 |
| Hospitalization Medium Stay (3-7 days) | 33 | 28 | 52 | 37.67 |
| Race/Ethnicity: Hispanic | 29 | 52 | 34 | 38.33 |
| Rheumatologic Disease | 58 | 27 | 33 | 39.33 |
| Hospitalization Short Stay (0-2 days) | 47 | 32 | ref. | 39.50 |
| Uncomplicated Diabetes | 35 | 40 | 47 | 40.67 |
| Race/Ethnicity: White NH | 21 | 63 | ref. | 42.00 |
| College Degree high (>25%) | 14 | 55 | 58 | 42.33 |
| Race/Ethnicity: Asian NH | 41 | 58 | 29 | 42.67 |
| Hypertension | 39 | 45 | 44 | 42.67 |
| Hemiplegia or Paraplegia | 62 | 48 | 21 | 43.67 |
| Sepsis during COVID-associated Hospitalization | 48 | 39 | 46 | 44.33 |
| Peptic Ulcer | 50 | 47 | 37 | 44.67 |
| Kidney Disease | 32 | 41 | 62 | 45.00 |
| AKI prior to COVID | 57 | 37 | 42 | 45.33 |
| Complicated Diabetes | 28 | 46 | 63 | 45.67 |
| Down Syndrome | 73 | 61 | 3 | 45.67 |
| COVID Treatment: Vasopressors | 37 | 44 | 56 | 45.67 |
| Congestive Heart Failure | 65 | 30 | 45 | 46.67 |
| COVID Treatment: ECMO | 59 | 54 | 28 | 47.00 |
| Heart Failure | 52 | 50 | 41 | 47.67 |
| Thalassemia | 60 | 70 | 14 | 48.00 |
| AKI during COVID-associated Hospitalization | 63 | 23 | 59 | 48.33 |
| Coronary Artery Disease | 55 | 43 | 54 | 50.67 |
| Tuberculosis | 71 | 68 | 13 | 50.67 |
| Moderate to Severe Liver Disease | 72 | 67 | 16 | 51.67 |
| Public health Insurance for ages 19-64: High (>18%) | 34 | 71 | 53 | 52.67 |
| Peripheral Vascular Disease | 53 | 65 | 43 | 53.67 |
| Sickle Cell Disease | 66 | 73 | 23 | 54.00 |
| Malignant Cancer | 46 | 56 | 64 | 55.33 |
| COVID Treatment: Corticosteroids | 54 | 57 | 61 | 57.33 |
| Myocardial Infarction | 70 | 66 | 38 | 58.00 |
| Age 90+ | 68 | 64 | 48 | 60.00 |
| HIV | 67 | 59 | 55 | 60.33 |
| Mild Liver Disease | 61 | 69 | 60 | 63.33 |
| Race/Ethnicity: Other race NH | 64 | 62 | 65 | 63.67 |
| Cerebrovascular Disease | 69 | 72 | 51 | 64.00 |

**eTable 8. Cohort Characteristics for PASC cases defined by U09.9 or clinic visit for Hospitalized During COVID Index**

|  | PASC (N=3062) | Method 1  Unrestricted Controls  (N=15605) | Method 2  Restricted  controls  (N=15353) | Method 3  Most Restricted  controls  (N=12071) |
| --- | --- | --- | --- | --- |
| **Demographics** |  |  |  |  |
| Age |  |  |  |  |
| 18-29 | 132 (4.3%) | 1800 (11.5%) | 1931 (12.6%) | 2498 (20.7%) |
| 30-39 | 298 (9.7%) | 2140 (13.7%) | 2117 (13.8%) | 2040 (16.9%) |
| 40-49 | 535 (17.5%) | 2279 (14.6%) | 2050 (13.4%) | 1360 (11.3%) |
| 50-59 | 733 (23.9%) | 2780 (17.8%) | 2574 (16.8%) | 1326 (11.0%) |
| 60-69 | 735 (24.0%) | 2915 (18.7%) | 2808 (18.3%) | 1450 (12.0%) |
| 70-79 | 433 (14.1%) | 2146 (13.8%) | 2166 (14.1%) | 1481 (12.3%) |
| 80-89 | 171 (5.6%) | 1253 (8.0%) | 1346 (8.8%) | 1403 (11.6%) |
| 90+ | 24 (0.8%) | 218 (1.4%) | 264 (1.7%) | 390 (3.2%) |
| Sex |  |  |  |  |
| Female | 1627 (53.1%) | 8115 (52.0%) | 8260 (53.8%) | 6143 (50.9%) |
| Male | 1434 (46.8%) | 7488 (48.0%) | 7091 (46.2%) | 5927 (49.1%) |
| Race/Ethnicity |  |  |  |  |
| White non-Hispanic (NH) | 1878 (61.3%) | 8821 (56.5%) | 8982 (58.5%) | 6638 (55.0%) |
| Hispanic | 397 (13.0%) | 2410 (15.4%) | 2096 (13.7%) | 1875 (15.5%) |
| Black NH | 586 (19.1%) | 3161 (20.3%) | 3250 (21.2%) | 2666 (22.1%) |
| Asian NH | 57 (1.9%) | 368 (2.4%) | 357 (2.3%) | 286 (2.4%) |
| Other race NH | 22 (0.7%) | 134 (0.9%) | 101 (0.7%) | 87 (0.7%) |
| **Comorbidities Prior to COVID Index Date** |  |  |  |  |
| AKI | 654 (21.4%) | 2987 (19.1%) | 3049 (19.9%) | 2119 (17.6%) |
| Cardiomyopathies | 147 (4.8%) | 679 (4.4%) | 780 (5.1%) | 457 (3.8%) |
| Cerebrovascular Disease | 226 (7.4%) | 1097 (7.0%) | 1304 (8.5%) | 956 (7.9%) |
| Chronic Lung Disease | 1113 (36.3%) | 3919 (25.1%) | 4008 (26.1%) | 2490 (20.6%) |
| Complicated Diabetes | 737 (24.1%) | 3164 (20.3%) | 3433 (22.4%) | 2139 (17.7%) |
| Congestive Heart Failure | 396 (12.9%) | 1738 (11.1%) | 1943 (12.7%) | 1293 (10.7%) |
| Coronary Artery Disease | 495 (16.2%) | 2153 (13.8%) | 2451 (16.0%) | 1637 (13.6%) |
| Dementia | 77 (2.5%) | 682 (4.4%) | 672 (4.4%) | 670 (5.6%) |
| Down Syndrome | <20 | <20 | <20 | <20 |
| Heart Failure | 490 (16.0%) | 2136 (13.7%) | 2320 (15.1%) | 1554 (12.9%) |
| Hemiplegia or Paraplegia | 36 (1.2%) | 254 (1.6%) | 319 (2.1%) | 220 (1.8%) |
| HIV | 21 (0.7%) | 103 (0.7%) | 156 (1.0%) | 104 (0.9%) |
| Hypertension | 1584 (51.7%) | 6684 (42.8%) | 7438 (48.4%) | 4941 (40.9%) |
| Kidney Disease | 836 (27.3%) | 3912 (25.1%) | 4135 (26.9%) | 2776 (23.0%) |
| Malignant Cancer | 415 (13.6%) | 1802 (11.5%) | 2172 (14.1%) | 1396 (11.6%) |
| Metastatic Solid Tumor Cancers | 55 (1.8%) | 339 (2.2%) | 432 (2.8%) | 239 (2.0%) |
| Mild Liver Disease | 97 (3.2%) | 531 (3.4%) | 626 (4.1%) | 388 (3.2%) |
| Moderate to Severe Liver Disease | 56 (1.8%) | 333 (2.1%) | 356 (2.3%) | 219 (1.8%) |
| Myocardial Infarction | 252 (8.2%) | 1348 (8.6%) | 1517 (9.9%) | 972 (8.1%) |
| Obesity | 2009 (65.6%) | 8284 (53.1%) | 8694 (56.6%) | 6206 (51.4%) |
| Peptic Ulcer | 140 (4.6%) | 503 (3.2%) | 581 (3.8%) | 379 (3.1%) |
| Peripheral Vascular Disease | 262 (8.6%) | 1018 (6.5%) | 1174 (7.6%) | 732 (6.1%) |
| Rheumatologic Disease | 155 (5.1%) | 474 (3.0%) | 528 (3.4%) | 239 (2.0%) |
| Sickle Cell Disease | <20 | 53 (0.3%) | 86 (0.6%) | 72 (0.6%) |
| Systemic Corticosteroids | 1879 (61.4%) | 7743 (49.6%) | 8090 (52.7%) | 5479 (45.4%) |
| Thalassemia | <20 | 39 (0.2%) | 61 (0.4%) | 57 (0.5%) |
| Tuberculosis | <20 | 46 (0.3%) | 67 (0.4%) | 36 (0.3%) |
| Uncomplicated Diabetes | 969 (31.6%) | 4228 (27.1%) | 4526 (29.5%) | 2986 (24.7%) |
| **Behavioral Health Indicators** |  |  |  |  |
| Depression | 742 (24.2%) | 2985 (19.1%) | 3462 (22.5%) | 2313 (19.2%) |
| Psychosis | 42 (1.4%) | 372 (2.4%) | 416 (2.7%) | 353 (2.9%) |
| Substance Abuse | 102 (3.3%) | 1003 (6.4%) | 1162 (7.6%) | 943 (7.8%) |
| Tobacco Smoker | 219 (7.2%) | 1628 (10.4%) | 1769 (11.5%) | 1253 (10.4%) |
| **Characteristics during Acute COVID Phase** |  |  |  |  |
| COVID Diagnosis during COVID-associated Hospitalization | 2889 (94.4%) | 13786 (88.3%) | 13264 (86.4%) | 10324 (85.5%) |
| COVID-associated Hospitalization | 3062 (100.0%) | 15605 (100.0%) | 15353 (100.0%) | 12071 (100.0%) |
| COVID-associated ED Visit | 608 (19.9%) | 2316 (14.8%) | 2586 (16.8%) | 1851 (15.3%) |
| Hospitalization Stay |  |  |  |  |
| Short Stay (0-2 days) | 603 (19.7%) | 5501 (35.3%) | 5447 (35.5%) | 4658 (38.6%) |
| Medium Stay (3-7 days) | 858 (28.0%) | 5430 (34.8%) | 5393 (35.1%) | 4131 (34.2%) |
| Long Stay (8-30 days) | 1017 (33.2%) | 3274 (21.0%) | 3076 (20.0%) | 2172 (18.0%) |
| Extended Stay (31+ days) | 443 (14.5%) | 572 (3.7%) | 461 (3.0%) | 299 (2.5%) |
| COVID Treatment |  |  |  |  |
| Corticosteroids | 2000 (65.3%) | 7734 (49.6%) | 7254 (47.2%) | 4947 (41.0%) |
| Remdesivir | 1390 (45.4%) | 4932 (31.6%) | 4417 (28.8%) | 2954 (24.5%) |
| Vasopressors | 588 (19.2%) | 1774 (11.4%) | 1856 (12.1%) | 1504 (12.5%) |
| ECMO | 67 (2.2%) | 52 (0.3%) | 22 (0.1%) | 21 (0.2%) |
| Mechanical Ventilation | 604 (19.7%) | 1130 (7.2%) | 858 (5.6%) | 551 (4.6%) |
| AKI during COVID-associated Hospitalization | 683 (22.3%) | 2497 (16.0%) | 2393 (15.6%) | 1692 (14.0%) |
| Sepsis during COVID-associated Hospitalization | 615 (20.1%) | 1974 (12.6%) | 1808 (11.8%) | 1248 (10.3%) |

**eTable 9. Cohort Characteristics for PASC cases defined by U09.9 or clinic visit for Not Hospitalized During COVID Index**

|  | PASC  (N=5232) | Method 1  Unrestricted Controls  (N=26245) | Method 2  Restricted  controls  (N=26160) | Method 3  Most Restricted  controls  (N=25616) |
| --- | --- | --- | --- | --- |
| **Demographics** |  |  |  |  |
| Age |  |  |  |  |
| 18-29 | 496 (9.5%) | 5697 (21.7%) | 5275 (20.2%) | 7149 (27.9%) |
| 30-39 | 930 (17.8%) | 5185 (19.8%) | 4832 (18.5%) | 4969 (19.4%) |
| 40-49 | 1207 (23.1%) | 4835 (18.4%) | 4542 (17.4%) | 4154 (16.2%) |
| 50-59 | 1192 (22.8%) | 4453 (17.0%) | 4637 (17.7%) | 3925 (15.3%) |
| 60-69 | 859 (16.4%) | 3389 (12.9%) | 3774 (14.4%) | 2552 (10.0%) |
| 70-79 | 406 (7.8%) | 1895 (7.2%) | 2172 (8.3%) | 1781 (7.0%) |
| 80-89 | 125 (2.4%) | 656 (2.5%) | 786 (3.0%) | 848 (3.3%) |
| 90+ | <20 | 112 (0.4%) | 122 (0.5%) | 195 (0.8%) |
| Sex |  |  |  |  |
| Female | 3581 (68.4%) | 14910 (56.8%) | 15670 (59.9%) | 14397 (56.2%) |
| Male | 1651 (31.6%) | 11294 (43.0%) | 10476 (40.0%) | 11206 (43.7%) |
| Race/Ethnicity |  |  |  |  |
| White non-Hispanic (NH) | 3815 (72.9%) | 17182 (65.5%) | 17877 (68.3%) | 17265 (67.4%) |
| Hispanic | 431 (8.2%) | 2769 (10.6%) | 2546 (9.7%) | 2721 (10.6%) |
| Black NH | 646 (12.3%) | 3651 (13.9%) | 3940 (15.1%) | 3674 (14.3%) |
| Asian NH | 77 (1.5%) | 534 (2.0%) | 582 (2.2%) | 625 (2.4%) |
| Other race NH | 32 (0.6%) | 190 (0.7%) | 181 (0.7%) | 186 (0.7%) |
| **Comorbidities Prior to COVID Index Date** |  |  |  |  |
| AKI | 202 (3.9%) | 758 (2.9%) | 1066 (4.1%) | 776 (3.0%) |
| Cardiomyopathies | 79 (1.5%) | 333 (1.3%) | 470 (1.8%) | 318 (1.2%) |
| Cerebrovascular Disease | 164 (3.1%) | 615 (2.3%) | 860 (3.3%) | 663 (2.6%) |
| Chronic Lung Disease | 1289 (24.6%) | 3203 (12.2%) | 3925 (15.0%) | 3020 (11.8%) |
| Complicated Diabetes | 466 (8.9%) | 1657 (6.3%) | 2317 (8.9%) | 1650 (6.4%) |
| Congestive Heart Failure | 179 (3.4%) | 581 (2.2%) | 854 (3.3%) | 616 (2.4%) |
| Coronary Artery Disease | 338 (6.5%) | 1265 (4.8%) | 1761 (6.7%) | 1324 (5.2%) |
| Dementia | 74 (1.4%) | 243 (0.9%) | 366 (1.4%) | 320 (1.2%) |
| Down Syndrome | <20 | <20 | <20 | <20 |
| Heart Failure | 243 (4.6%) | 776 (3.0%) | 1095 (4.2%) | 798 (3.1%) |
| Hemiplegia or Paraplegia | 23 (0.4%) | 90 (0.3%) | 161 (0.6%) | 110 (0.4%) |
| HIV | 28 (0.5%) | 145 (0.6%) | 206 (0.8%) | 207 (0.8%) |
| Hypertension | 1765 (33.7%) | 6166 (23.5%) | 8258 (31.6%) | 6485 (25.3%) |
| Kidney Disease | 417 (8.0%) | 1478 (5.6%) | 2075 (7.9%) | 1537 (6.0%) |
| Malignant Cancer | 426 (8.1%) | 1635 (6.2%) | 2348 (9.0%) | 1760 (6.9%) |
| Metastatic Solid Tumor Cancers | 35 (0.7%) | 206 (0.8%) | 293 (1.1%) | 215 (0.8%) |
| Mild Liver Disease | 69 (1.3%) | 287 (1.1%) | 422 (1.6%) | 304 (1.2%) |
| Moderate to Severe Liver Disease | 24 (0.5%) | 114 (0.4%) | 179 (0.7%) | 115 (0.4%) |
| Myocardial Infarction | 138 (2.6%) | 552 (2.1%) | 804 (3.1%) | 616 (2.4%) |
| Obesity | 2657 (50.8%) | 9966 (38.0%) | 12238 (46.8%) | 10592 (41.3%) |
| Peptic Ulcer | 139 (2.7%) | 396 (1.5%) | 538 (2.1%) | 445 (1.7%) |
| Peripheral Vascular Disease | 142 (2.7%) | 458 (1.7%) | 683 (2.6%) | 471 (1.8%) |
| Rheumatologic Disease | 190 (3.6%) | 452 (1.7%) | 614 (2.3%) | 430 (1.7%) |
| Sickle Cell Disease | <20 | 20 (0.1%) | 46 (0.2%) | 41 (0.2%) |
| Systemic Corticosteroids | 2431 (46.5%) | 8019 (30.6%) | 10054 (38.4%) | 8365 (32.7%) |
| Thalassemia | <20 | 40 (0.2%) | 65 (0.2%) | 48 (0.2%) |
| Tuberculosis | <20 | 53 (0.2%) | 52 (0.2%) | 41 (0.2%) |
| Uncomplicated Diabetes | 729 (13.9%) | 2821 (10.7%) | 3810 (14.6%) | 2899 (11.3%) |
| **Behavioral Health Indicators** |  |  |  |  |
| Depression | 1314 (25.1%) | 3376 (12.9%) | 4622 (17.7%) | 3688 (14.4%) |
| Psychosis | 23 (0.4%) | 155 (0.6%) | 229 (0.9%) | 194 (0.8%) |
| Substance Abuse | 101 (1.9%) | 628 (2.4%) | 804 (3.1%) | 699 (2.7%) |
| Tobacco Smoker | 297 (5.7%) | 1452 (5.5%) | 1751 (6.7%) | 1577 (6.2%) |
| **Characteristics during Acute COVID Phase** |  |  |  |  |
| COVID-associated ED Visit | 945 (18.1%) | 4021 (15.3%) | 3559 (13.6%) | 3633 (14.2%) |

**eTable 10. PASC Risk Factors from Logistic Regression (PASC defined as U09.9 or long-COVID visit) Hospitalized during COVID Index**

|  | Method 1  Unrestricted Controls  (N=28473) | Method 2  Restricted  controls  (N=28408) | Method 3  Most Restricted  controls  (N=27817) |
| --- | --- | --- | --- |
| **Demographics** |  |  |  |
| Age |  |  |  |
| 18-29 | REF | REF | REF |
| 30-39 | 1.91 (1.69-2.17) | 2.1 (1.85-2.38) | 2.73 (2.41-3.09) |
| 40-49 | 2.57 (2.28-2.9) | 2.98 (2.64-3.36) | 4.37 (3.87-4.94) |
| 50-59 | 2.67 (2.36-3.02) | 2.97 (2.62-3.36) | 4.6 (4.06-5.22) |
| 60-69 | 2.47 (2.16-2.82) | 2.66 (2.33-3.04) | 5.2 (4.54-5.97) |
| 70-79 | 1.89 (1.61-2.23) | 2.09 (1.78-2.45) | 3.13 (2.65-3.69) |
| 80-89 | 1.66 (1.3-2.11) | 1.82 (1.43-2.3) | 1.97 (1.55-2.52) |
| 90+ | 0.94 (0.5-1.76) | 1.13 (0.61-2.1) | 0.78 (0.42-1.45) |
| Sex |  |  |  |
| Male or Unknown Sex | REF | REF | REF |
| Female | 1.44 (1.35-1.55) | 1.34 (1.25-1.43) | 1.71 (1.59-1.84) |
| Race/Ethnicity |  |  |  |
| White NH | REF | REF | REF |
| Hispanic | 0.82 (0.72-0.92) | 0.84 (0.75-0.95) | 0.84 (0.75-0.95) |
| Black NH | 0.71 (0.64-0.79) | 0.69 (0.62-0.76) | 0.68 (0.61-0.76) |
| Asian NH | 0.77 (0.59-0.99) | 0.62 (0.48-0.8) | 0.6 (0.46-0.79) |
| Other race NH | 1.02 (0.66-1.59) | 1.06 (0.68-1.64) | 1.28 (0.81-2.02) |
| **Comorbidities Prior to COVID Index Date** |  |  |  |
| AKI | 0.83 (0.65-1.05) | 0.91 (0.72-1.14) | 0.78 (0.61-1.0) |
| Cardiomyopathies | 0.74 (0.54-1.0) | 0.68 (0.51-0.91) | 0.82 (0.59-1.12) |
| Cerebrovascular Disease | 0.91 (0.74-1.11) | 0.9 (0.74-1.1) | 0.91 (0.74-1.13) |
| Chronic Lung Disease | 1.78 (1.63-1.93) | 1.77 (1.63-1.92) | 2.03 (1.86-2.21) |
| Complicated Diabetes | 1.12 (0.93-1.34) | 1.14 (0.96-1.36) | 1.2 (0.99-1.44) |
| Congestive Heart Failure | 1.0 (0.69-1.45) | 0.93 (0.65-1.32) | 1.06 (0.71-1.57) |
| Coronary Artery Disease | 1.04 (0.88-1.22) | 0.96 (0.83-1.13) | 1.05 (0.89-1.23) |
| Dementia | 1.19 (0.87-1.61) | 0.98 (0.74-1.31) | 0.96 (0.72-1.3) |
| Down Syndrome | 2.28 (0.19-27.23) | 4.38 (0.27-72.1) | 1.22 (0.13-11.18) |
| Heart Failure | 1.16 (0.84-1.6) | 1.21 (0.89-1.65) | 1.15 (0.82-1.63) |
| Hemiplegia or Paraplegia | 0.84 (0.5-1.42) | 0.64 (0.4-1.05) | 0.64 (0.38-1.07) |
| HIV | 0.84 (0.53-1.32) | 0.64 (0.41-1.01) | 0.6 (0.38-0.95) |
| Hypertension | 1.14 (1.05-1.24) | 0.95 (0.88-1.04) | 1.0 (0.91-1.09) |
| Kidney Disease | 1.14 (0.95-1.36) | 1.05 (0.89-1.25) | 1.18 (0.98-1.41) |
| Malignant Cancer | 0.97 (0.85-1.1) | 0.82 (0.72-0.93) | 0.94 (0.82-1.07) |
| Metastatic Solid Tumor Cancers | 0.57 (0.38-0.85) | 0.6 (0.41-0.89) | 0.6 (0.4-0.89) |
| Mild Liver Disease | 1.0 (0.72-1.39) | 0.86 (0.63-1.18) | 0.97 (0.69-1.35) |
| Moderate to Severe Liver Disease | 0.64 (0.36-1.11) | 0.63 (0.37-1.07) | 0.59 (0.33-1.06) |
| Myocardial Infarction | 0.77 (0.6-0.97) | 0.78 (0.62-0.98) | 0.66 (0.52-0.85) |
| Obesity | 1.18 (1.1-1.27) | 0.97 (0.91-1.04) | 1.06 (0.99-1.14) |
| Peptic Ulcer | 1.23 (0.99-1.53) | 1.15 (0.94-1.42) | 1.11 (0.89-1.39) |
| Peripheral Vascular Disease | 1.16 (0.93-1.45) | 1.06 (0.86-1.31) | 1.27 (1.02-1.6) |
| Rheumatologic Disease | 1.24 (1.03-1.5) | 1.21 (1.01-1.44) | 1.4 (1.15-1.7) |
| Sickle Cell Disease | 0.7 (0.19-2.54) | 0.4 (0.12-1.36) | 0.47 (0.14-1.66) |
| Systemic Corticosteroids | 1.35 (1.26-1.45) | 1.14 (1.07-1.22) | 1.3 (1.21-1.39) |
| Thalassemia | 1.92 (1.01-3.66) | 1.5 (0.83-2.72) | 1.73 (0.92-3.25) |
| Tuberculosis | 0.99 (0.52-1.9) | 1.31 (0.68-2.51) | 1.85 (0.92-3.73) |
| Uncomplicated Diabetes | 0.79 (0.69-0.92) | 0.77 (0.67-0.89) | 0.8 (0.69-0.93) |
| **Behavioral Health** |  |  |  |
| Depression | 1.67 (1.54-1.82) | 1.41 (1.3-1.52) | 1.63 (1.5-1.77) |
| Psychosis | 0.55 (0.33-0.91) | 0.52 (0.32-0.86) | 0.47 (0.28-0.78) |
| Substance Abuse | 0.61 (0.48-0.78) | 0.61 (0.48-0.77) | 0.64 (0.5-0.83) |
| Tobacco Smoker | 0.78 (0.67-0.9) | 0.75 (0.65-0.86) | 0.72 (0.63-0.84) |
| **Characteristics of Index COVID "Acute Phase"** |  |  |  |
| COVID-associated ED Visit | 1.2 (1.1-1.31) | 1.46 (1.34-1.6) | 1.43 (1.3-1.56) |
| **Social Determinants of Health** |  |  |  |
| Households with Income below poverty: medium (11-15%) | 0.87 (0.79-0.94) | 0.77 (0.7-0.84) | 0.76 (0.69-0.83) |
| Households with Income below poverty: high (>15%) | 0.92 (0.84-1.0) | 0.94 (0.86-1.03) | 0.9 (0.82-0.99) |
| College Degree medium (19-25%) | 0.97 (0.86-1.09) | 0.98 (0.88-1.1) | 0.92 (0.82-1.04) |
| College Degree high (>25%) | 1.0 (0.87-1.14) | 0.97 (0.85-1.11) | 0.93 (0.81-1.08) |
| Public health Insurance for ages 19-64: Medium (13-18%) | 1.04 (0.96-1.12) | 1.05 (0.98-1.14) | 1.06 (0.98-1.15) |
| Public health Insurance for ages 19-64: High (>18%) | 0.94 (0.86-1.03) | 0.9 (0.83-0.99) | 0.9 (0.82-0.99) |
| MDs per 1000 residents: medium (1.91-3.61%) | 1.22 (1.08-1.36) | 1.15 (1.03-1.29) | 1.22 (1.08-1.38) |
| MDs per 1000 residents: High (>3.61%) | 1.18 (1.04-1.33) | 1.04 (0.92-1.18) | 1.18 (1.04-1.34) |

**eTable 11. PASC Risk Factors from Logistic Regression including SDoH (PASC defined as U09.9 or long-COVID visit) Not Hospitalized during COVID Index**

|  | Method 1  Unrestricted Controls  (N=16394) | Method 2  Restricted  controls  (N=16110) | Method 3  Most Restricted  controls  (N=12395) |
| --- | --- | --- | --- |
| **Demographics** |  |  |  |
| Age |  |  |  |
| 18-29 | REF | REF | REF |
| 30-39 | 1.73 (1.37-2.18) | 1.91 (1.51-2.42) | 2.76 (2.1-3.64) |
| 40-49 | 2.44 (1.96-3.05) | 3.07 (2.46-3.85) | 6.82 (5.24-8.89) |
| 50-59 | 2.58 (2.07-3.2) | 3.19 (2.56-3.97) | 8.8 (6.77-11.43) |
| 60-69 | 2.23 (1.79-2.79) | 2.77 (2.22-3.46) | 7.02 (5.38-9.16) |
| 70-79 | 1.76 (1.39-2.23) | 2.22 (1.74-2.82) | 3.86 (2.92-5.11) |
| 80-89 | 1.24 (0.94-1.64) | 1.38 (1.03-1.84) | 1.53 (1.1-2.12) |
| 90+ | 1.08 (0.64-1.81) | 0.98 (0.58-1.66) | 0.79 (0.44-1.41) |
| Sex |  |  |  |
| Male or Unknown Sex | REF | REF | REF |
| Female | 1.07 (1.0-1.15) | 1.06 (0.96-1.16) | 1.56 (1.39-1.75) |
| Race/Ethnicity |  |  |  |
| White NH | REF | REF | REF |
| Hispanic | 0.75 (0.65-0.86) | 0.88 (0.77-1.01) | 0.81 (0.68-0.96) |
| Black NH | 0.91 (0.81-1.03) | 0.9 (0.8-1.01) | 0.82 (0.7-0.95) |
| Asian NH | 0.79 (0.58-1.08) | 0.77 (0.56-1.05) | 0.68 (0.47-1.0) |
| Other race NH | 0.92 (0.54-1.58) | 1.3 (0.75-2.26) | 0.74 (0.34-1.6) |
| **Comorbidities Prior to COVID Index Date** |  |  |  |
| AKI | 0.89 (0.72-1.1) | 0.94 (0.76-1.15) | 0.84 (0.64-1.1) |
| Cardiomyopathies | 0.94 (0.75-1.19) | 0.93 (0.74-1.17) | 1.2 (0.9-1.58) |
| Cerebrovascular Disease | 1.07 (0.89-1.29) | 0.9 (0.75-1.08) | 0.81 (0.65-1.02) |
| Chronic Lung Disease | 1.22 (1.1-1.35) | 1.29 (1.16-1.42) | 1.57 (1.38-1.79) |
| Complicated Diabetes | 0.86 (0.72-1.03) | 0.88 (0.73-1.05) | 0.91 (0.73-1.14) |
| Congestive Heart Failure | 1.02 (0.77-1.36) | 0.95 (0.71-1.27) | 1.03 (0.72-1.5) |
| Coronary Artery Disease | 1.17 (1.01-1.36) | 1.17 (1.01-1.36) | 1.28 (1.07-1.54) |
| Dementia | 0.61 (0.46-0.81) | 0.68 (0.51-0.9) | 0.58 (0.42-0.81) |
| Down Syndrome | 2.64 (0.55-12.72) | 0.47 (0.08-2.68) | 0.28 (0.02-4.3) |
| Heart Failure | 0.99 (0.76-1.28) | 1.06 (0.81-1.38) | 0.94 (0.67-1.32) |
| Hemiplegia or Paraplegia | 0.48 (0.31-0.75) | 0.38 (0.24-0.59) | 0.28 (0.16-0.49) |
| HIV | 1.04 (0.61-1.76) | 0.7 (0.4-1.2) | 0.78 (0.41-1.49) |
| Hypertension | 1.19 (1.07-1.32) | 0.98 (0.88-1.09) | 1.1 (0.97-1.26) |
| Kidney Disease | 0.87 (0.72-1.04) | 0.84 (0.7-1.0) | 0.98 (0.78-1.23) |
| Malignant Cancer | 1.12 (0.97-1.3) | 1.02 (0.88-1.18) | 1.18 (0.99-1.4) |
| Metastatic Solid Tumor Cancers | 0.72 (0.51-1.01) | 0.67 (0.47-0.93) | 0.69 (0.45-1.04) |
| Mild Liver Disease | 0.99 (0.75-1.3) | 0.88 (0.68-1.15) | 0.94 (0.67-1.32) |
| Moderate to Severe Liver Disease | 0.65 (0.45-0.93) | 0.68 (0.47-0.99) | 0.63 (0.4-1.0) |
| Myocardial Infarction | 0.79 (0.66-0.94) | 0.74 (0.61-0.88) | 0.63 (0.5-0.78) |
| Obesity | 1.34 (1.21-1.47) | 1.22 (1.1-1.35) | 1.28 (1.13-1.44) |
| Peptic Ulcer | 1.26 (1.01-1.58) | 1.26 (1.01-1.58) | 1.11 (0.83-1.49) |
| Peripheral Vascular Disease | 1.17 (0.98-1.4) | 1.18 (0.98-1.41) | 1.37 (1.1-1.71) |
| Rheumatologic Disease | 1.44 (1.16-1.79) | 1.33 (1.08-1.65) | 1.83 (1.38-2.43) |
| Sickle Cell Disease | 2.56 (1.28-5.12) | 1.74 (0.89-3.38) | 1.85 (0.85-4.0) |
| Systemic Corticosteroids | 1.22 (1.14-1.31) | 1.17 (1.06-1.29) | 1.23 (1.09-1.39) |
| Thalassemia | 0.39 (0.13-1.2) | 0.5 (0.17-1.49) | 0.43 (0.12-1.52) |
| Tuberculosis | 1.74 (0.86-3.52) | 1.21 (0.62-2.37) | 1.71 (0.75-3.86) |
| Uncomplicated Diabetes | 1.0 (0.85-1.18) | 0.97 (0.82-1.14) | 1.0 (0.82-1.22) |
| **Behavioral Health** |  |  |  |
| Depression | 1.26 (1.12-1.41) | 1.22 (1.09-1.36) | 1.38 (1.2-1.59) |
| Psychosis | 0.57 (0.38-0.83) | 0.53 (0.36-0.79) | 0.43 (0.27-0.69) |
| Substance Abuse | 0.62 (0.49-0.79) | 0.53 (0.41-0.68) | 0.4 (0.3-0.55) |
| Tobacco Smoker | 0.68 (0.57-0.8) | 0.69 (0.58-0.82) | 0.62 (0.49-0.77) |
| **Characteristics of Index COVID "Acute Phase"** |  |  |  |
| COVID Diagnosis during COVID-associated Hospitalization | 1.81 (1.48-2.21) | 2.07 (1.73-2.46) | 1.94 (1.54-2.44) |
| COVID-associated Hospitalization | 0.19 (nan-nan) | 0.19 (nan-nan) | 0.12 (0.0-inf) |
| COVID-associated ED Visit | 1.77 (1.58-1.99) | 1.52 (1.36-1.71) | 1.69 (1.46-1.95) |
| Hospitalization Stay |  |  |  |
| Not Hospitalized | REF | REF | REF |
| Hospitalization Short Stay (0-2 days) | 0.58 (0.45-0.75) | 0.58 (0.44-0.76) | 0.51 (0.38-0.68) |
| Hospitalization Medium Stay (3-7 days) | 0.69 (0.53-0.9) | 0.65 (0.5-0.86) | 0.56 (0.42-0.76) |
| Hospitalization Long Stay (8-30 days) | 1.33 (1.02-1.74) | 1.29 (0.99-1.7) | 1.26 (0.93-1.71) |
| Hospitalization Extended Stay (31+ days) | 2.66 (1.94-3.64) | 2.81 (2.04-3.88) | 3.03 (2.07-4.43) |
| COVID Treatment |  |  |  |
| Corticosteroids | 1.08 (0.96-1.22) | 1.17 (1.05-1.31) | 1.21 (1.05-1.4) |
| Remdesivir | 1.14 (1.02-1.26) | 1.25 (1.12-1.4) | 1.38 (1.2-1.58) |
| ECMO | 2.28 (1.48-3.54) | 4.0 (2.29-6.99) | 2.54 (1.3-4.96) |
| Mechanical Ventilation | 1.43 (1.21-1.68) | 1.77 (1.56-2.02) | 1.68 (1.35-2.1) |
| Vasopressors | 0.8 (0.73-0.88) | 0.94 (0.81-1.09) | 0.76 (0.63-0.92) |
| AKI during COVID-associated Hospitalization | 1.08 (0.92-1.27) | 1.09 (0.93-1.29) | 1.06 (0.86-1.3) |
| Sepsis during COVID-associated Hospitalization | 0.94 (0.82-1.06) | 0.95 (0.87-1.04) | 0.93 (0.79-1.1) |
| **Social Determinants of Health** |  |  |  |
| Households with Income below poverty: medium (11-15%) | 1.05 (0.92-1.19) | 0.99 (0.85-1.16) | 1.0 (0.86-1.17) |
| Households with Income below poverty: high (>15%) | 1.0 (0.88-1.14) | 1.06 (0.97-1.15) | 1.07 (0.92-1.25) |
| College Degree medium (19-25%) | 0.86 (0.75-0.98) | 0.91 (0.75-1.11) | 0.88 (0.74-1.06) |
| College Degree high (>25%) | 0.84 (0.7-1.01) | 0.83 (0.69-0.99) | 0.77 (0.62-0.96) |
| Public health Insurance for ages 19-64: Medium (13-18%) | 0.9 (0.82-0.99) | 0.89 (0.8-0.99) | 0.84 (0.74-0.95) |
| Public health Insurance for ages 19-64: High (>18%) | 0.87 (0.77-0.98) | 0.84 (0.71-0.99) | 0.8 (0.69-0.92) |
| MDs per 1000 residents: medium (1.91-3.61%) | 1.26 (1.11-1.44) | 1.16 (1.07-1.27) | 1.32 (1.09-1.6) |
| MDs per 1000 residents: High (>3.61%) | 1.54 (1.35-1.76) | 1.36 (nan-nan) | 1.53 (1.27-1.84) |

**eTable 12. Comparison of Feature Importance Across Models for Hospitalized during Index COVID for Unrestricted Sample including SDOH variables (PASC defined as U09.9 or Long-COVID Clinic Visit)**

| features | Random Forest | XGBoost | Logistic Regression | Mean Rank |
| --- | --- | --- | --- | --- |
| Hospitalization Extended Stay (31+ days**)** | 1 | 1 | 2 | 1.33 |
| COVID-associated ED Visit | 3 | 2 | 14 | 6.33 |
| ECMO during COVID-associated Hospitalization | 7 | 12 | 8 | 9.00 |
| Age 50-59 | 15 | 8 | 4 | 9.00 |
| Mechanical Ventilation during COVID-associated Hospitalization | 2 | 4 | 24 | 10.00 |
| MDs per 1000 residents: High (>3.61%) | 14 | 6 | 20 | 13.33 |
| Age 40-49 | 37 | 5 | 7 | 16.33 |
| COVID Diagnosis during COVID-associated Hospitalization | 22 | 11 | 17 | 16.67 |
| Hospitalization Long Stay | 25 | 24 | 10 | 19.67 |
| Obesity | 27 | 10 | 26 | 21.00 |
| Race/Ethnicity: White non-Hispanic (NH) | 12 | 31 | ref. | 21.50 |
| Age 60-69 | 11 | 47 | 9 | 22.33 |
| College Degree medium (19-25%) | 4 | 30 | 38 | 24.00 |
| Chronic Lung Disease | 9 | 49 | 35 | 31.00 |
| Peripheral Vascular Disease | 35 | 23 | 40 | 32.67 |
| Female | 13 | 35 | 52 | 33.33 |
| AKI during COVID-associated Hospitalization | 33 | 17 | 53 | 34.33 |
| Depression | 55 | 26 | 32 | 37.67 |
| Received COVID Regimen Corticosteroids | 26 | 34 | 54 | 38.00 |
| Received Remdisivir | 66 | 9 | 44 | 39.67 |
| Malignant Cancer | 40 | 37 | 45 | 40.67 |
| Sickle Cell Disease | 64 | 60 | 6 | 43.33 |
| Hypertension | 49 | 46 | 37 | 44.00 |
| Cerebrovascular Disease | 57 | 21 | 55 | 44.33 |
| Coronary Artery Disease | 38 | 57 | 39 | 44.67 |
| Down Syndrome | 70 | 68 | 3 | 47.00 |
| Households with Income below poverty: medium (11-15%) | 19 | 63 | 60 | 47.33 |
| Peptic Ulcer | 48 | 67 | 31 | 48.67 |
| Tuberculosis | 71 | 59 | 16 | 48.67 |
| Systemic Corticosteroids | 60 | 69 | 34 | 54.33 |
| Rheumatologic Disease | 72 | 70 | 23 | 55.00 |
| Heart Failure | 46 | 58 | 64 | 56.00 |
| Congestive Heart Failure | 45 | 71 | 61 | 59.00 |
| HIV | 65 | 66 | 59 | 63.33 |
| COVID-associated Hospitalization | 73 | 73 | ref. | 73.00 |
| College Degree low (<19%) | 16 | 29 | ref. | 22.50 |
| Public health Insurance for ages 19-64: Low (<13%) | 24 | 22 | ref. | 23.00 |
| Households with Income below poverty: low (<11%) | 23 | 38 | ref. | 30.50 |
| MDs per 1000 residents: Low (<1.91%) | 6 | 14 | ref. | 10.00 |
| Hospitalization Short Stay (0-2 days) | 32 | 3 | ref. | 17.50 |
| Substance Abuse | 31 | 7 | 19 | 19.00 |
| MDs per 1000 residents: medium (1.91-3.61%) | 5 | 27 | 30 | 20.67 |
| Age 70-79 | 36 | 25 | 13 | 24.67 |
| Tobacco Smoker | 43 | 16 | 22 | 27.00 |
| Age 30-39 | 28 | 43 | 15 | 28.67 |
| College Degree high (>25%) | 17 | 36 | 36 | 29.67 |
| Public health Insurance for ages 19-64: High (>18%) | 20 | 28 | 43 | 30.33 |
| Hospitalization Medium Stay (3-7 days) | 41 | 13 | 46 | 33.33 |
| Public health Insurance for ages 19-64: Medium (13-18%) | 8 | 44 | 48 | 33.33 |
| Kidney Disease | 47 | 15 | 42 | 34.67 |
| Age 80-89 | 39 | 33 | 33 | 35.00 |
| AKI | 42 | 18 | 47 | 35.67 |
| Race/Ethnicity: Hispanic | 29 | 52 | 27 | 36.00 |
| Male | 10 | 62 | ref. | 36.00 |
| Age 18-29 | 54 | 19 | ref. | 36.50 |
| Hemiplegia or Paraplegia | 59 | 40 | 11 | 36.67 |
| Dementia | 52 | 42 | 18 | 37.33 |
| Received Vasopressors | 30 | 32 | 58 | 40.00 |
| Households with Income below poverty: high (>15%) | 18 | 41 | 62 | 40.33 |
| Complicated Diabetes | 34 | 48 | 41 | 41.00 |
| Psychosis | 56 | 55 | 12 | 41.00 |
| Thalassemia | 68 | 53 | 5 | 42.00 |
| Race/Ethnicity: Black NH | 21 | 61 | 49 | 43.67 |
| Moderate to Severe Liver Disease | 62 | 50 | 21 | 44.33 |
| Sepsis during COVID-associated Hospitalization | 44 | 39 | 56 | 46.33 |
| Cardiomyopathies | 63 | 20 | 57 | 46.67 |
| Myocardial Infarction | 61 | 54 | 29 | 48.00 |
| Metastatic Solid Tumor Cancers | 67 | 56 | 25 | 49.33 |
| Race/Ethnicity: Asian NH | 53 | 72 | 28 | 51.00 |
| Mild Liver Disease | 58 | 45 | 63 | 55.33 |
| Age 90+ | 50 | 65 | 51 | 55.33 |
| Uncomplicated Diabetes | 51 | 51 | 65 | 55.67 |
| Race/Ethnicity: Other race NH | 69 | 64 | 50 | 61.00 |

**eTable 13. Comparison of Feature Importance Across Models for Not-Hospitalized during Index COVID for Unrestricted Sample including SDOH variables**

| features | Random Forest | XGBoost | Logistic Regression | Mean Rank |
| --- | --- | --- | --- | --- |
| Households with Income below poverty: low (<11%) | 5 | 4 | ref. | 4.50 |
| Female | 1 | 1 | 16 | 6.00 |
| Age 40-49 | 11 | 23 | 3 | 12.33 |
| Age 60-69 | 18 | 16 | 4 | 12.67 |
| Chronic Lung Disease | 23 | 8 | 10 | 13.67 |
| Systemic Corticosteroids | 24 | 6 | 20 | 16.67 |
| Age 50-59 | 13 | 36 | 2 | 17.00 |
| Depression | 40 | 2 | 12 | 18.00 |
| MDs per 1000 residents: High (>3.61%) | 3 | 31 | 35 | 23.00 |
| Race/Ethnicity: White non-Hispanic (NH) | 19 | 28 | ref. | 23.50 |
| Peptic Ulcer | 34 | 15 | 26 | 25.00 |
| College Degree high (>25%) | 12 | 11 | 52 | 25.00 |
| Peripheral Vascular Disease | 42 | 7 | 36 | 28.33 |
| MDs per 1000 residents: medium (1.91-3.61%) | 8 | 50 | 28 | 28.67 |
| Coronary Artery Disease | 28 | 12 | 47 | 29.00 |
| Rheumatologic Disease | 57 | 9 | 25 | 30.33 |
| Dementia | 49 | 10 | 33 | 30.67 |
| Thalassemia | 54 | 39 | 6 | 33.00 |
| COVID-associated ED Visit | 21 | 53 | 30 | 34.67 |
| Hypertension | 22 | 44 | 39 | 35.00 |
| Complicated Diabetes | 27 | 38 | 41 | 35.33 |
| Obesity | 46 | 27 | 34 | 35.67 |
| Kidney Disease | 41 | 26 | 40 | 35.67 |
| Public health Insurance for ages 19-64: Medium (13-18%) | 15 | 51 | 46 | 37.33 |
| Heart Failure | 35 | 47 | 37 | 39.67 |
| Congestive Heart Failure | 47 | 20 | 54 | 40.33 |
| Down Syndrome | 60 | 60 | 5 | 41.67 |
| Mild Liver Disease | 43 | 41 | 53 | 45.67 |
| Race/Ethnicity: Other race NH | 50 | 42 | 50 | 47.33 |
| Tuberculosis | 56 | 55 | 51 | 54.00 |
| Male | 2 | 5 | ref. | 3.50 |
| MDs per 1000 residents: Low (<1.91%) | 9 | 13 | ref. | 11.00 |
| Uncomplicated Diabetes | 29 | 3 | 24 | 18.67 |
| Age 30-39 | 17 | 37 | 7 | 20.33 |
| Race/Ethnicity: Black NH | 20 | 24 | 18 | 20.67 |
| Households with Income below poverty: medium (11-15%) | 6 | 25 | 38 | 23.00 |
| Psychosis | 48 | 14 | 9 | 23.67 |
| Tobacco Smoker | 32 | 17 | 23 | 24.00 |
| Race/Ethnicity: Hispanic | 30 | 18 | 27 | 25.00 |
| Age 70-79 | 25 | 45 | 8 | 26.00 |
| College Degree medium (19-25%) | 4 | 32 | 49 | 28.33 |
| College Degree low (<19%) | 10 | 49 | ref. | 29.50 |
| Households with Income below poverty: high (>15%) | 7 | 40 | 43 | 30.00 |
| Metastatic Solid Tumor Cancers | 33 | 48 | 11 | 30.67 |
| Moderate to Severe Liver Disease | 53 | 29 | 15 | 32.33 |
| Substance Abuse | 51 | 34 | 14 | 33.00 |
| Hemiplegia or Paraplegia | 45 | 22 | 32 | 33.00 |
| Cardiomyopathies | 59 | 21 | 19 | 33.00 |
| Cerebrovascular Disease | 39 | 19 | 42 | 33.33 |
| AKI | 37 | 35 | 29 | 33.67 |
| Public health Insurance for ages 19-64: High (>18%) | 26 | 30 | 45 | 33.67 |
| Age 80-89 | 44 | 46 | 13 | 34.33 |
| Public health Insurance for ages 19-64: Low (<13%) | 14 | 57 | ref. | 35.50 |
| Age 18-29 | 16 | 56 | ref. | 36.00 |
| Race/Ethnicity: Asian NH | 36 | 52 | 22 | 36.67 |
| Malignant Cancer | 31 | 33 | 48 | 37.33 |
| Myocardial Infarction | 38 | 54 | 21 | 37.67 |
| Sickle Cell Disease | 58 | 58 | 17 | 44.33 |
| Age 90+ | 52 | 43 | 44 | 46.33 |
| HIV | 55 | 59 | 31 | 48.33 |

**Other Definitions of Long-COVID**

**eTable 14. Characteristics of Cohorts for U09.9 only**

|  | U09.9 Only  (N=7512) | Method 1  Unrestricted controls (N=37575) | Method 2  Restricted controls (N=37560) | Method 3  Most restricted  controls (N=37560) |
| --- | --- | --- | --- | --- |
| **Demographics** |  |  |  |  |
| Age |  |  |  |  |
| Age 18-29 | 577 (7.7%) | 7633 (20.3%) | 7488 (19.9%) | 6890 (18.3%) |
| Age 30-39 | 1090 (14.5%) | 7206 (19.2%) | 7030 (18.7%) | 6681 (17.8%) |
| Age 40-49 | 1580 (21.0%) | 6698 (17.8%) | 6246 (16.6%) | 6482 (17.3%) |
| Age 50-59 | 1738 (23.1%) | 6235 (16.6%) | 6346 (16.9%) | 6534 (17.4%) |
| Age 60-69 | 1450 (19.3%) | 5235 (13.9%) | 5269 (14.0%) | 5838 (15.5%) |
| Age 70-79 | 766 (10.2%) | 3094 (8.2%) | 3450 (9.2%) | 3457 (9.2%) |
| Age 80-89 | 268 (3.6%) | 1220 (3.2%) | 1425 (3.8%) | 1393 (3.7%) |
| Age 90+ | 43 (0.6%) | 254 (0.7%) | 306 (0.8%) | 285 (0.8%) |
| Sex |  |  |  |  |
| Female | 4752 (63.3%) | 20826 (55.4%) | 21974 (58.5%) | 22483 (59.9%) |
| Male | 2759 (36.7%) | 16694 (44.4%) | 15567 (41.4%) | 15052 (40.1%) |
| Race |  |  |  |  |
| White non-Hispanic (NH) | 5301 (70.6%) | 24371 (64.9%) | 25307 (67.4%) | 25573 (68.1%) |
| Hispanic NH | 711 (9.5%) | 4217 (11.2%) | 3936 (10.5%) | 3783 (10.1%) |
| Black NH | 1035 (13.8%) | 5330 (14.2%) | 5771 (15.4%) | 5599 (14.9%) |
| Asian NH | 116 (1.5%) | 770 (2.0%) | 779 (2.1%) | 749 (2.0%) |
| Other race NH | 52 (0.7%) | 299 (0.8%) | 261 (0.7%) | 263 (0.7%) |
| **Comorbidities Prior to COVID Index Date** |  |  |  |  |
| AKI | 814 (10.8%) | 2056 (5.5%) | 2638 (7.0%) | 2626 (7.0%) |
| Cardiomyopathies | 208 (2.8%) | 648 (1.7%) | 933 (2.5%) | 913 (2.4%) |
| Cerebrovascular Disease | 364 (4.8%) | 1226 (3.3%) | 1718 (4.6%) | 1682 (4.5%) |
| Chronic Lung Disease | 2245 (29.9%) | 5248 (14.0%) | 6586 (17.5%) | 6642 (17.7%) |
| Complicated Diabetes | 1136 (15.1%) | 3276 (8.7%) | 4210 (11.2%) | 4285 (11.4%) |
| Congestive Heart Failure | 522 (6.9%) | 1424 (3.8%) | 1875 (5.0%) | 1886 (5.0%) |
| Coronary Artery Disease | 778 (10.4%) | 2459 (6.5%) | 3198 (8.5%) | 3246 (8.6%) |
| Dementia | 145 (1.9%) | 604 (1.6%) | 772 (2.1%) | 689 (1.8%) |
| Down Syndrome | <20 | <20 | <20 | <20 |
| Heart Failure | 669 (8.9%) | 1757 (4.7%) | 2310 (6.2%) | 2334 (6.2%) |
| Hemiplegia or Paraplegia | 58 (0.8%) | 235 (0.6%) | 379 (1.0%) | 309 (0.8%) |
| HIV | 41 (0.5%) | 203 (0.5%) | 262 (0.7%) | 289 (0.8%) |
| Hypertension | 3145 (41.9%) | 10101 (26.9%) | 12844 (34.2%) | 13026 (34.7%) |
| Kidney Disease | 1183 (15.7%) | 3313 (8.8%) | 4240 (11.3%) | 4250 (11.3%) |
| Malignant Cancer | 783 (10.4%) | 2588 (6.9%) | 3642 (9.7%) | 3717 (9.9%) |
| Metastatic Solid Tumor Cancers | 87 (1.2%) | 363 (1.0%) | 515 (1.4%) | 537 (1.4%) |
| Mild Liver Disease | 159 (2.1%) | 564 (1.5%) | 790 (2.1%) | 775 (2.1%) |
| Moderate to Severe Liver Disease | 77 (1.0%) | 259 (0.7%) | 380 (1.0%) | 371 (1.0%) |
| Myocardial Infarction | 371 (4.9%) | 1272 (3.4%) | 1630 (4.3%) | 1542 (4.1%) |
| Obesity | 4279 (57.0%) | 14822 (39.4%) | 17954 (47.8%) | 18008 (47.9%) |
| Peptic Ulcer | 270 (3.6%) | 712 (1.9%) | 958 (2.6%) | 934 (2.5%) |
| Perpheral Vascular Disease | 388 (5.2%) | 960 (2.6%) | 1395 (3.7%) | 1375 (3.7%) |
| Rheumatologic Disease | 334 (4.4%) | 749 (2.0%) | 945 (2.5%) | 1027 (2.7%) |
| Sickle Cell Disease | <20 | 42 (0.1%) | 99 (0.3%) | 85 (0.2%) |
| Systemic Corticosteroids | 4015 (53.4%) | 12627 (33.6%) | 15483 (41.2%) | 15526 (41.3%) |
| Thalassemia | <20 | 48 (0.1%) | 102 (0.3%) | 89 (0.2%) |
| Tuberculosis | 24 (0.3%) | 72 (0.2%) | 105 (0.3%) | 87 (0.2%) |
| Uncomplicated Diabetes | 1589 (21.2%) | 5088 (13.5%) | 6359 (16.9%) | 6452 (17.2%) |
| **Behavioral Health** |  |  |  |  |
| Depression | 1946 (25.9%) | 5395 (14.4%) | 7257 (19.3%) | 7430 (19.8%) |
| Psychosis | 61 (0.8%) | 350 (0.9%) | 460 (1.2%) | 435 (1.2%) |
| Substance Abuse | 196 (2.6%) | 1165 (3.1%) | 1592 (4.2%) | 1424 (3.8%) |
| Tobacco Smoker | 504 (6.7%) | 2515 (6.7%) | 3143 (8.4%) | 3052 (8.1%) |
| **Characteristics of Index COVID "Acute Phase"** |  |  |  |  |
| COVID Diagnosis during COVID-associated Hospitalization | 1757 (23.4%) | 4136 (11.0%) | 4135 (11.0%) | 3971 (10.6%) |
| COVID-associated Hospitalization | 2683 (35.7%) | 5382 (14.3%) | 5680 (15.1%) | 5436 (14.5%) |
| COVID-associated ED Visit | 1479 (19.7%) | 5858 (15.6%) | 5412 (14.4%) | 5264 (14.0%) |
| Hospitalization stay |  |  |  |  |
| Short Stay (0-2 days) | 560 (7.5%) | 1860 (5.0%) | 2006 (5.3%) | 1834 (4.9%) |
| Medium Stay (3-7 days) | 792 (10.5%) | 1966 (5.2%) | 2013 (5.4%) | 1974 (5.3%) |
| Long Stay (8-30 days) | 854 (11.4%) | 1137 (3.0%) | 1193 (3.2%) | 1187 (3.2%) |
| Extended Stay (31+ days) | 384 (5.1%) | 228 (0.6%) | 227 (0.6%) | 175 (0.5%) |
| COVID treatment |  |  |  |  |
| Corticosteroids^a^ | 1841 (24.5%) | 2752 (7.3%) | 2667 (7.1%) | 2636 (7.0%) |
| Remdisivir^a^ | 1261 (16.8%) | 1700 (4.5%) | 1585 (4.2%) | 1584 (4.2%) |
| Vasopressors^a^ | 464 (6.2%) | 617 (1.6%) | 674 (1.8%) | 659 (1.8%) |
| ECMO^a^ | 56 (0.7%) | 29 (0.1%) | 21 (0.1%) | <20 |
| Mechanical Ventilation^a^ | 528 (7.0%) | 434 (1.2%) | 413 (1.1%) | 369 (1.0%) |
| AKI during COVID-associated Hospitalization | 623 (8.3%) | 929 (2.5%) | 974 (2.6%) | 944 (2.5%) |
| Sepsis during COVID-associated Hospitalization | 580 (7.7%) | 796 (2.1%) | 761 (2.0%) | 793 (2.1%) |

^a^Only captured for individuals hospitalized for COVID-19

**eTable 15. PASC Risk Factors from Logistic Regression (PASC defined as U09.9)**

|  | Method 1  Unrestricted Controls  (N=45090) | Method 2  Restricted  controls  (N=45072) | Method 3  Most Restricted  controls  (N=45072) |
| --- | --- | --- | --- |
| **Demographics** |  |  |  |
| Age |  |  |  |
| 18-29 | REF | REF | REF |
| 30-39 | 1.76 (1.58-1.96) | 1.91 (1.71-2.13) | 1.84 (1.65-2.05) |
| 40-49 | 2.46 (2.21-2.73) | 2.95 (2.66-3.28) | 2.62 (2.36-2.92) |
| 50-59 | 2.63 (2.37-2.93) | 3.09 (2.78-3.44) | 2.78 (2.5-3.09) |
| 60-69 | 2.32 (2.08-2.6) | 2.87 (2.56-3.22) | 2.4 (2.15-2.69) |
| 70-79 | 1.89 (1.66-2.15) | 2.23 (1.96-2.54) | 1.97 (1.73-2.25) |
| 80-89 | 1.61 (1.35-1.92) | 1.83 (1.54-2.18) | 1.63 (1.36-1.95) |
| 90+ | 1.26 (0.88-1.8) | 1.26 (0.89-1.79) | 1.19 (0.83-1.7) |
| Sex |  |  |  |
| Male or Unknown Sex | REF | REF | REF |
| Female | 1.38 (1.31-1.46) | 1.32 (1.24-1.39) | 1.25 (1.18-1.32) |
| Race/ethnicity |  |  |  |
| White NH | REF | REF | REF |
| Hispanic | 0.78 (0.71-0.85) | 0.82 (0.75-0.9) | 0.86 (0.78-0.94) |
| Black NH | 0.77 (0.71-0.84) | 0.77 (0.71-0.84) | 0.81 (0.75-0.88) |
| Asian NH | 0.85 (0.69-1.05) | 0.75 (0.61-0.92) | 0.79 (0.64-0.98) |
| Other race NH | 0.92 (0.67-1.27) | 0.95 (0.69-1.31) | 1.0 (0.73-1.38) |
| **Comorbidities Prior to COVID Index Date** |  |  |  |
| AKI | 0.93 (0.79-1.09) | 0.91 (0.78-1.07) | 0.84 (0.71-0.98) |
| Cardiomyopathies | 0.92 (0.76-1.11) | 0.83 (0.7-1.0) | 0.85 (0.71-1.02) |
| Cerebrovascular Disease | 0.88 (0.76-1.01) | 0.84 (0.73-0.96) | 0.86 (0.75-0.98) |
| Chronic Lung Disease | 1.68 (1.57-1.79) | 1.57 (1.47-1.67) | 1.6 (1.5-1.7) |
| Complicated Diabetes | 1.03 (0.9-1.17) | 1.07 (0.94-1.21) | 1.0 (0.88-1.14) |
| Congestive Heart Failure | 0.87 (0.68-1.11) | 0.86 (0.68-1.08) | 0.87 (0.69-1.1) |
| Coronary Artery Disease | 1.03 (0.92-1.15) | 0.97 (0.87-1.08) | 0.96 (0.86-1.07) |
| Dementia | 0.84 (0.68-1.03) | 0.79 (0.65-0.96) | 0.88 (0.72-1.08) |
| Down Syndrome | 7.49 (0.74-75.47) | 4.44 (0.66-30.07) | 11.54 (1.14-116.27) |
| Heart Failure | 1.16 (0.93-1.44) | 1.24 (1.01-1.54) | 1.15 (0.93-1.42) |
| Hemiplegia or Paraplegia | 0.66 (0.48-0.91) | 0.55 (0.4-0.74) | 0.64 (0.47-0.88) |
| HIV | 0.85 (0.59-1.22) | 0.9 (0.63-1.28) | 0.74 (0.52-1.05) |
| Hypertension | 1.17 (1.09-1.25) | 0.96 (0.9-1.03) | 0.98 (0.91-1.04) |
| Kidney Disease | 1.01 (0.89-1.15) | 0.99 (0.87-1.13) | 1.04 (0.92-1.18) |
| Malignant Cancer | 1.07 (0.97-1.18) | 0.87 (0.79-0.96) | 0.91 (0.83-1.0) |
| Metastatic Solid Tumor Cancers | 0.68 (0.52-0.88) | 0.65 (0.51-0.84) | 0.64 (0.49-0.82) |
| Mild Liver Disease | 0.91 (0.74-1.13) | 0.9 (0.73-1.1) | 0.91 (0.74-1.12) |
| Moderate to Severe Liver Disease | 0.74 (0.54-1.01) | 0.7 (0.52-0.94) | 0.67 (0.49-0.9) |
| Myocardial Infarction | 0.76 (0.65-0.88) | 0.78 (0.67-0.9) | 0.85 (0.74-0.99) |
| Obesity | 1.28 (1.21-1.36) | 1.05 (0.99-1.11) | 1.07 (1.01-1.14) |
| Peptic Ulcer | 1.17 (1.0-1.37) | 1.13 (0.98-1.32) | 1.11 (0.96-1.29) |
| Perpheral Vascular Disease | 1.24 (1.07-1.43) | 1.1 (0.96-1.26) | 1.14 (0.99-1.31) |
| Rheumatologic Disease | 1.29 (1.11-1.49) | 1.24 (1.08-1.42) | 1.18 (1.03-1.36) |
| Sickle Cell Disease | 1.45 (0.77-2.73) | 0.85 (0.48-1.52) | 0.76 (0.42-1.36) |
| Systemic Corticosteroids | 1.36 (1.29-1.45) | 1.16 (1.1-1.23) | 1.19 (1.12-1.26) |
| Thalassemia | 1.66 (0.94-2.94) | 1.07 (0.63-1.8) | 1.22 (0.72-2.07) |
| Tuberculosis | 1.17 (0.71-1.92) | 1.12 (0.7-1.77) | 1.48 (0.92-2.36) |
| Uncomplicated Diabetes | 0.85 (0.76-0.95) | 0.83 (0.74-0.92) | 0.84 (0.76-0.94) |
| **Behavioral Health** |  |  |  |
| Depression | 1.56 (1.46-1.67) | 1.32 (1.24-1.41) | 1.28 (1.2-1.37) |
| Psychosis | 0.6 (0.45-0.81) | 0.58 (0.44-0.78) | 0.56 (0.42-0.76) |
| Substance Use | 0.65 (0.55-0.78) | 0.55 (0.47-0.65) | 0.65 (0.55-0.76) |
| Tobacco Smoker | 0.74 (0.66-0.83) | 0.7 (0.63-0.78) | 0.69 (0.62-0.76) |
| **Characteristics of Index COVID "Acute Phase"** |  |  |  |
| COVID Diagnosis during COVID-associated Hospitalization | 0.5 (0.45-0.57) | 0.65 (0.58-0.73) | 0.65 (0.58-0.73) |
| COVID-associated Hospitalization | 3.49 (2.67-4.56) | 2.85 (2.21-3.67) | 2.55 (1.99-3.27) |
| COVID-associated ED Visit | 1.27 (1.18-1.36) | 1.48 (1.38-1.59) | 1.51 (1.41-1.62) |
| Hospitalization stay |  |  |  |
| Not Hospitalized | REF | REF | REF |
| Short Stay | 0.78 (0.59-1.04) | 0.76 (0.58-1.0) | 0.96 (0.73-1.25) |
| Medium Stay | 0.81 (0.6-1.08) | 0.79 (0.6-1.05) | 1.0 (0.76-1.31) |
| Long Stay | 1.51 (1.12-2.03) | 1.42 (1.06-1.89) | 1.76 (1.33-2.34) |
| Extended Stay | 2.82 (1.95-4.08) | 2.52 (1.76-3.62) | 4.34 (3.02-6.25) |
| COVID treatment |  |  |  |
| Corticosteroidsᵃ | 1.19 (1.05-1.35) | 1.42 (1.25-1.61) | 1.37 (1.21-1.55) |
| Remdisivirᵃ | 1.38 (1.22-1.57) | 1.51 (1.34-1.71) | 1.43 (1.27-1.62) |
| Vasopressorsᵃ | 0.7 (0.58-0.83) | 0.67 (0.57-0.8) | 0.64 (0.54-0.77) |
| ECMOᵃ | 1.51 (0.92-2.49) | 2.04 (1.17-3.55) | 2.15 (1.18-3.89) |
| Medical Ventilationᵃ | 1.5 (1.22-1.84) | 1.61 (1.31-1.98) | 1.66 (1.35-2.04) |
| AKI during COVID-associated Hospitalization | 1.07 (0.91-1.25) | 1.13 (0.97-1.32) | 1.16 (0.99-1.35) |
| Sepsis during COVID-associated Hospitalization | 0.92 (0.8-1.07) | 0.98 (0.84-1.13) | 0.84 (0.73-0.98) |

ᵃOnly captured for individuals hospitalized for COVID-19

Odds ratios presented with 95% CI in parenthesis

**eTable 16. Characteristics of PASC defined as Long-COVID Clinic Visits**

|  | Long-COVID Clinic Visit Only  (N=1241) | Method 1  Unrestricted controls (N=6205) | Method 2  Restricted controls (N=6205) | Method 3  Most restricted controls (N=6205) |
| --- | --- | --- | --- | --- |
| **Demographics** |  |  |  |  |
| Age |  |  |  |  |
| 18-29 | 86 (6.9%) | 1105 (17.8%) | 1123 (18.1%) | 1155 (18.6%) |
| 30-39 | 211 (17.0%) | 1110 (17.9%) | 1025 (16.5%) | 1027 (16.6%) |
| 40-49 | 274 (22.1%) | 1109 (17.9%) | 1055 (17.0%) | 980 (15.8%) |
| 50-59 | 307 (24.7%) | 1094 (17.6%) | 1053 (17.0%) | 1069 (17.2%) |
| 60-69 | 230 (18.5%) | 907 (14.6%) | 971 (15.6%) | 969 (15.6%) |
| 70-79 | 104 (8.4%) | 580 (9.3%) | 613 (9.9%) | 610 (9.8%) |
| 80-89 | 29 (2.3%) | 236 (3.8%) | 287 (4.6%) | 308 (5.0%) |
| 90+ | 0 (0.0%) | 64 (1.0%) | 78 (1.3%) | 87 (1.4%) |
| Sex |  |  |  |  |
| Female | 776 (62.5%) | 3438 (55.4%) | 3642 (58.7%) | 3688 (59.4%) |
| Male | 464 (37.4%) | 2757 (44.4%) | 2561 (41.3%) | 2512 (40.5%) |
| Race/ethnicity |  |  |  |  |
| White non-Hispanic (NH) | 699 (56.3%) | 3482 (56.1%) | 3618 (58.3%) | 3665 (59.1%) |
| Hispanic | 176 (14.2%) | 948 (15.3%) | 823 (13.3%) | 850 (13.7%) |
| Black NH | 267 (21.5%) | 1181 (19.0%) | 1334 (21.5%) | 1254 (20.2%) |
| Asian NH | 26 (2.1%) | 188 (3.0%) | 156 (2.5%) | 171 (2.8%) |
| Other race NH | <20 | <20 | <20 | 22 (0.4%) |
| **Comorbidities Prior to COVID Index Date** |  |  |  |  |
| AKI | 71 (5.7%) | 278 (4.5%) | 301 (4.9%) | 290 (4.7%) |
| Cardiomyopathies | 32 (2.6%) | 97 (1.6%) | 141 (2.3%) | 112 (1.8%) |
| Cerebrovascular Disease | 35 (2.8%) | 164 (2.6%) | 204 (3.3%) | 196 (3.2%) |
| Chronic Lung Disease | 275 (22.2%) | 731 (11.8%) | 896 (14.4%) | 799 (12.9%) |
| Complicated Diabetes | 126 (10.2%) | 474 (7.6%) | 598 (9.6%) | 535 (8.6%) |
| Congestive Heart Failure | 76 (6.1%) | 194 (3.1%) | 261 (4.2%) | 231 (3.7%) |
| Coronary Artery Disease | 90 (7.3%) | 336 (5.4%) | 423 (6.8%) | 400 (6.4%) |
| Dementia | <20 | 83 (1.3%) | 125 (2.0%) | 128 (2.1%) |
| Down Syndrome | 0 (0.0%) | <20 | 0 (0.0%) | 0 (0.0%) |
| Heart Failure | 97 (7.8%) | 259 (4.2%) | 341 (5.5%) | 302 (4.9%) |
| Hemiplegia or Paraplegia | <20 | 28 (0.5%) | 36 (0.6%) | 37 (0.6%) |
| HIV | <20 | 32 (0.5%) | 40 (0.6%) | 34 (0.5%) |
| Hypertension | 371 (29.9%) | 1446 (23.3%) | 1780 (28.7%) | 1710 (27.6%) |
| Kidney Disease | 119 (9.6%) | 450 (7.3%) | 566 (9.1%) | 512 (8.3%) |
| Malignant Cancer | 91 (7.3%) | 444 (7.2%) | 536 (8.6%) | 485 (7.8%) |
| Metastatic Solid Tumor Cancers | <20 | 60 (1.0%) | 73 (1.2%) | 69 (1.1%) |
| Mild Liver Disease | <20 | 61 (1.0%) | 99 (1.6%) | 75 (1.2%) |
| Moderate to Severe Liver Disease | <20 | 25 (0.4%) | 40 (0.6%) | 29 (0.5%) |
| Myocardial Infarction | 32 (2.6%) | 169 (2.7%) | 213 (3.4%) | 194 (3.1%) |
| Obesity | 643 (51.8%) | 2418 (39.0%) | 2923 (47.1%) | 2798 (45.1%) |
| Peptic Ulcer | 21 (1.7%) | 79 (1.3%) | 114 (1.8%) | 99 (1.6%) |
| Perpheral Vascular Disease | 42 (3.4%) | 149 (2.4%) | 197 (3.2%) | 170 (2.7%) |
| Rheumatologic Disease | 33 (2.7%) | 108 (1.7%) | 138 (2.2%) | 113 (1.8%) |
| Sickle Cell Disease | <20 | <20 | <20 | <20 |
| Systemic Corticosteroids | 491 (39.6%) | 1816 (29.3%) | 2147 (34.6%) | 2010 (32.4%) |
| Thalassemia | <20 | <20 | <20 | <20 |
| Tuberculosis | <20 | <20 | <20 | <20 |
| Uncomplicated Diabetes | 189 (15.2%) | 740 (11.9%) | 898 (14.5%) | 856 (13.8%) |
| **Behavioral Health** |  |  |  |  |
| Depression | 218 (17.6%) | 630 (10.2%) | 844 (13.6%) | 780 (12.6%) |
| Psychosis | <20 | 47 (0.8%) | 62 (1.0%) | 55 (0.9%) |
| Substance Abuse | <20 | 145 (2.3%) | 171 (2.8%) | 165 (2.7%) |
| Tobacco Smoker | 21 (1.7%) | 269 (4.3%) | 348 (5.6%) | 297 (4.8%) |
| **Characteristics of Index COVID "Acute Phase"** |  |  |  |  |
| COVID Diagnosis during COVID-associated Hospitalization | 394 (31.7%) | 754 (12.2%) | 793 (12.8%) | 756 (12.2%) |
| COVID-associated Hospitalization | 547 (44.1%) | 1089 (17.6%) | 1167 (18.8%) | 1085 (17.5%) |
| COVID-associated ED Visit | 167 (13.5%) | 1098 (17.7%) | 978 (15.8%) | 982 (15.8%) |
| Hospitalization Stay |  |  |  |  |
| Short Stay (0-2 days) | 76 (6.1%) | 383 (6.2%) | 404 (6.5%) | 374 (6.0%) |
| Medium Stay (3-7 days) | 106 (8.5%) | 369 (5.9%) | 395 (6.4%) | 352 (5.7%) |
| Long Stay (8-30 days) | 220 (17.7%) | 214 (3.4%) | 215 (3.5%) | 228 (3.7%) |
| Extended Stay (31+ days) | 86 (6.9%) | 32 (0.5%) | 52 (0.8%) | 46 (0.7%) |
| COVID Treatment |  |  |  |  |
| Corticosteroids^a^ | 272 (21.9%) | 475 (7.7%) | 503 (8.1%) | 463 (7.5%) |
| Remdisivir^a^ | 217 (17.5%) | 335 (5.4%) | 325 (5.2%) | 300 (4.8%) |
| Vasopressors^a^ | 170 (13.7%) | 114 (1.8%) | 149 (2.4%) | 132 (2.1%) |
| ECMO^a^ | <20 | <20 | <20 | <20 |
| Mechanical Ventilation^a^ | 121 (9.8%) | 64 (1.0%) | 74 (1.2%) | 69 (1.1%) |
| AKI during COVID-associated Hospitalization | 67 (5.4%) | 122 (2.0%) | 145 (2.3%) | 119 (1.9%) |
| Sepsis during COVID-associated Hospitalization | 68 (5.5%) | 92 (1.5%) | 121 (2.0%) | 102 (1.6%) |

**eTable 17. PASC Risk Factors from Logistic Regression (PASC defined as Long-COVID**

**Clinic Visits)**

|  | Method 1  Unrestricted Controls  (N=7446) | Method 2  Restricted  controls  (N=7446) | Method 3  Most Restricted  controls  (N=7446) |
| --- | --- | --- | --- |
| **Demographics** |  |  |  |
| Age |  |  |  |
| 18-29 | REF | REF | REF |
| 30-39 | 2.24 (1.7-2.95) | 2.6 (1.98-3.42) | 2.69 (2.04-3.55) |
| 40-49 | 2.63 (2.01-3.45) | 3.14 (2.4-4.11) | 3.53 (2.69-4.62) |
| 50-59 | 2.57 (1.96-3.38) | 3.23 (2.46-4.25) | 3.18 (2.42-4.17) |
| 60-69 | 1.94 (1.45-2.6) | 2.29 (1.72-3.06) | 2.09 (1.56-2.79) |
| 70-79 | 1.22 (0.86-1.72) | 1.46 (1.04-2.07) | 1.48 (1.05-2.08) |
| 80-89 | 0.69 (0.41-1.15) | 0.59 (0.36-0.99) | 0.53 (0.32-0.88) |
| 90+ | 0.0 (0.0-inf) | 0.0 (0.0-inf) | 0.0 (0.0-inf) |
| Sex |  |  |  |
| Male or Unknown Sex | REF | REF | REF |
| Female | 1.49 (1.29-1.72) | 1.35 (1.16-1.55) | 1.23 (1.06-1.42) |
| Race/ethnicity |  |  |  |
| White NH | REF | REF | REF |
| Hispanic | 0.68 (0.55-0.84) | 0.75 (0.61-0.93) | 0.71 (0.58-0.88) |
| Black NH | 0.77 (0.65-0.93) | 0.67 (0.56-0.8) | 0.72 (0.6-0.87) |
| Asian NH | 0.57 (0.36-0.89) | 0.61 (0.38-0.97) | 0.57 (0.36-0.91) |
| Other race NH | 0.76 (0.15-3.7) | 0.49 (0.09-2.61) | 0.33 (0.07-1.54) |
| **Comorbidities Prior to COVID Index Date** |  |  |  |
| AKI | 0.49 (0.28-0.84) | 0.72 (0.42-1.21) | 0.59 (0.34-1.03) |
| Cardiomyopathies | 1.2 (0.7-2.04) | 0.9 (0.55-1.48) | 1.01 (0.61-1.67) |
| Cerebrovascular Disease | 0.71 (0.45-1.11) | 0.87 (0.56-1.34) | 0.88 (0.57-1.37) |
| Chronic Lung Disease | 1.48 (1.22-1.79) | 1.52 (1.26-1.83) | 1.67 (1.39-2.02) |
| Complicated Diabetes | 0.87 (0.6-1.26) | 0.79 (0.55-1.13) | 0.92 (0.63-1.32) |
| Congestive Heart Failure | 1.6 (0.8-3.17) | 1.33 (0.69-2.58) | 1.67 (0.85-3.29) |
| Coronary Artery Disease | 1.67 (1.21-2.31) | 1.56 (1.14-2.14) | 1.41 (1.03-1.94) |
| Dementia | 0.98 (0.54-1.8) | 0.96 (0.53-1.73) | 0.83 (0.46-1.51) |
| Heart Failure | 0.78 (0.42-1.43) | 0.97 (0.54-1.76) | 0.87 (0.47-1.61) |
| Hemiplegia or Paraplegia | 0.84 (0.3-2.33) | 0.7 (0.27-1.8) | 0.53 (0.2-1.38) |
| HIV | 1.57 (0.73-3.4) | 1.71 (0.84-3.49) | 1.64 (0.79-3.39) |
| Hypertension | 1.14 (0.94-1.37) | 1.0 (0.84-1.2) | 0.99 (0.82-1.18) |
| Kidney Disease | 1.06 (0.72-1.57) | 0.87 (0.61-1.25) | 0.88 (0.61-1.27) |
| Malignant Cancer | 0.89 (0.67-1.19) | 0.81 (0.62-1.07) | 0.9 (0.68-1.19) |
| Metastatic Solid Tumor Cancers | 0.42 (0.17-1.01) | 0.45 (0.19-1.05) | 0.5 (0.21-1.19) |
| Mild Liver Disease | 1.13 (0.58-2.21) | 0.77 (0.41-1.44) | 0.89 (0.47-1.68) |
| Moderate to Severe Liver Disease | 0.57 (0.2-1.62) | 0.56 (0.21-1.49) | 0.84 (0.29-2.4) |
| Myocardial Infarction | 0.44 (0.27-0.73) | 0.46 (0.28-0.73) | 0.51 (0.32-0.82) |
| Obesity | 1.07 (0.92-1.25) | 0.86 (0.74-0.99) | 0.9 (0.78-1.05) |
| Peptic Ulcer | 1.0 (0.57-1.75) | 1.07 (0.63-1.79) | 0.93 (0.54-1.59) |
| Perpheral Vascular Disease | 1.29 (0.82-2.03) | 1.19 (0.78-1.83) | 1.46 (0.94-2.26) |
| Rheumatologic Disease | 1.01 (0.65-1.59) | 0.95 (0.61-1.47) | 1.51 (0.97-2.35) |
| Sickle Cell Disease | 0.87 (0.2-3.85) | 0.56 (0.11-2.96) | 1.57 (0.31-7.89) |
| Systemic Corticosteroids | 1.12 (0.95-1.31) | 0.99 (0.84-1.15) | 1.04 (0.9-1.22) |
| Thalassemia | 1.46 (0.36-5.85) | 0.77 (0.17-3.54) | 4.86 (1.08-21.88) |
| Tuberculosis | 1.55 (0.46-5.18) | 3.46 (1.03-11.61) | 1.44 (0.47-4.42) |
| Uncomplicated Diabetes | 0.79 (0.58-1.07) | 0.86 (0.64-1.16) | 0.77 (0.57-1.04) |
| **Behavioral Health** |  |  |  |
| Depression | 1.82 (1.49-2.22) | 1.51 (1.25-1.83) | 1.6 (1.32-1.94) |
| Psychosis | 0.4 (0.16-1.04) | 0.25 (0.1-0.64) | 0.39 (0.15-1.04) |
| Substance Abuse | 0.6 (0.34-1.04) | 0.66 (0.39-1.13) | 0.48 (0.27-0.83) |
| Tobacco Smoker | 0.27 (0.16-0.45) | 0.27 (0.17-0.44) | 0.32 (0.2-0.52) |
| **Characteristics of Index COVID "Acute Phase"** |  |  |  |
| COVID Diagnosis during COVID-associated Hospitalization | 1.0 (0.75-1.32) | 1.13 (0.86-1.5) | 1.01 (0.76-1.34) |
| COVID-associated Hospitalization | 5.04 (3.47-7.32) | 4.62 (3.22-6.63) | 6.21 (4.26-9.05) |
| COVID-associated ED Visit | 0.85 (0.7-1.03) | 1.01 (0.83-1.24) | 0.98 (0.81-1.2) |
| Hospitalization stay |  |  |  |
| Not Hospitalized | REF | REF | REF |
| Short Stay | 0.34 (0.22-0.54) | 0.35 (0.22-0.54) | 0.29 (0.18-0.46) |
| Medium Stay | 0.56 (0.35-0.88) | 0.55 (0.35-0.87) | 0.47 (0.29-0.75) |
| Long Stay | 2.16 (1.35-3.47) | 2.25 (1.41-3.59) | 1.73 (1.07-2.8) |
| Extended Stay | 4.05 (2.02-8.14) | 2.43 (1.24-4.74) | 2.68 (1.35-5.33) |
| COVID treatment |  |  |  |
| Corticosteroidsᵃ | 0.76 (0.55-1.04) | 0.76 (0.56-1.03) | 0.67 (0.49-0.92) |
| Remdesivirᵃ | 1.15 (0.83-1.6) | 1.35 (0.98-1.86) | 1.73 (1.25-2.4) |
| Vasopressorsᵃ | 1.51 (1.0-2.29) | 1.38 (0.94-2.04) | 1.63 (1.09-2.46) |
| ECMOᵃ | 1.82 (0.56-5.94) | 2.4 (0.75-7.68) | 1.81 (0.6-5.45) |
| Mechanical Ventilationᵃ | 1.38 (0.83-2.3) | 1.43 (0.87-2.36) | 0.99 (0.6-1.66) |
| AKI during COVID-associated Hospitalization | 0.95 (0.57-1.6) | 1.08 (0.66-1.79) | 1.08 (0.64-1.83) |
| Sepsis during COVID-associated Hospitalization | 0.64 (0.41-0.99) | 0.49 (0.32-0.75) | 0.55 (0.36-0.85) |

ᵃOnly captured individuals hospitalized for COVID-19

Odds ratios presented with 95% CI in parenthesis

**eTable 18. Comparison of U09.9 and Long-COVID Visit Cohorts**

|  | U09.9 OR Long-COVID Visit (N=8325) | U09.9 Only (N=7512) | Long-COVID Clinic Visit Only (N=1241) |
| --- | --- | --- | --- |
| **Demographics** |  |  |  |
| Age |  |  |  |
| 18-29 | 630 (7.6%) | 577 (7.7%) | 86 (6.9%) |
| 30-39 | 1229 (14.8%) | 1090 (14.5%) | 211 (17.0%) |
| 40-49 | 1749 (21.0%) | 1580 (21.0%) | 274 (22.1%) |
| 50-59 | 1933 (23.2%) | 1738 (23.1%) | 307 (24.7%) |
| 60-69 | 1605 (19.3%) | 1450 (19.3%) | 230 (18.5%) |
| 70-79 | 840 (10.1%) | 766 (10.2%) | 104 (8.4%) |
| 80-89 | 293 (3.5%) | 268 (3.6%) | 29 (2.3%) |
| 90+ | 37 (0.4%) | 43 (0.6%) | 0 (0.0%) |
| Sex |  |  |  |
| Female | 5225 (62.8%) | 4752 (63.3%) | 776 (62.5%) |
| Male | 3096 (37.2%) | 2759 (36.7%) | 464 (37.4%) |
| Race/ethnicity |  |  |  |
| White non-Hispanic (NH) | 5707 (68.6%) | 5301 (70.6%) | 699 (56.3%) |
| Hispanic | 835 (10.0%) | 711 (9.5%) | 176 (14.2%) |
| Black NH | 1235 (14.8%) | 1035 (13.8%) | 267 (21.5%) |
| Asian NH | 136 (1.6%) | 116 (1.5%) | 26 (2.1%) |
| Other race NH | 54 (0.6%) | 52 (0.7%) | <20 |
| **Comorbidities Prior to COVID Index Date** |  |  |  |
| AKI | 862 (10.4%) | 814 (10.8%) | 71 (5.7%) |
| Cardiomyopathies | 225 (2.7%) | 208 (2.8%) | 32 (2.6%) |
| Cerebrovascular Disease | 390 (4.7%) | 364 (4.8%) | 35 (2.8%) |
| Chronic Lung Disease | 2404 (28.9%) | 2245 (29.9%) | 275 (22.2%) |
| Complicated Diabetes | 1210 (14.5%) | 1136 (15.1%) | 126 (10.2%) |
| Congestive Heart Failure | 573 (6.9%) | 522 (6.9%) | 76 (6.1%) |
| Coronary Artery Disease | 832 (10.0%) | 778 (10.4%) | 90 (7.3%) |
| Dementia | 153 (1.8%) | 145 (1.9%) | <20 |
| Down Syndrome | <20 | <20 | 0 (0.0%) |
| Heart Failure | 737 (8.9%) | 669 (8.9%) | 97 (7.8%) |
| Hemiplegia or Paraplegia | 61 (0.7%) | 58 (0.8%) | <20 |
| HIV | 51 (0.6%) | 41 (0.5%) | <20 |
| Hypertension | 3365 (40.4%) | 3145 (41.9%) | 371 (29.9%) |
| Kidney Disease | 1262 (15.2%) | 1183 (15.7%) | 119 (9.6%) |
| Malignant Cancer | 837 (10.1%) | 783 (10.4%) | 91 (7.3%) |
| Metastatic Solid Tumor Cancers | 91 (1.1%) | 87 (1.2%) | <20 |
| Mild Liver Disease | 170 (2.0%) | 159 (2.1%) | <20 |
| Moderate to Severe Liver Disease | 82 (1.0%) | 77 (1.0%) | <20 |
| Myocardial Infarction | 392 (4.7%) | 371 (4.9%) | 32 (2.6%) |
| Obesity | 4691 (56.4%) | 4279 (57.0%) | 643 (51.8%) |
| Peptic Ulcer | 279 (3.4%) | 270 (3.6%) | 21 (1.7%) |
| Perpheral Vascular Disease | 405 (4.9%) | 388 (5.2%) | 42 (3.4%) |
| Rheumatologic Disease | 350 (4.2%) | 334 (4.4%) | 33 (2.7%) |
| Sickle Cell Disease | <20 | <20 | <20 |
| Systemic Corticosteroids | 4325 (52.0%) | 4015 (53.4%) | 491 (39.6%) |
| Thalassemia | 21 (0.3%) | <20 | <20 |
| Tuberculosis | 27 (0.3%) | 24 (0.3%) | <20 |
| Uncomplicated Diabetes | 1708 (20.5%) | 1589 (21.2%) | 189 (15.2%) |
| **Behavioral Health** |  |  |  |
| Depression | 2059 (24.7%) | 1946 (25.9%) | 218 (17.6%) |
| Psychosis | 65 (0.8%) | 61 (0.8%) | <20 |
| Substance Abuse | 205 (2.5%) | 196 (2.6%) | <20 |
| Tobacco Smoker | 515 (6.2%) | 504 (6.7%) | 21 (1.7%) |
| **Characteristics of Index COVID "Acute Phase"** |  |  |  |
| COVID Diagnosis during COVID-associated Hospitalization | 2065 (24.8%) | 1757 (23.4%) | 394 (31.7%) |
| COVID-associated Hospitalization | 3100 (37.3%) | 2683 (35.7%) | 547 (44.1%) |
| COVID-associated ED Visit | 1564 (18.8%) | 1479 (19.7%) | 167 (13.5%) |
| Hospitalization Stay |  |  |  |
| Short Stay | 610 (7.3%) | 560 (7.5%) | 76 (6.1%) |
| Medium Stay | 870 (10.5%) | 792 (10.5%) | 106 (8.5%) |
| Long Stay | 1029 (12.4%) | 854 (11.4%) | 220 (17.7%) |
| Extended Stay | 449 (5.4%) | 384 (5.1%) | 86 (6.9%) |
| COVID treatment |  |  |  |
| Corticosteroidsᵃ | 2025 (24.3%) | 1841 (24.5%) | 272 (21.9%) |
| Remdisivirᵃ | 1409 (16.9%) | 1261 (16.8%) | 217 (17.5%) |
| Vasopressorsᵃ | 601 (7.2%) | 464 (6.2%) | 170 (13.7%) |
| ECMOᵃ | 66 (0.8%) | 56 (0.7%) | <20 |
| Mechanical Ventilationᵃ | 615 (7.4%) | 528 (7.0%) | 121 (9.8%) |
| AKI during COVID-associated Hospitalization | 664 (8.0%) | 623 (8.3%) | 67 (5.4%) |
| Sepsis during COVID-associated Hospitalization | 614 (7.4%) | 580 (7.7%) | 68 (5.5%) |

ᵃOnly captured for individuals hospitalized with COVID-19
